# Multiplexed detection of febrile infections using CARMEN

**DOI:** 10.1101/2024.07.15.24310364

**Authors:** M. Kamariza, K. McMahon, L. Kim, N.L. Welch, L. Stenson, L. Allan-Blitz, G. Sanders, P. Eromon, A.M. Iluoreh, A. Sijuwola, O.O. Ope-ewe, A.O. Ayinla, C. l’Anson, I. Baudi, M.F. Paye, C. Wilkason, J. Lemieux, A. Ozonoff, E. Stachler, C.T. Happi, P.C. Sabeti

## Abstract

Detection and diagnosis of bloodborne pathogens are critical for patients and for preventing outbreaks, yet challenging due to these diseases’ nonspecific initial symptoms. We advanced CRISPR-based Combinatorial Arrayed Reactions for Multiplexed Evaluation of Nucleic acids (CARMEN) technology for simultaneous detection of pathogens on numerous samples. We developed three specialized panels that target viral hemorrhagic fevers, mosquito-borne viruses, and sexually transmitted infections, collectively identifying 23 pathogens. We used deep learning to design CARMEN assays with enhanced sensitivity and specificity, validating them and evaluating their performance on synthetic targets, spiked healthy normal serum samples, and patient samples for *Neisseria gonorrhoeae* in the United States and for Lassa and mpox virus in Nigeria. Our results show multiplexed CARMEN assays match or outperform individual assay RT-PCR in sensitivity, with matched specificity. These findings underscore CARMEN’s potential as a highly effective tool for rapid, accurate pathogen detection for clinical diagnosis and public health surveillance.

## Introduction

Bloodborne pathogens (BBPs) cause major disruptions to human life and public health. While the prevalence of bloodborne viruses varies widely across the world, BBPs harbor some of the deadliest diseases driven by hemorrhagic fevers (e.g., Ebola, Lassa). These diseases are often difficult to diagnose because early symptoms, including fever, vomiting, and aches, are indistinguishable from each other^1–3^. Similarly, sexually-transmitted infections (STIs) (e.g. HIV, mpox, Hepatitis B) are endemic throughout the world, and present a particularly significant burden on morbidity and mortality in low- or middle-income countries (LMICs)^4–6^. Furthermore, such settings often lack the required infrastructure to support pathogen diagnosis, and instead rely on syndromic management of symptomatic cases. Thus, rapid and accurate diagnostic testing is needed to control the disease spread.

The gold standard for diagnosing BBPs and STIs are polymerase chain reaction (PCR)- based methods such as reverse transcription quantitative polymerase chain reaction (RT-qPCR) due to their high sensitivity and specificity^7^. However, these methods present several limitations, such as low throughput, high cost, and complexity. In particular, the time-consuming nature of RT-qPCRs for each test has led to a growing interest in developing alternative diagnostic tools that are equally robust yet simpler, faster, and more cost-effective^8,9^.

Moreover, an ideal diagnostic and surveillance technology should be capable of processing hundreds of patient samples simultaneously and detecting multiple pathogens concurrently. Indeed, circulating and emerging diseases require widespread deployability of molecular tests for rapidly diagnosing and surveilling various pathogens. The recent SARS-CoV-2 viral outbreak emphasized the challenges of detecting known and potentially unknown coronaviruses, rapidly-evolving viral variants, as well as distinguishing related viruses that trigger similar symptoms^10,11^.

CRISPR-based technologies, specifically the CRISPR-Cas13 system, have shown promise in diagnosing viral infections, including COVID-19^12–14^. The CRISPR-Cas13 assay is an RNA-targeting CRISPR enzyme system that can be programmed to detect specific RNA sequences, including viral RNA^13^. Upon recognition of the target RNA, the Cas13 endonuclease cleaves non-target RNAs, leading to a detectable fluorescent signal^12^. The CRISPR-Cas13-based Specific High Sensitivity Enzymatic Reporter UnLOCKing (SHERLOCK) diagnostic assay was shown to detect viral particles as low as 1.25 cp/μl, which in this instance was found to be of higher sensitivity compared to standard RT-PCR^14^. Using SHERLOCK, researchers achieved 100% specificity and 100% sensitivity with a limit of detection (LOD) of 42 RNA copies per reaction using the SHERLOCK system on 534 clinical samples of SARS-CoV-2 RNA^15^. This system demonstrated robust performance by integrating an internal control for ribonuclease contamination which enhanced the test’s suitability for resource-limited settings with an elevated risk of RNAse contamination. These Cas13-based approaches inherently offer multiplexing advantages, enabling targeting of a single viral genome at multiple sites, differentiation between related viruses or serotypes, and detection of different mutations within viral genomes^14,16,17^. Ultimately, the development of SHERLOCK established a foundation for sensitive and scalable pathogen detection.

We previously combined the power of CRISPR-Cas13 detection of amplified targets with the multiplexing capacity of microfluidics to create Combinatorial Arrayed Reactions for Multiplexed Evaluation of Nucleic acids (CARMEN)^18^. Leveraging our ability to test thousands of assay-target pairs with CARMEN, we created an automated design algorithm using deep learning, Activity-informed Design with All-inclusive Patrolling of Targets (ADAPT). ADAPT can identify optimal guide sequences for detecting pathogens with high specificity and sensitivity^19^. We further developed a microfluidic CARMEN (mCARMEN) system that uses Standard BioTools’ BiomarkHD instrument to enable automation and reduce labor^20^. This CRISPR-Cas13 detection system has enabled the multiplexed detection of SARS-CoV-2 RNA as well as influenza A and B viruses in a single reaction^18^. It does so with greater sensitivity and 100% specificity compared to RT-qPCR, and reduces the risk of false-positive results through demonstrating minimal cross-reactivity to non-target RNAs^20^. Additionally, the mCARMEN assay can distinguish between closely related SARS-CoV-2 variants, such as Delta and Omicron, with high accuracy, making it a valuable tool for surveillance and epidemiological studies^20^. With its superior multiplexing capacity, the (now collectively called) CARMEN assay allows for high-throughput analysis of clinical samples to reduce the time and cost of testing compared to traditional RT-PCR methods.

In this paper, we set out to optimize the CARMEN workflow by further reducing processing time and using simplified instrumentation suitable for use in routine clinical laboratories. Using this optimized CARMEN platform, we develop and evaluate three bloodborne pathogen (BBP) panels for large-scale and high-throughput detection and surveillance of viral hemorrhagic fevers (e.g. Ebola virus, Lassa virus), mosquito-borne viral diseases (e.g. West Nile virus, Yellow fever virus,), and sexually transmitted illnesses (e.g., Mpox virus, Hepatitis B virus).

Using ADAPT, we designed multiple crRNAs for each target to identify optimal guide sequences for detecting them with high specificity and sensitivity^19^. Then we tested the condensed panels using synthetic materials and validated them with seedstocks, totaling 23 targets – 9 targets for BBP panel 1, 10 targets for panel 2, and 7 targets for panel 3 (each including the human internal control, RNAse P). We also assessed the sensitivity and limit of detection (LOD) of BBP assays using healthy normal serum (HNS) samples spiked with known concentrations of viral seedstocks. Further, we performed comparative experiments against RT-qPCR using *Lassa virus* and *mpox virus* (formerly *monkeypox virus*) genomic materials spiked into HNS samples. Finally, we conducted validation tests using 10 *Neisseria gonorrhoeae*, 90 *lassa virus*, or 42 *mpox virus* confirmed-positive patient samples, benchmarked against commercial RT-qPCR kits. Ultimately, we present a versatile diagnostic pipeline capable of facilitating high-throughput pathogen identification and microbial surveillance, enabling early detection and prevention of infectious disease outbreaks.

## Results

### Optimization of CARMEN workflow with Fluidigm’s BiomarkX

We first sought to improve the CARMEN workflow to enable scalability for routine high-throughput and multiplexed analysis of patient samples in standard clinical laboratories. Previously, we described a microfluidic CARMEN (mCARMEN) system that uses Standard BioTools’ BiomarkHD instrument to enable automation and reduce labor^20^. Briefly, samples are extracted and amplified, followed by detection using integrated fluidic circuits (IFCs, **Figure S1**). Specifically, these microfluidic IFCs enable highly multiplexed detection by combining 192 samples with 24 detection assays or 96 samples with 96 detection assays, depending on the IFC. Subsequently, fluorescence is measured on the Fluidigm BiomarkHD using automated protocols, capturing images every 5 minutes for 1-3 hours at 37°C. Taken together, this workflow requires an estimated 5 to 6 hours.

To optimize this pipeline, we assessed whether the newer, simplified, model of the Biomark series, namely the BiomarkX, would improve the simplicity and workflow time of the BiomarkHD (**Figure 1A**). Indeed, the BiomarkX demonstrated several advantages important for our goals: (1) a smaller footprint compared to the BiomarkHD, (2) a user-friendly interface with a single button to launch data acquisition, and (3) a system that can be used with all types of integrated fluidic circuits (IFCs), and (3) a shorter runtime (total workflow requires ∼ 4 hours). Together, these advantages deem the BiomarkX adaptable for any given purpose and suggest its easy deployment in clinical laboratories. We tested our previously designed respiratory virus panel (RVP) using the BiomarkX and evaluated detection of the corresponding nine respiratory virus synthetic DNA targets^20^. Specifically, synthetic materials at dilutions ranging from 10^4^ to 10^0^ cp/µL were amplified by RT-PCR, followed by CARMEN fluorescence detection using BiomarkX (**Figure 1B**). We observed robust and sensitive detection of all nine targets included in the assay, demonstrating that detection performance was unaffected by the streamlined workflow with the BiomarkX.

**Figure 1.**
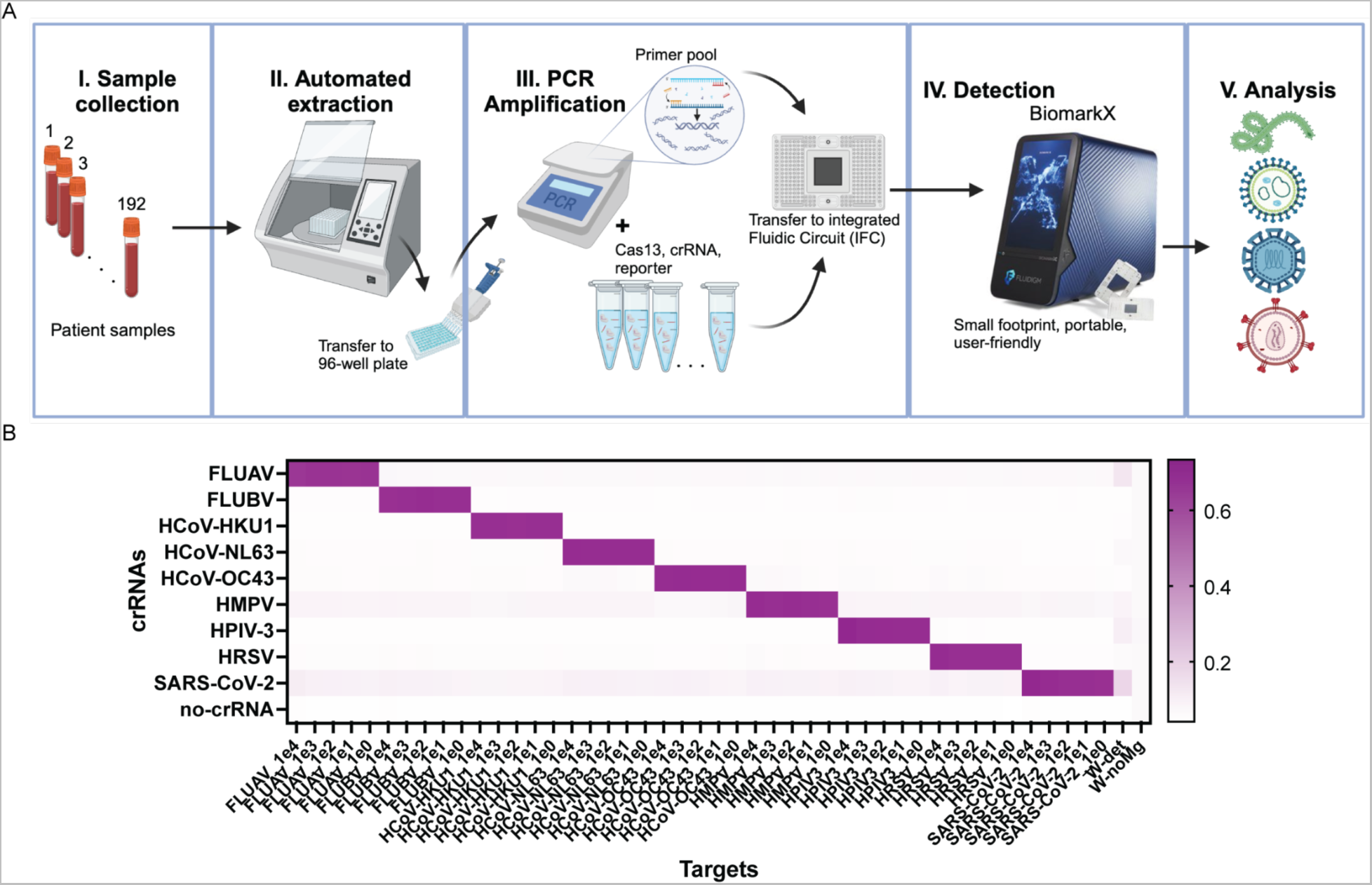
Robust detection of the respiratory virus panel using the CARMEN-BiomarkX. (**A**) Schematic of CARMEN workflow from blood samples to final results using the BiomarkX instrument from Standard BioTools. (**B**) Heatmaps illustrating CARMEN assay performance for the respiratory virus panel (RVP) including SARS-CoV-2, HCoV-HKU1, HCoV-OC43, HCoV-NL63, FLUAV, FLUBV, HPIV-3, HRSV, HMPV. Data shows fluorescence intensity of synthetic DNA fragments, at 10^4^-10^0^ copies per µL, and corresponding viral Cas13 crRNAs at 1 hour post-reaction initiation.

### Assay design of bloodborne pathogen panels (BBPs)

To create each BBP panel, we strategically selected clinically relevant microbes and grouped them based on their likelihood to co-infect or extent to which their disease manifestations resembled one another (**Table S1**). Notably, diseases such as Ebola, Marburg, and Lassa fever fall under the category of viral hemorrhagic fevers, which often present similar symptoms like headaches, abdominal pain, fever, nausea, and frequent progression to hemorrhagic bleeds^21^. Accordingly, we included these symptomatically similar pathogens in a single panel, BBP1, alongside other fever-causing viruses (**Table 1, S1**). Similarly, we developed panels BBP2 and BBP3 to target mosquito-borne viral diseases and sexually transmitted illnesses (STIs), respectively.

**Table 1.** BBP organisms and percent genomes covered by the respective CRISPR assays. . Table lists the virus names included in this study, categorized into three panels: Panel 1 (BBP1), Panel 2 (BBP2), and Panel 3 (BBP3). For each organism, the percent of genomes covered by their assay designs compared to available complete genomes on NCBI is listed. The analysis was performed as described by Metsky *et. al.* using the script on Github.

| No. | Panel | Targets | Percent of genomes covered |
| --- | --- | --- | --- |
| 1 | BBP1 | Crimean Congo Hemorrhagic fever Virus (CCHFV) | 90 |
| 2 |  | Ebola virus (EBOV) | 100 |
| 3 |  | Lassa Fever (LASV) | 100 |
| 4 |  | Human Immunodeficiency Virus 1 (HIV1) | 100 |
| 5 |  | Human Immunodeficiency Virus 2 (HIV2) | 100 |
| 6 |  | Marburg Virus (MBV) | 100 |
| 7 |  | West Nile Fever Virus (WNV) | 99 |
| 8 |  | Yellow Fever Virus (YFV) | 92 |
| 9 | BBP2 | Chikungunya (CHI) | 98 |
| 10 |  | Dengue Virus (4 targets, DENV) | 99.9 |
| 11 |  | Hantaan orthohantavirus (HTV) | 100 |
| 12 |  | Measles morbillivirus (MMV) | 100 |
| 13 |  | Nipah henipavirus (NPV) | 100 |
| 14 |  | Rabies lyssavirus (RBV) | 92 |
| 15 |  | Rift Valley Fever Virus (RVFV) | 100 |
| 16 |  | Zika Virus (Zika) | 85 |
| 17 |  | Hepatitis C Virus (HCV) | 100 |
| 18 | BBP3 | Mpox virus (MKP) | 95.9 |
| 19 |  | Hepatitis B Virus (HBV) | 99.9 |
| 20 |  | Herpes Simplex Virus Type 2 (HSV-2) | 98.7 |
| 21 |  | Chlamydia trachomatis (Chla) | 99.8 |
| 22 |  | Treponema pallidum subsp. pallidum (Syph) | 100 |
| 23 |  | Trichomonas vaginalis (Trich) | 98.7 |

Ultimately, we designed assays to detect 23 pathogens across three distinct panels, including 9 targets for viral hemorrhagic fevers (BBP1), 10 targets for mosquito-borne viral diseases (BBP2) and 7 targets for STIs (BBP3). BBP1 contained (1) *Crimean congo hemorrhagic fever virus* (CCHFV), (2) *Zaire ebolavirus* (EBOV), (3) *Human immunodeficiency virus 1* (HIV1), (4) *Human immunodeficiency virus 2* (HIV2), (5) *Lassa virus* (LASV), (6) *Marburg virus* (MBV), (7) *West-nile virus* (WNV), (8) *Yellow fever virus* (YFV). BBP2 contained (9) *Chikungunya virus* (CHI), (10) *Dengue virus* (DENV), (11) *Hantaan orthohantavirus* (HTV), (12) *Measles morbillivirus* (MMV), (13) *Rabies lyssavirus* (RBV), (14) *Rift Valley Fever virus* (RVFV), (15) *Zika virus* (Zika), (16) *Nipah henipavirus* (NPV). BBP3 contained (17) *mpox virus* (MKP, formerly *monkeypox virus*), (18) *Hepatitis B virus* (HBV), (19) *Hepatitis C virus* (HCV), (20) *Herpes Simplex Virus Type 2* (HSV2), (21) *Chlamydia trachomatis* (Chla), (22) *Treponema pallidum subsp. pallidum* (Syph), and (23) *Trichomonas vaginalis* (Trich) (**Table S2**).

For each target, we used ADAPT to design five microbe-specific crRNA assays and PCR primer pairs, totaling 115 assay designs^19^. This involved collecting complete genomes from NCBI and utilizing FASTA files to generate compatible crRNAs and primer sequences, as predicted by the ADAPT program. While most of ADAPT’s designs included a single crRNA guide, LASV and DENV required multiple crRNA designs to maximize coverage across all analyzed genomes and ensure efficient capture of genomic diversity^16,22^. Indeed, this multi-guide system is predicted to capture LASV-Sierra Leone and LASV-Nigeria clades, as well as all four DENV serotypes (**Figure S2-S3**). Our sequence analysis demonstrated that ADAPT-designed assays represented between 85 and 100% of the complete genomes collected from NCBI for all but four of the pathogens, suggesting their effectiveness in detecting the intended target sequences (**Table 1**).

### BBP CARMEN development and testing using synthetic material

Using synthetic DNA targets at a concentration of 10^8^ cp/µL, we conducted CARMEN experiments to identify which crRNA assay yielded the highest fluorescent signals over background ratio and lowest LOD values (**Figure S4-S6**). This combination would ensure specific and sensitive sequence detection and the best-performing assays were selected for downstream analyses. After downselection, we integrated these optimized assays into their respective panels and evaluated their collective performance for multiplexed detection of BBP1, BBP2, and BBP3 pathogens (**Figures 2A-C**). In line with previous CARMEN studies, our results demonstrated no off-target signals, indicating compatibility and suitability for the multiplexed detection of BBPs.

**Figure 2.**
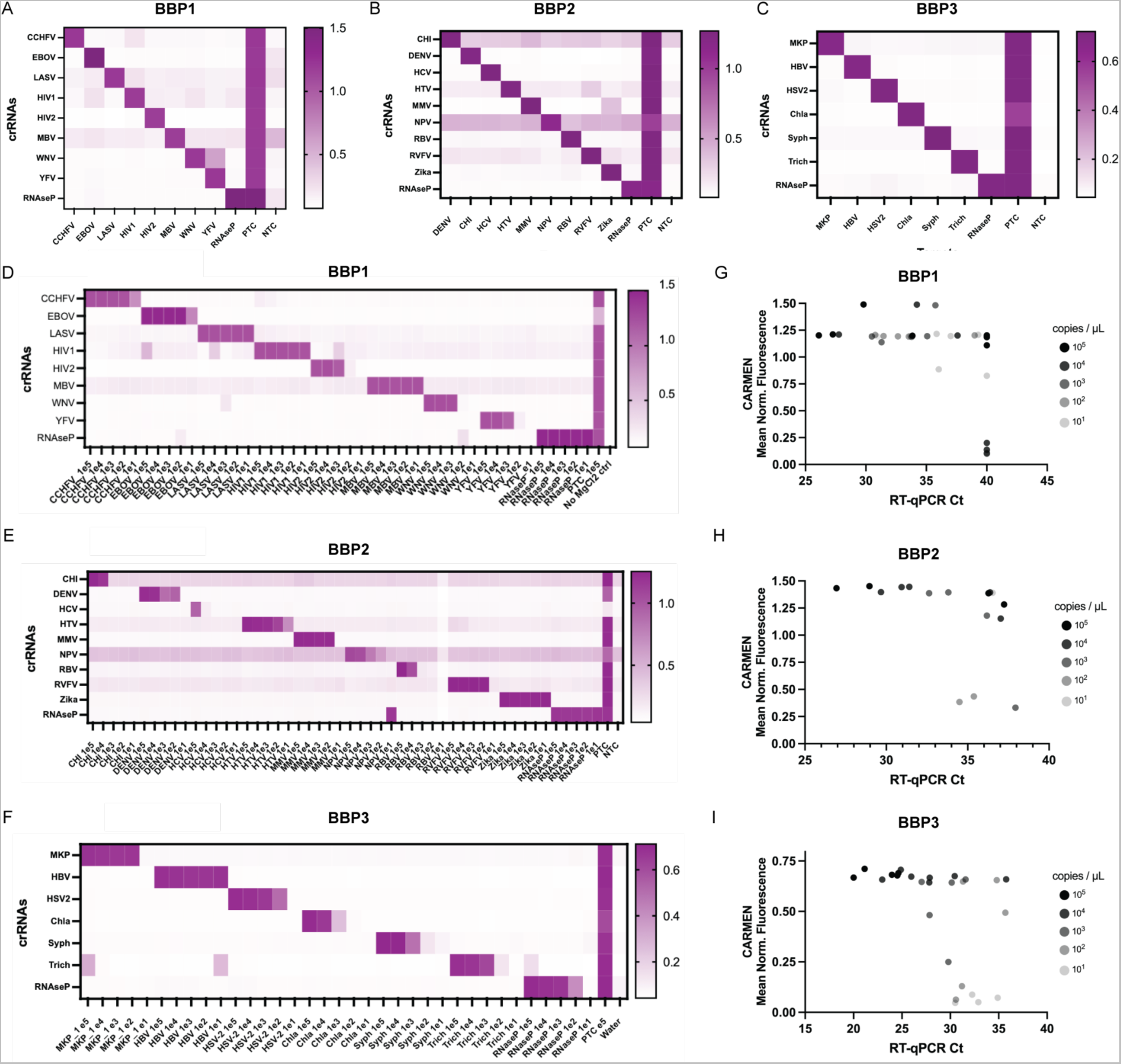
Sensitivity and specificity assessment of CARMEN assays for Bloodborne Pathogen Panels (BBPs) synthetic detection. (**A-C**) Heatmaps illustrating CARMEN assay performance for (**A**) BBP1, (**B**) BBP2, and (**C**) BBP3 panels, showing fluorescence intensity of synthetic DNA fragments, at 10^8^ cp/µL, and corresponding viral Cas13 crRNAs at 1 hour post-reaction initiation. (**D-F**) Heatmaps showing fluorescence values of the targets at the indicated concentrations (ranging from 10^5^ to 10^1^ cp/µL) for (**D**) BBP1, (**E**) BPP2, and (**F**) BBP3. Normalized fluorescence signal at 1 h post-reaction initiation of corresponding viral Cas13 crRNAs is shown. All samples were background-subtracted from the no target control (NTC). (**G-I**) Comparison analysis of CARMEN fluorescence to RT-qPCR Ct values for (**G**) BBP1, (**H**) BBP2, and (**I**) BBP3.

We then sought to characterize the LOD values for each of the 23 targets in the BBP panels. We prepared dilutions of synthetic materials ranging from 10^5^ cp/µL down to 10^1^ cp/µL for amplification, followed by CARMEN fluorescence detection (**Figures 2D-F**). We observed variable LODs across the 23 targets. Notably, for BBP1 and BBP3, all targets exhibited LODs of 10^3^-10^1^ cp/µL (**Figures 2D, 2F**). Many targets, such as EBOV, Lassa, and Zika, demonstrated significant fluorescence even at the lowest dilution tested (10 cp/µL), indicating highly sensitive detection. In contrast, BBP2 exhibited higher LODs (9 for BBP2 compared to 8 and 6 for BBP1 and BBP3, respectively), which has been associated with lower assay performance^17^. This difference is likely due to the higher number of pooled assays in BBP2 compared to the other two panels. Nevertheless, with the exception of CHI, HCV, and RBV, the remaining targets in BBP2 still demonstrated LODs of 10^3^-10^1^ cp/µL. These results demonstrate sensitive multiplexed detection of synthetic BBP samples with minimal off-target signals, capable of detecting target sequences.

We compared the performance of CARMEN fluorescence LODs for individual assays within our multiplexed panels against an RT-qPCR gold-standard assay for singular targets. Using the same synthetic materials at dilutions ranging from 10^5^ to 10^1^ cp/µL, we observed a general trend of higher LODs (i.e. lower sensitivity) for qPCR when compared with corresponding CARMEN results (**Figures 2G-I, S7**). Specifically, we observed sensitive fluorescent detection at concentrations of 10 cp/µL for BBP1 despite yielding Ct values of 35 or higher (**Figure 2G**, **S7A**). While BBP2 exhibited lower performance for the NPV assay, resulting in higher CARMEN LODs compared to RT-qPCR LODs (**Figure S7B**), the remaining assays demonstrated increased sensitivity using CARMEN compared to RT-qPCR analysis (**Figure 2H**). For BBP3, we observed slightly higher CARMEN LODs––particularly for Chla and Syph––which may be attributable to the relatively higher GC content of bacterial species (**Figure 2I, S7C**). Nonetheless, on average, CARMEN detection outperformed RT-qPCR analysis (**Figure S7D)**.

Thus, comparison with RT-qPCR analysis consistently showed CARMEN’s superior performance, highlighting its potential for multiplexed BBP detection.

### Specificity and sensitivity analysis of BBP detection using Healthy Normal Serum samples

We evaluated the specificity of BBP detection using clinical samples obtained from healthy individuals. We purchased twenty (20) serum samples labeled as Healthy Normal Serum (HNS) from Boca Biolistics, each containing one milliliter of serum. Following standard extraction and RT-PCR protocols, we assessed whether BBP CARMEN detection resulted in background fluorescence using these samples (**Figure S8**). After thresholding against the NTC, with the exception of the Trich assay, we found no false positive results across all three BBP panels for all 20 HNS samples, consistent with RT-qPCR data (**Figure S8A-C**). Thus, Trich was eliminated from BBP3 for downstream analyses (**Figure S8D**).

Subsequently, we investigated whether the LODs determined using synthetic samples were affected by the extraction step upstream of RT-qPCR and detection analysis. We utilized pooled HNS samples, subsequently aliquoting and spiking them with known concentrations of viral genomic DNA or RNA (**Figure 3, S9-S10**). We first performed initial optimization analysis with synthetic LASV and identified 50 nM concentration as the optimal RNAse P primer concentration (**Figure S9**). Next, we spiked LASV and MPOX viral genomic material (RNA for LASV and DNA for MPOX) at final concentrations of 10^4^ to 1 cp/μL and 10^3^ to 1, respectively, into each HNS aliquot, followed by extraction of nucleic acid materials using the Zymo Research Quick-DNA/RNA MagBead™ extraction kit (R2130/R2131) on the KingFisher Flex instrument. We then used extracted material as input into the Aldatu Biosciences PANDAA LASV RT-qPCR detection kit (2011096) and standard CARMEN analysis.

**Figure 3.**
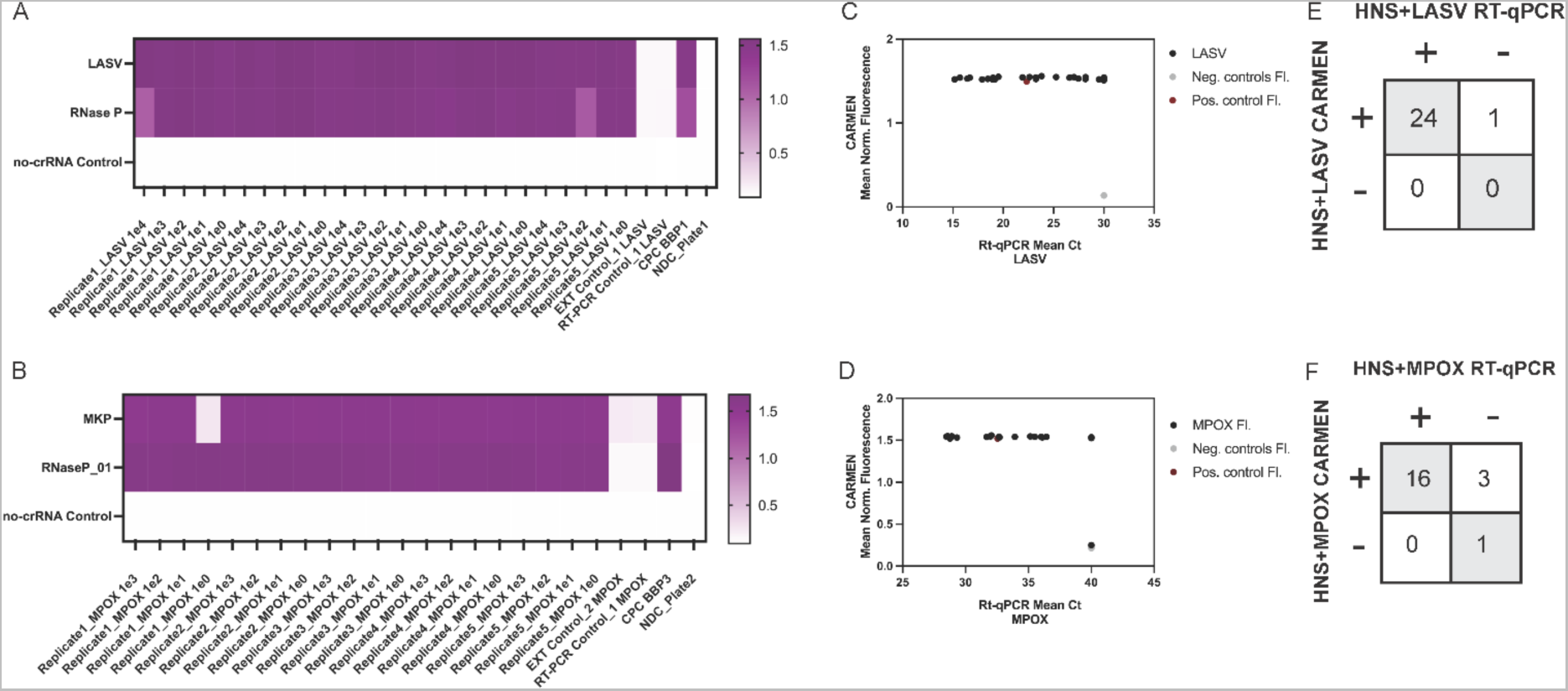
Sensitivity and specificity analysis with contrived human normal samples. (**A-B**) Heatmaps showing fluorescence values of (**A**) LASV or (**B**) MPOX RNA extracts spiked into HNS at the indicated concentrations (ranging from 10^3^ to 10^0^ cp/µL) followed by CARMEN detection with BBP1 and BBP3 respectively. Normalized fluorescence signal at 1 h post-reaction initiation of corresponding viral Cas13 crRNAs is shown. (**C-D**) Comparison analysis of CARMEN fluorescence to RT-qPCR Ct values for contrived (**C**) LASV and (**D**) MPOX HNS samples. (**E-F**) Concordance table of CARMEN fluorescence to RT-qPCR Ct values for contrived (**E**) LASV and (**F**) MPOX HNS samples.

Our results demonstrated specific and sensitive detection of BBP targets, with an improved sensitivity compared to RT-qPCR, and high concordance between the two assays. We found 100% of LASV and 95% of MPOX-contrived HNS samples positive using CARMEN, including samples at concentrations of 1 cp/μL (**Figure 3A-B**). Under these conditions, the LOD was determined to be as low as 1 cp/μL, with notable effects of the extraction step on detection sensitivity. We observed a robust CARMEN signal, albeit with a single WNV contaminant, even at the lowest concentration tested (**Figure S10A**). Using these same samples, we found 96% of LASV and 80% of MPOX contrived HNS samples positive using RT-qPCR (**Figure 3C-D, Figure S10B-C**). By comparing these results with CARMEN, we found 100% and 96% concordance between the two assays for LASV and MPOX, respectively (**Figure 3E-F**).

These findings underscore the reliability of BBP CARMEN as a robust assay for the accurate and sensitive detection of microbial pathogens in human samples.

### Validation of BBP CARMEN assays using patient samples

We assessed whether our BBP-CARMEN platform could successfully detect pathogens from confirmed-positive patient samples in hospital and field settings (**Figure 4**). We partnered with the Massachusetts General Hospital (MGH) Sexual Health Clinic and obtained 10 patient samples with confirmed-positive diagnoses for *N. gonorrhoeae* (Gon), as well as 38 Gon negative samples, by standard clinical diagnostic tests. Of these 38 negative samples, three were also annotated as *Chlamydia trachomatis* (Chla) positive (Chla is included in BBP3). We had previously shown that this CRISPR-Cas13 assay performed well on clinical samples in a single assay design^23,24^. We conducted CARMEN detection on these patient samples and analyzed the data using the above-mentioned threshold calculations.

**Figure 4.**
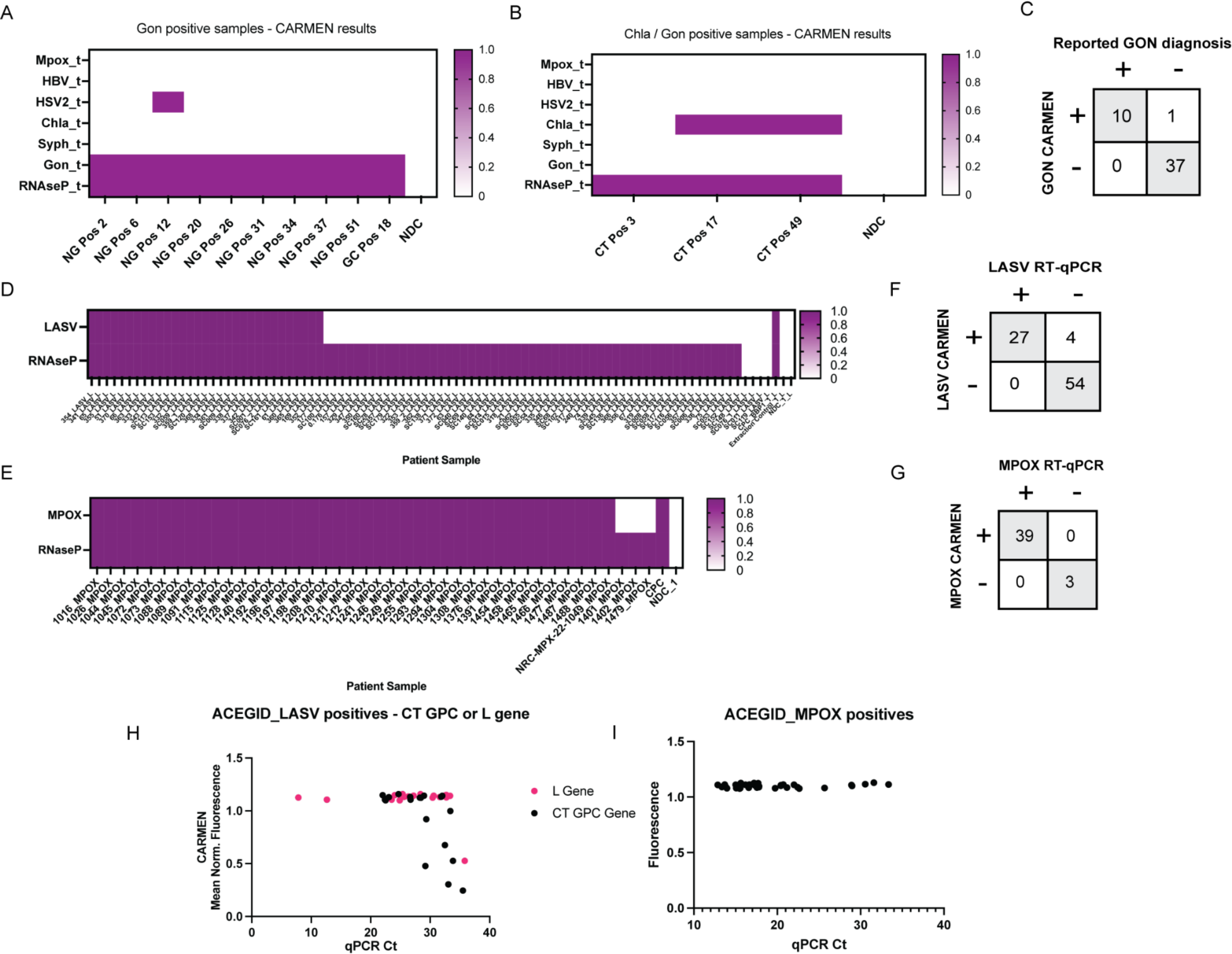
Sensitive, accurate, detection of Gonorrhea, Chlamydia, Lassa virus, and Monkeypox in patient samples. (**A-B**) Heatmaps illustrating CARMEN detection of (**A**) *Neisseria gonorrhoeae*, and (**B**) *Chlamydia trachomatis*. (**C**) Comparison analysis of CARMEN fluorescence to RT-qPCR Ct values for Gon patient samples. (**D-E**) Heatmaps showing fluorescence values of (**D**) LASV or (**E**) MPOX clinically positive patient samples. (**F-G**) Concordance table of CARMEN fluorescence to RT-qPCR Ct values for (**F**) LASV and (**G**) MPOX patient sample. (**H-I**) Comparison analysis of CARMEN fluorescence to RT-qPCR Ct values for (**H**) LASV, and (**I**) MPOX. For all heatmaps, normalized fluorescence signal at 1 h post-reaction initiation of corresponding viral Cas13 crRNAs is shown.

All ten confirmed-positive samples were found positive for Gon using BBP3-CARMEN (**Figure 4A**) yielding 100 % positive concordance. Serendipitously, one confirmed-positive sample was also found positive for HSV2 (included in the same BBP3 panel). Interestingly, one confirmed-negative Gon sample was found positive by CARMEN, and the remaining 37 Gon negative samples were found negative for Gon, yielding a 97 % negative concordance rate (**Figure S11**). Notably, of the three Chla positive samples, two were found positive for Chla using CARMEN (**Figure 4B**). These results demonstrated 100% concordance between BBP3-CARMEN and diagnostic test results from the MGH Sexual Health Clinic (**Figure 4C**).

Finally, we validated our BBP CARMEN assays in Nigeria using patient samples with confirmed-positive diagnoses of LASV and MPOX. In total, we extracted 90 LASV and 42 MPOX-positive samples previously identified at the hospital. Of note, the extended storage duration of the samples raised the potential for sample degradation. We thus amplified and analyzed the samples following standardized operating procedures and optimizations based on contrived sample testing using BBP CARMEN alongside simultaneous RT-qPCR testing (see **Supplementary Information** for specific SOP details).

The CARMEN LASV and MPOX outperformed and matched RT-qPCR sensitivity, respectively. We found 31 LASV (34%) and 39 MPOX (93%) samples were positive using CARMEN (**Figure 4D-E**), and 27 LASV (30%) and 39 MPOX (93%) samples positive using RT-qPCR (**Figure 4F-G**). No false-negative results were observed for LASV samples, suggesting the possibility that sample integrity may have been compromised during storage or retrieval (**Figure 4H**). Notably, 4 LASV samples tested positive by CARMEN but negative by RT-qPCR, indicating the increased sensitivity of the CARMEN assay compared to RT-qPCR. This result corroborated our findings with synthetic and contrived samples, and previous findings on clinical samples^20^. The MPOX samples demonstrated 100% concordance between the two assays, with 39 positive and 3 negative outcomes (**Figure 4I**).

## Discussion

The development of bloodborne pathogen panels (BBPs) for CARMEN detection represents a crucial step toward addressing the diagnostic challenges posed by illnesses sharing similar clinical symptoms. By strategically selecting microbes with overlapping symptoms, we designed three panels broadly targeting: viral hemorrhagic fevers, mosquito-borne viral diseases, and sexually transmitted illnesses. The comprehensive design of 23 CRISPR-Cas13 assays using the ADAPT program ensured broad coverage of target sequences across diverse genomes, reflecting the effectiveness of our assay design strategy.

Subsequently, using synthetic materials, we characterized the limit of detection (LOD) for each target in the BBP panels. Our results demonstrated sensitive detection of BBP targets, with many exhibiting LODs of 1,000 copies per microliter (cp/µL) or less. Notably, CARMEN fluorescence LODs outperformed RT-qPCR Ct analysis in terms of sensitivity, highlighting the potential of CARMEN for sensitive multiplexed BBP detection. However, variations in LODs across targets and panels underscore the importance of assay optimization and validation for each specific application. Furthermore, specificity analysis using healthy normal serum (HNS) samples confirmed the absence of false-positive results across all BBP panels, validating the specificity of our assays. The compatibility of CARMEN with patient samples was further validated through analyzing contrived HNS samples spiked with known concentrations of viral seedstocks, which demonstrated specific and sensitive detection of BBP targets even in complex sample matrices.

Lastly, validation using patient samples with confirmed diagnoses of gonorrhea (Gon), lassa (LASV), and mpox (MPOX) infections highlighted the robustness of the BBP CARMEN assays in real-world clinical settings. The discrepancy observed between original testing and current RT-PCR results for LASV samples highlights issues with sample integrity during long-term storage. However the high concordance observed between CARMEN and RT-PCR reinforces, and greater positive detections using the CARMEN assay, highlights the reliability of CARMEN - with increased sensitivity - for bloodborne pathogen detection. The high pathogenicity of BBP targets presents a substantial obstacle in procuring suitable seedstocks and positive patient samples essential for panel validation. Future research aims to address this challenge through the establishment of international partnerships and efforts to create sample repositories.

The optimization of the CARMEN workflow using Fluidigm’s BiomarkX also represents a significant advancement toward enabling scalable, high-throughput, and multiplexed analysis of patient samples in standard clinical laboratories. By transitioning from the BiomarkHD to the BiomarkX instrument, we aimed to streamline the workflow and reduce the turnaround time of diagnostic testing. Indeed, the BiomarkX offers several advantages over its predecessor, including a smaller footprint, user-friendly operation, and compatibility with various integrated fluidic circuits (IFCs). Our evaluation of the BiomarkX instrument for detecting synthetic DNA from respiratory viruses demonstrated robust performance, validating the suitability of the BiomarkX for CARMEN analysis. More broadly, the ease of transition of our CRISPR-based assays to these new platforms points to the robustness of these assays, and the opportunities to integrate them into a range of technological platforms.

Ultimately, our study demonstrates the successful optimization and validation of CARMEN assays for sensitive and specific detection of bloodborne pathogens, laying the foundation for their widespread application in clinical diagnostics and public health surveillance. Future research efforts should focus on further refining assay performance, expanding the range of detectable pathogens, and evaluating the clinical utility of CARMEN in diverse settings. Ultimately, the adoption of CARMEN technology has the potential to transform pathogen detection and facilitate early intervention strategies for infectious disease control and management.

## Methods

Sample size was not predetermined using statistical methods. The experiments lacked randomization, and the investigators were not blinded to allocation during both experiments and outcome assessment.

### MGH Patient Samples statement

Stored clinical urine specimens were previously collected between March and September 2023 as a part of an ongoing clinical trial in the Massachusetts General Hospital (MGH) Sexual Health Clinic (NCT05564299). Participants were selected using the following criteria: 18 years or older, presented with symptoms of urethritis or cervicitis (urethral or virginal discharge, dysuria, or dyspareunia), not known to be pregnant at the time of enrollment, no known exposure to *N. gonorrhoeae* or *Chlamydia trachomatis* within the previous six weeks, no concurrent symptoms at extragenital sites, and were willing to provide informed consent. 1.5 mL of each urine sample were aliquoted into cryotube vials, assigned a unique study identifier, and stored at −80° C within 30 minutes of sample aliquoting. As a part of routine care, confirmatory diagnostic testing included gram stain, nucleic acid amplification testing, and culture with susceptibility testing using standard methods. These specimens were deemed to be *N. gonorrhoeae* positive if standard clinical diagnostics showed positive results.

### ACEGID Patient samples statement

Patients were selected based on PCR outcome: confirmed Positive, with Ct values ranging from 15-40. Plasma and swab samples, for LASV and MPOX respectively, were collected in EDTA tubes for further processing. Samples were stored at −80° C and at −196° C liquid nitrogen for up to one year in plasma AVL for LASV, and Viral Transport Media for MPOX. Prior to CARMEN analysis, samples were processed by inactivation with AVL and extraction via Qiagen protocol.

#### 1. General procedures

A detailed standard operating procedure for running the BBP panels on CARMEN can be found in the Supplementary Information.

##### 1. a. Production and processing of synthetic materials

Synthetic materials were synthesized and handled as described before. Briefly, crRNAs (Integrated DNA Technologies) were resuspended in nuclease-free water to 100 μM and then diluted for input into the detection reaction. Primer sequences (Eton or Integrated DNA Technologies) were also resuspended to 100 μM in nuclease-free water and combined at specified concentrations for pooled amplification.

##### 1. b. *In-vitro* transcription (IVT) of DNA targets

DNA targets were *in-vitro* transcribed to use as RNA standards for experiments as described previously. Briefly, DNA targets were procured from Integrated DNA Technologies and subjected to *in-vitro* transcription (IVT) using the HiScribe T7 High Yield RNA Synthesis Kit (New England Biolabs). Transcriptions adhered to the manufacturer’s recommendations, with a 20 μL reaction volume (30 µL if the target of interest was above 300 base pairs in length), incubated overnight at 37 °C and followed by DNAse I (New England BioLabs) treatment to remove the DNA template. Purification of transcribed RNA products employed RNAClean XP beads (Beckman Coulter), and quantification was carried out using the Invitrogen™ Qubit™ RNA High Sensitivity (HS) kit as recommended by the manufacturer. For experimentation purposes, RNA was serially diluted from 10^8^ down to 10^1^ cp/µL and utilized as input for the subsequent amplification reaction.

##### 1. c. Nucleic acid extraction and processing

Samples underwent automated total nucleic acid extraction utilizing the Zymo Research Quick- DNA/RNA™ MagBead kit on the KingFisher Flex 96 Deep-well Head Magnetic Particle Processor (Thermo Fisher Scientific). Following the manufacturer’s instructions, samples were pre-processed based on their sample matrix type as outlined in “Sample Preparation” in the Quick-DNA/RNA™ MagBead protocol provided by Zymo Research (pages 6 – 9, catalog nos. R2130/R2131). 200 µl of sample material produced from pre-processing was inputted into the extraction pipeline as outlined in “Automation Reference Guide – KingFisher Flex” in the Quick-DNA/RNA™ Magbead – Co-Purification protocol provided by Zymo Research (pages 3 – 4, catalog nos. R2130/R2131). Sample material was then placed on the KingFisher Flex instrument alongside required reagents and the KingFisher Flex protocol “R2130_Quick DNARNA Magbead_KingFisherFlex_Copurification_v2.bdz” was run (provided by Zymo Research). RNA and/or DNA from inputted sample material was eluted in 50 µl of nuclease-free water and either added as input into the RT-PCR amplification step or stored at −80°C until usage.

##### 1. d. QIAGEN OneStep RT-PCR amplification

Reverse-transcription PCR amplification was performed as described previously. Briefly, a total reaction volume of 50 μl was used: 12.5 µl QIAGEN OneStep RT–PCR 5x buffer, 3 µl each of pooled target forward and reverse primers, 2 µl of QIAGEN enzyme mix, 2 µl of QIAGEN dNTP mix, 17.5 µl nuclease-free water, and a 10 µl RNA input. The final concentrations for target primers and RNase P primers were set at 300 nM and 50 nM, respectively. Thermal cycling conditions comprised reverse transcription at 50 °C for 30 min, initial PCR activation at 95 °C for 15 minutes, followed by 40 cycles at 94 °C for 30 s, 58 °C for 30 s, and 72 °C for 30 s. Reactions were stopped by dropping the temperature to 4 °C. Please refer to <u>Supplementary Information</u> - Table S1 for details on the primer sequences used in each BBP panel.

##### 1. e. Standard BioTools BiomarkX detection

Fluidigm detection was performed as described previously with major updates on data collection and processing with the newer Fluidigm model, the BiomarkX. The Cas13 detection reactions were divided into two distinct mixes—namely, the assay mix and the sample mix (details can be found below). These mixes were loaded onto a 192.24 microfluidic Integrated Fluidic Circuit (IFC) for analysis with BiomarkX or BiomarkHD (Fluidigm instruments).

###### Assay mix

The assay mix comprised LwaCas13a (GenScript) at 20nM final concentration. The assay also included 2X Assay Loading Reagent (Fluidigm), 69 U T7 RNA Polymerase mix (NEB), and crRNAs at 1 µM concentrations, resulting in a total volume of 16 μl per reaction.

###### Sample mix

The sample mix contained 1x homemade 10X cleavage buffer (1M Tris-HCl (pH 7.5), 0.1M dithiothreitol, and nuclease-free water), 25.2 U RNase Inhibitor (New England Biolabs), 0.1X ROX reference dye (Invitrogen), 20X loading reagent (Fluidigm), 1 mM rNTPs (New England Biolabs), 9 mM MgCl_2_ in water, and a 500 nM quenched synthetic fluorescent RNA reporter (FAM/rUrUrUrUrUrUrU/3IABkFQ/; Integrated DNA Technologies) was used for a total volume of 14 μl per reaction.

###### IFC loading and running on Fluidigm instruments

192.24 IFCs were prepared and loaded with samples as described previously. The IFCs were run on the BiomarkHD and the BiomarkX (all subsequent data) according to the manufacturer’s instructions. For the BiomarkHD, the IFC was loaded onto the IFC Controller RX (Fluidigm) where the ‘Load Mix’ script was run. For the BiomarkX, the IFC was loaded directly into the instrument. After proper IFC loading, images were collected over a 1-h period at 37 °C using a custom protocol pre-loaded into the Fluidigm’s instruments.

###### Fluidigm data analysis

Fluidigm data analysis was performed as described previously. Briefly, fluorescence analysis involved plotting reference-normalized and background-subtracted values for guide-target pairs. The reference-normalized value for a guide-target pair (at time point t and target concentration) was calculated as (median(P_t_ − P_0_) / (R_t_ − R_0_)), where P_t_ is the guide signal (FAM) at the time point, P_0_ is its background measurement, R_t_ is the reference signal (ROX) at time point *t*, R_0_ is its background measurement, and the median is taken across replicates. The same calculation was applied to the no-template control (NTC) of the guide, providing a background fluorescence value for the guide at time point *t*. The reference-normalized, background-subtracted fluorescence for a guide-target pair is the difference between these two values. A sample was considered positive if the signal produced was greater than the average signal produced by that assay’s NTC plus three times the standard deviation of the signal in the NTCs.

#### 2. Designs and development of bloodborne pathogen panels

##### 2.a. Designs

Oligonucleotide primers and crRNA guides were designed to detect conserved regions. Briefly, these conserved regions were from the following 23 pathogens: (1) *Crimean Congo Hemorrhagic fever Virus* (CCHFV), (2) *Zaire ebolavirus* (EBOV), (3) *Human Immunodeficiency Virus 2* (HIV2), (4) *Human Immunodeficiency Virus 1* (HIV1), (5) *Lassa virus* (LASV), (6) *Marburg virus*, (7) *West-Nile Virus* (WNV), (8) *Yellow Fever virus* (YFV), (9) *Chikungunya virus* (CHI), (10) *Dengue virus* (DENV), (11) *Hantaan orthohantavirus* (HTV), (12) *Measles morbillivirus* (MMV), (13) *Rabies lyssavirus* (RBV), (14) *Rift Valley Fever virus* (RVFV), (15) *Zika virus* (Zika), (16) *Nipah henipavirus* (NPV), (17) *mpox virus* (MKP, formerly *Monkeypox virus*), (18) *Hepatitis B virus* (HBV), (19) *Hepatitis C virus* (HCV), (20) *Herpes Simplex Virus Type 2* (HSV2), (21) *Chlamydia trachomatis* (Chla), (22) *Treponema pallidum subsp. pallidum* (Syph), and (23) *Trichomonas vaginalis* (Trich). If available, we leveraged pre-designed assays as published on the Activity-informed Design with All-inclusive Patrolling of Targets (ADAPT; https://adapt.run/) program. Otherwise, complete genomes were collected from National Center for Biotechnology Information (NCBI), aligned, and fed into ADAPT for crRNA design with >90% coverage. Highly-conserved regions were selected, and primers were manually designed using NCBI’s Primer-BLAST for optimal amplification target regions with crRNA binding regions in the middle. For our positive controls, we used RNase P primers and crRNA sequences as described previously. All sequences used in this study can be found in Supplementary Information Table S1.

##### 2.b. *In-vitro* analysis and limit of detection testing

For each organism, five assays were designed and tested for specificity. The synthetic DNA targets contained the consensus sequence that was position-matched to the location of the BBP virus of interest targets in the viral genome. Samples were serially diluted down to a concentration of 10^6^ to 10^1^ cp/µL and were prepared for the specificity experiments according to the methods described above in Section 1.b. ‘*In-vitro* transcription (IVT) of DNA targets’.

Synthetic RNA of each BBP target ranging from 10^8^ to 10 cp/µL were analyzed on the CARMEN platform (QIAGEN amplification and Standard BioTools BioMarkX Detection) as well as by RT-qPCR as described in Section 2d. Utilizing the reference-normalized value of each guide-target pair, the threshold for each assay was calculated as (average(NTC) + 3(standard deviation(NTC)). Through this calculation each sample was thresholded as either negative or positive for each tested assay; whereas if a sample is above an assay threshold it is considered positive and if it is below then it is considered negative. The synthetic RNA of each target ranging from 10^8^ to 10 cp/µL was then evaluated against each guide specific threshold demonstrating the lowest concentration of detection per guide.

##### 2.c. Testing of contrived samples

Genomic RNA/DNA from BEI Resources and Boston University’s National Emerging Infectious Diseases Laboratories (NEIDL) were quantified using the Power SYBR Green RNA-to-CT 1-Step Kit (Thermo Fisher Scientific). Reactions, performed in triplicate with 1 μL RNA input in 10-μL reactions, utilized a singleplex primer mix at 500 nM (see Supplementary Information for sequences). Thermal cycling conditions included reverse transcription at 48 °C for 30 min, enzyme activation at 95 °C for 10 min, and 40 cycles at 95 °C for 15 sec and 58 °C for 1 min, followed by a melt curve step at 95 °C for 15 sec, 58 °C for sec, and 95 °C for sec. Standard curves were generated using spike-in RNA templates (generated as described in section 1b) in tenfold serial dilutions and analyzed on the QuantStudio 6 Flex Real-Time PCR System. Contrived samples were created as previously described. Briefly, 160 μL of pooled commercially available healthy normal blood serum samples (Boca Biolistics) were mixed with 40 μL of available nucleic acid extracts (BEI Resources and Boston University’s National Emerging Infectious Diseases Laboratories (NEIDL)) at known concentrations for a 200 μl total sample volume. Specifically, the following reagent was obtained through BEI Resources, NIAID, NIH: RNA from Lassa Virus, Josiah, NR-31821. Following documentation from the Zymo Research Quick-DNA/RNA™ MagBead kit, 200 μL of each contrived sample was utilized in the automated extraction on the ThermoFisher KingFisher Sample Purification System (details of extraction process found above). Following automated extraction, nucleic acid extracts were tested in parallel; both on the CARMEN platform and validated commercially available detection kits (Aldatu Biosciences PANDAA LASV RT-qPCR for Lassa virus and RayBiotech Mpox virus (MPXV) PCR Nucleic Acid Detection Kit for mpox). Following specifications outlined from the manufacturers of the commercially available detection kits, results were determined and compared to the CARMEN output.

#### 3. Patient specimen validation

Aiming to demonstrate the specificity and sensitivity of the CARMEN platform on confirmed-positive patient samples, clinical Gonorrhea, Lassa, and MPOX samples were tested. Gonorrhea samples were provided by the Massachusetts General Hospital. Lassa- and MPOX-positive samples were provided by the African Centre of Excellence for Genomics of Infectious Diseases (ACEGID). All samples were tested on the CARMEN platform and, with the exception of Gonorrhea, samples were validated with commercially available detection kits (RealStar Lassa virus RT-PCR Kit 2.0 for Lassa virus and RayBiotech Mpox Virus (MPXV) PCR Nucleic Acid Detection Kit for Monkeypox). Utilizing threshold parameters detailed in Section 2.e. ‘*In-vitro* analysis’, results for each sample were determined on the CARMEN platform. Dually, using specifications from the manufacturers of the RT-qPCR detection kits, results were determined and compared to the CARMEN output.

## Acknowledgements

We thank J. Arizti Sanz, A. Si, and H. Metsky for helping with guide design, sequence analysis, or sharing reagents. This work is made possible by support from Flu Lab and a cohort of generous donors through TED’s Audacious Project, including the ELMA Foundation, MacKenzie Scott, the Skoll Foundation, and Open Philanthropy. P.C.S. was supported by the Howard Hughes Medical Institute and Merck KGaA Future Insight Prize.

## Data availability

All requests for raw and analyzed data and materials will be reviewed by the Broad Institute of Harvard and MIT to verify if the request is subject to any intellectual property or confidentiality obligations. Data and materials that can be shared will be released via a material transfer agreement.

## Contributions

M.K. and P.C.S. initially conceived this study and then involved L.K., K.P. and E.S. M.K. and L.K. designed the primers and crRNAs for the pathogens presented in this study. M.K., L.K., and K.P. performed the initial experiments on the Fluidigm instrumentation. K.P. conducted the Lassa and Monkeypox contrived sample testing in an academic setting. G.S. and L.A-B. performed the clinical evaluation of the CARMEN BBPs at MGH under guidance from M.K., K.P. and J. L. K.P. and L.S. conducted the clinical evaluation of the CARMEN BBPs at ACEGID in Nigeria with assistance from P.E., A.M.I., A.S., O.O.O., A.O.A., C.l’A., I.B., M.F.P., C.W. and supervision from M.K., E.S., A.O., C.H., and P.C.S. M.K. generated the figures with help from K.P. and wrote the paper with help from K.P. and E.S., and guidance from P.C.S. All authors reviewed the manuscript.

