## Supplemental Information for "Multiplexed detection of febrile infections using CARMEN"

### Materials and Methods

#### Panel designs

The oligonucleotide primers and crRNA guides were designed as described before (cite). Briefly, oligonucleotide primers and crRNA guides were designed to detect the conserved regions of the following 23 pathogens: (1) *Crimean Congo Hemorrhagic fever Virus* (CCHFV), (2) *Zaire ebolavirus* (EBOV), (3) *Human Immunodeficiency Virus 2* (HIV2), (4) *Human Immunodeficiency Virus 1* (HIV1), (5) *Lassa virus* (LASV), (6) *Marburg virus*, (7) *West-Nile Virus* (WNV), (8) *Yellow Fever virus* (YFV), (9) *Chikungunya virus* (CHI), (10) *Dengue virus* (DENV), (11) *Hantaan orthohantavirus* (HTV), (12) *Measles morbillivirus* (MMV), (13) *Rabies lyssavirus* (RBV), (14) *Rift Valley Fever virus* (RVFV), (15) *Zika virus* (Zika), (16) *Nipah henipavirus* (NPV), (17) *Monkeypox virus* (MKP), (18) *Hepatitis B virus* (HBV), (19) *Hepatitis C virus* (HCV), (20) *Herpes Simplex Virus Type 2* (HSV2), (21) *Chlamydia trachomatis* (Chla), (22) *Treponema pallidum subsp. pallidum* (Syph), and (23) *Trichomonas vaginalis* (Trich). If available, we leveraged pre-designed assays as published on the Activity-informed Design with All-inclusive Patrolling of Targets (ADAPT; https://adapt.run/) program (cite). For a select few not pre-designed on ADAPT, we designed the primers and crRNAs as follows: for each organism, FASTA files containing complete genomes were retrieved from the National Center for Biotechnology Information (NCBI), aligned, and manually uploaded into ADAPT to select highly conserved a region with the crRNA binding within. The software returned CRISPR-Cas13 assay designs, including target of interest and corresponding primers and crRNA guide sequences. In rare cases, multiple primers were designed to maximize full genomic diversity coverage. Primers were optimized for PCR using NCBI's Primer-BLAST Tool and T7 promoter sequence was added to each target and forward primer sequence to enable *in vitro* transcription. We used RNase P primers and crRNA sequences as described previously (cite). Primer pools were generated for each of the three panels (including RNAseP primers) and these pools were used for downstream analyses. All sequences used in this study can be found in Supplementary Information Table S2.

### Supporting Schemes and Tables

| No. | Panel 1 target (BBP1) | Panel 2 target (BBP2) | Panel 3 (BBP3) |
|---|---|---|---|
| 1 | Crimean Congo Hemorrhagic fever Virus (CCHFV | Chikungunya (CHI) | Mpox virus (MKP) |
| 2 | Ebola virus (EBOV) | Dengue Virus (4 targets, DENV) | Hepatitis B Virus (HBV) |
| 3 | Lassa Fever (LASV) | Hantaan orthohantavirus (HTV) | Herpes Simplex Virus Type 2 (HSV-2) |
| 4 | Human Immunodeficiency Virus 1 (HIV1) | Measles morbillivirus (MMV) | Chlamydia trachomatis (Chla) |
| 5 | Human Immunodeficiency Virus 2 (HIV2) | Nipah henipavirus (NPV) | Treponema pallidum subsp. pallidum (Syph) |
| 6 | Marburg Virus (MBV) | Rabies lyssavirus (RBV) | Trichomonas vaginalis (Trich) |
| 7 | West Nile Fever Virus (WNV) | Rift Valley Fever Virus (RVFV) | |
| 8 | Yellow Fever Virus (YFV) | Zika Virus (Zika) | |
| 9 | | Hepatitis C Virus (HCV) | |

**Table S1. Composition of bloodborne pathogen panels (BBPs)**. BBP1 contains: *Crimean Congo Hemorrhagic fever Virus* (CCHFV), *Zaire ebolavirus* (EBOV), *Human Immunodeficiency Virus 2* (HIV2), *Human Immunodeficiency Virus 1* (HIV1), *Lassa virus* (LASV), *Marburg virus*, *West-Nile Virus* (WNV), *Yellow Fever virus* (YFV). BBP2 contains: *Chikungunya virus* (CHI), *Dengue virus* (DENV), *Hantaan orthohantavirus* (HTV), *Measles morbillivirus* (MMV), *Rabies lyssavirus* (RBV), *Rift Valley Fever virus* (RVFV), *Zika virus* (Zika), *Nipah henipavirus* (NPV). BBP3 contains: *Mpox virus* (MKP, formerly *Monkeypox virus*), *Hepatitis B virus* (HBV), *Hepatitis C virus* (HCV), *Herpes Simplex Virus Type 2* (HSV2), *Chlamydia trachomatis* (Chla), *Treponema pallidum subsp. pallidum* (Syph), and *Trichomonas vaginalis* (Trich).

| No. | Pathogen Name | Mode of infection | Organism Family | Symptoms |
|---|---|---|---|---|
| 1 | Crimean Congo Hemorrhagic fever Virus (CCHFV) | Blood | Bunyavirirdae | Fever, hemorrage, headache/muscle ache, loss of appetite, sensitivity to light, nausea/vomitting |
| 2 | Ebola virus (EBOV) | Blood | Filoviridae | Fever, hemorrage, headache/muscle ache, malaise, sore throat, loss of appetite, nausea/vomitting, diarrhea, abominal pain |
| 3 | Lassa Fever (LASV) | Blood | Arenaviridae | Fever, hemorrage, headache/muscle ache, malaise, sore throat, chest pain, nausea/vomitting, diarrhea, abominal pain |
| 4 | Human Immunodeficiency Virus 1 (HIV1) | Blood, fluids | Retroviridae | Fever, muscle aches, malaise, swollen lymph nodes, chills, sore throat |
| 5 | Human Immunodeficiency Virus 2 (HIV2) | Blood, fluids | Retroviridae | Fever, muscle aches, malaise, swollen lymph nodes, chills, sore throat |
| 6 | Marburg Virus (MBV) | Blood | Filoviridae | Fever, hemorrage, sore throat, chest pain, nausea/vomitting, diarrhea, abominal pain, jaundice, inflammation of the pancreas, delirum, liver failure |
| 7 | West Nile Fever Virus (WNV) | Blood | Flaviviridae | Fever, muscle aches, abnominal pain, joint pain, vomiting, diarrhea, rash, swollen lymph nodes, loss of appetite |
| 8 | Yellow Fever Virus (YFV) | Blood | Flaviviridae | Fever, juandice, headache/muscle ache, nausea/vomitting, reddening of face/eyes/tongue, delirum, heart arrhythmias |
| 9 | Chikungunya (CHI) | Blood | Alphavirus | Fever, joint pain, rash, headache/muscle ache, joint swelling |
| 10 | Dengue Virus (4 targets, DENV) | Blood | Flaviviridae | Fever, vomitting, joint pain, rash, headache/muscle ache, severe-internal bleeding |
| 11 | Sin Nombre orthohantavirus (HTV) | Aerosol | Hantaviridae | Fever, fatigue, headache/muscle ache, nausea/vomitting |
| 12 | Measles morbillivirus (MMV) | Aerosol | Paramyxoviridae | Fever, muscle aches, swollen/watery eyes, mouth spots |
| 13 | Nipah henipavirus (NPV) | Blood, fluids | Paramyxoviridae | Fever, headache, muscle aches, vomiting, diarrhea, respitory distress |
| 14 | Rabies lyssavirus (RBV) | Blood, fluids | Rhabdoviridae | Flu-like symptoms, fever, headache, malaise, cerebral dysfunction, anxiety, confusion, agitation |
| 15 | Rift Valley Fever Virus (RVFV) | Blood | Bunyavirirdae | Fever, hemorrage, headache/muscle ache, loss of appetite, sensitivity to light, |
| 16 | Zika Virus (Zika) | Blood | Flaviviridae | Fever, joint pain, rash, conjunctivitis, headache/muscle ache |
| 17 | Hepatitis C Virus (HCV) | Blood | Flaviviridae | Fever, joint pain, rash, jaundice, loss of appetite, nausea/vomitting, abnormally colored stool and urine |
| 18 | Monkeypox virus (MKP) | Blood | Poxviridae | Fever, headache/muscle ache, rash, lesions, chills, exhaustion |
| 19 | Hepatitis B Virus (HBV) | Blood | Hepadnaviridae | Fever, malaise, vomitting, abnominal pain, jaundice, loss of appetite, abnormally colored stool and urine |
| 20 | Herpes Simplex Virus Type 2 (HSV-2) | Fluids | Herpesviridae | Fever, headache, muscle and body aches, swollen lymph nodes (especially around groin), pain/ithcing around genitals, bumps/blisters around mouth, genitals, and anus, urethetal/vaginal discharge |
| 21 | Chlamydia trachomatis (Chla) | Fluids | Chlamydiaceae | Abnormal urethetal/vaginal discharge, burning or discomfort while urinating, rectal pain/discharge/bleeding |
| 22 | Treponema pallidum subsp. pallidum (Syph) | Fluids | Spirochaetaceae | Chancres at area of infection, rash at area of infection, fever, swollen lymph nodes, sore throat, headaches/muscle aches, malaise |
| 23 | Trichomonas vaginalis (Trich) | Fluids | Trichomonadidae | Abnormal urethetal/vaginal discharge, itching/burning of genitals, burning or discomfort while urinating |

**Table S2**. Clinical symptoms of organisms included in the bloodborne pathogen panels.

#### Supporting Figures

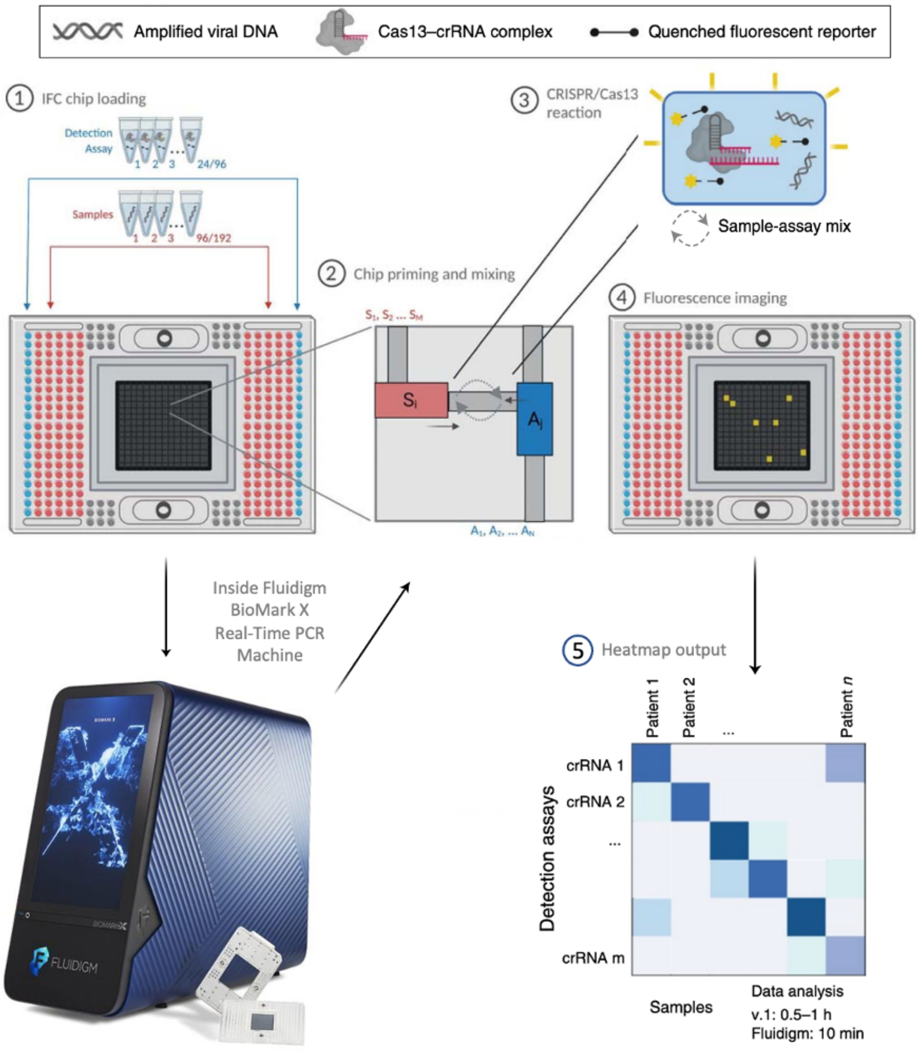

**Figure S1. Detailed CARMEN-BiomarkX workflow. (1)** In the Standard BioTools 192.24 Dynamic Array integrated fluidic circuit (IFC) chip, amplified samples (up to 192) are introduced into microwells (highlighted in red). Simultaneously, up to 24 detection assay reactions, each containing Cas13-crRNAs resuspended in buffer, are added to the sides of the IFC (highlighted in blue). The loaded IFC is then inserted into the BiomarkX instrument. **(2)** The protocol includes chip priming and mixing within a specialized IFC chamber. **(3)** In this chamber, sequence-specific crRNA complexes recognize and bind to RNA, initiating the cleavage activity of Cas13. Once activated, Cas13 cleaves the quenched fluorescent reporters within the system. **(4)** The BiomarkX chamber captures images at 37ºC in the ROX and FAM channels every 5 minutes for 1 hour following the initiation of the reaction. **(5)** This monitoring enables the observation of fluorescence resulting from the detections. Figure adapted from Welch et al., *Nature*, 2022.

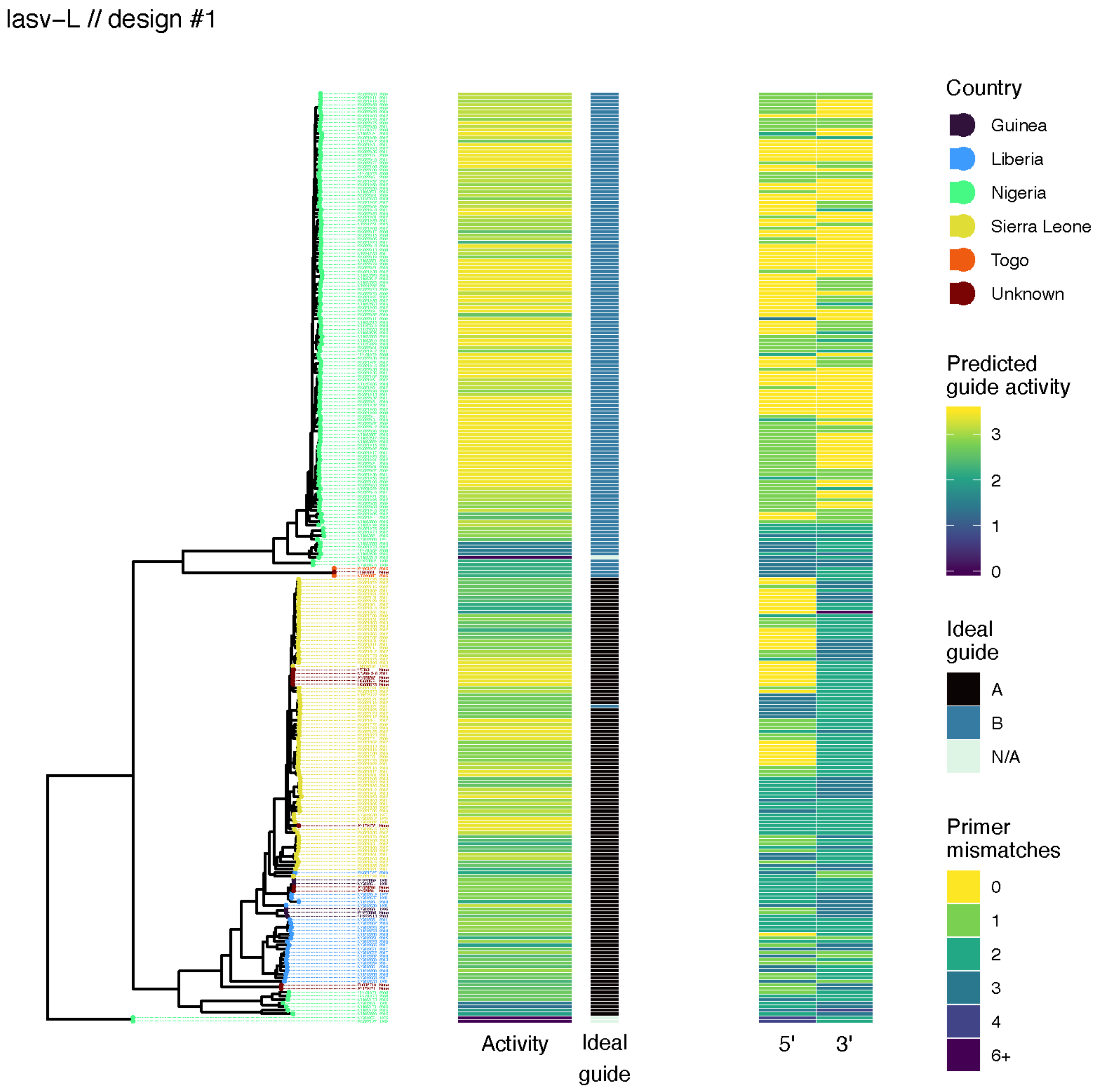

**Figure S2.** Predicted Lassa lineage genomes covered by the dual-crRNA system.

denv // design #1

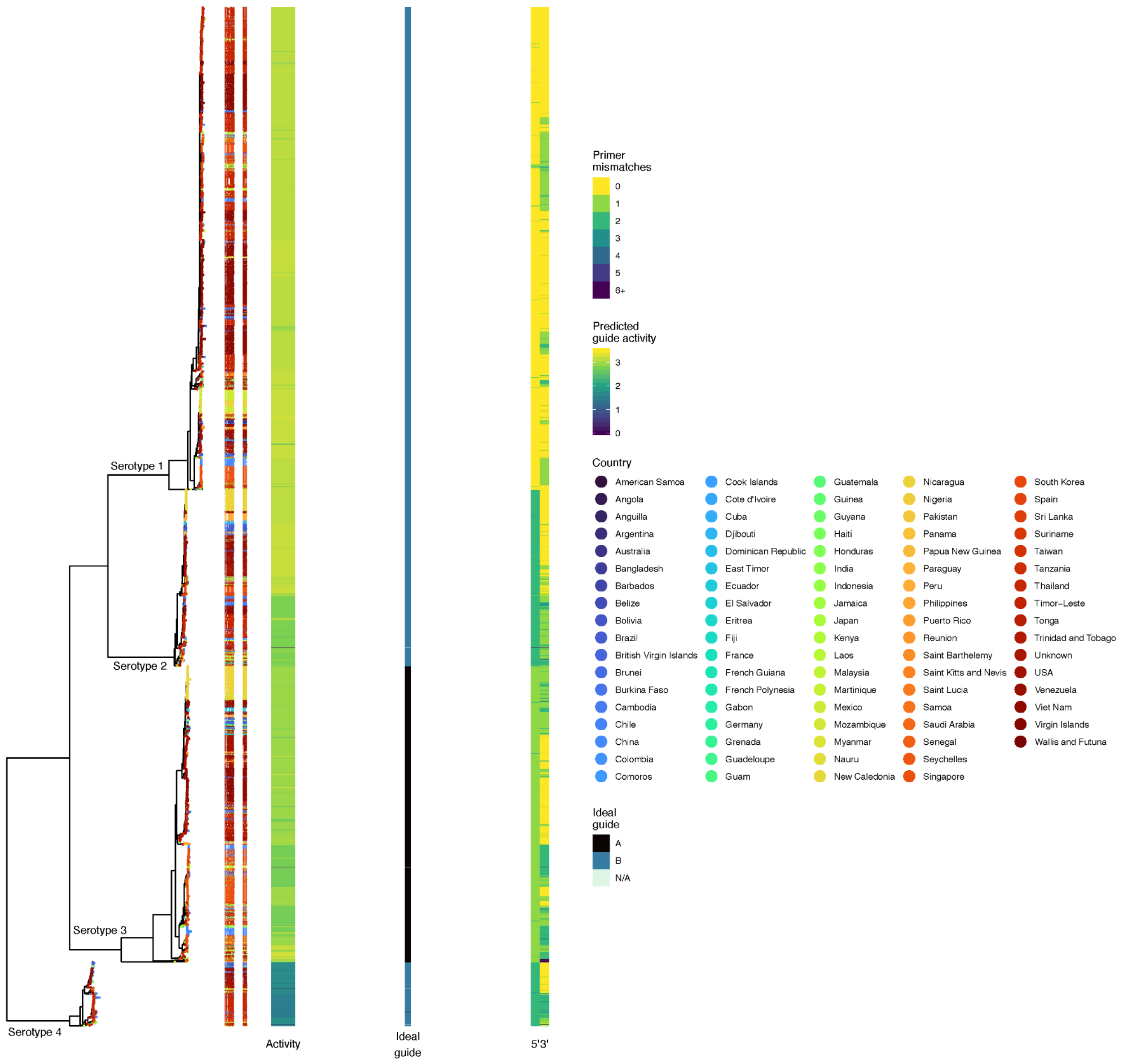

**Figure S3.** Predicted DENV genome lineages covered by the dual-crRNA and primer designs.

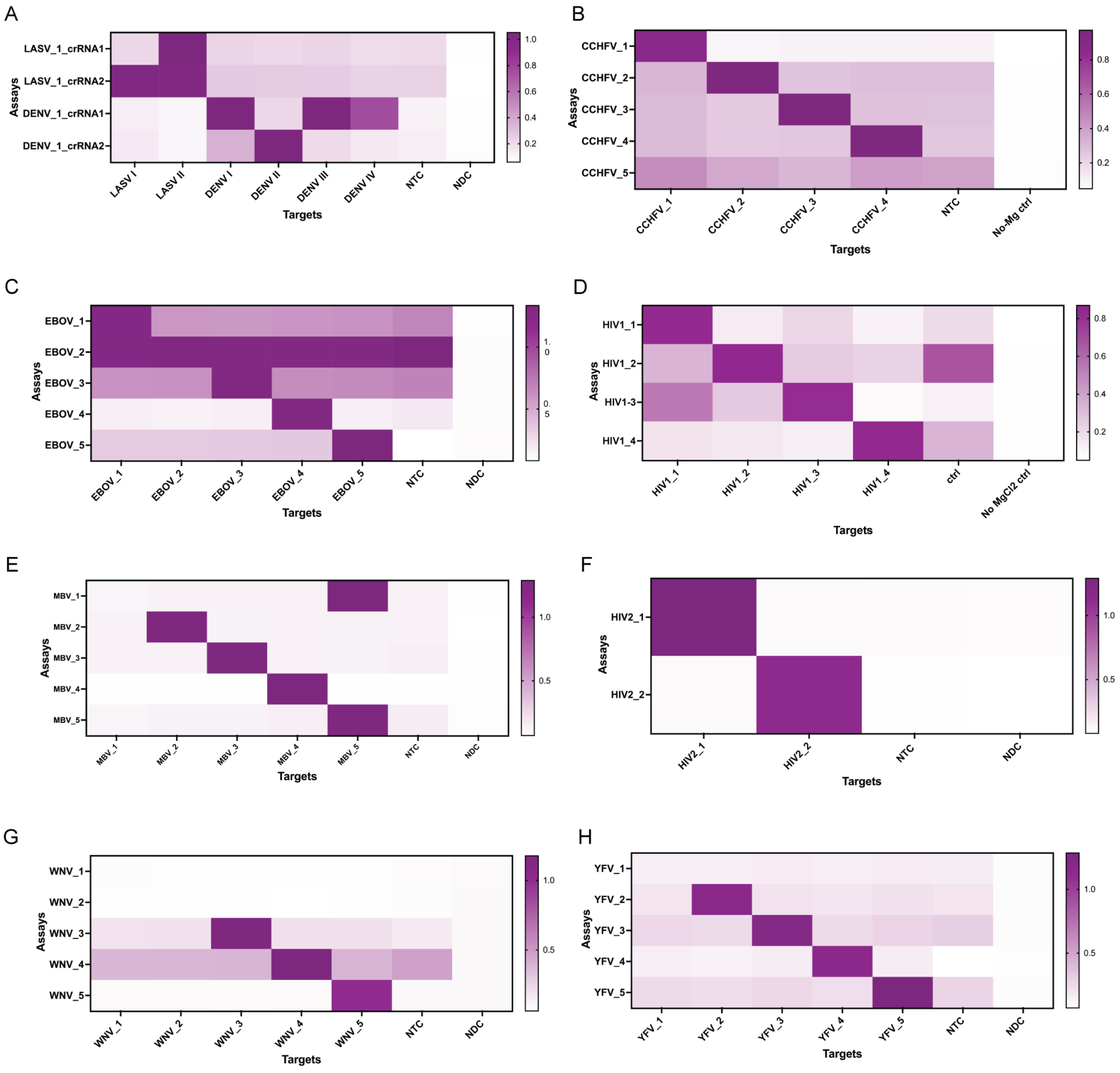

**Figure S4. Downselection of BBP1 assay design**. Heatmaps illustrating CARMEN assay performance for (**A**) *Lassa virus* (LASV) and *Dengue virus* (DENV), (**B**) Congo (CCHFV), (**C**) Ebola virus (EBOV), (**D**) *Human immunodeficiency virus 1* (HIV1), (**E**) *Marburg virus* (MBV), (**F**) *Human immunodeficiency virus 2* (HIV2), (**G**) *West-nile virus* (WNV), and (**H**) *Yellow fever virus* (YFV), showing fluorescence intensity of synthetic DNA fragments, at $10^8$ copies per µL, and corresponding viral Cas13 crRNAs at 1 hour post-reaction initiation.

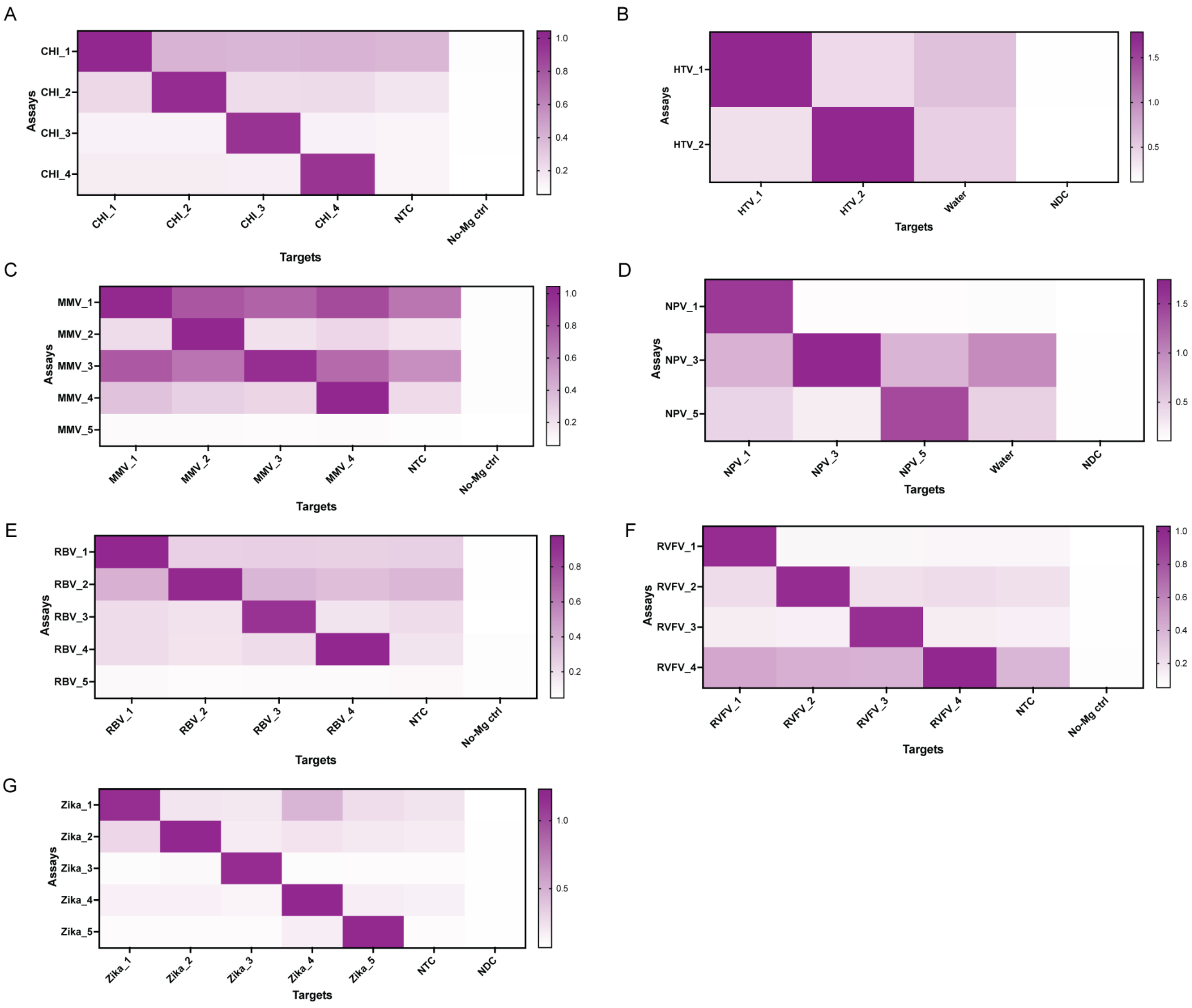

**Figure S5. Downselection of BBP2 assay design**. Heatmaps illustrating CARMEN assay performance for (**A**) *Chikungunya virus* (CHI), (**B**) *Hantaan orthohantavirus* (HTV), (**C**) *Measles morbillivirus* (MMV), (**D**) *Nipah henipavirus* (NPV), (**E**) *Rabies lyssavirus* (RBV), (**F**) *Rift Valley Fever virus* (RVFV), and (**G**) *Zika virus* (Zika), showing fluorescence intensity of synthetic DNA fragments, at $10^8$ copies per µL, and corresponding viral Cas13 crRNAs at 1 hour post-reaction initiation.

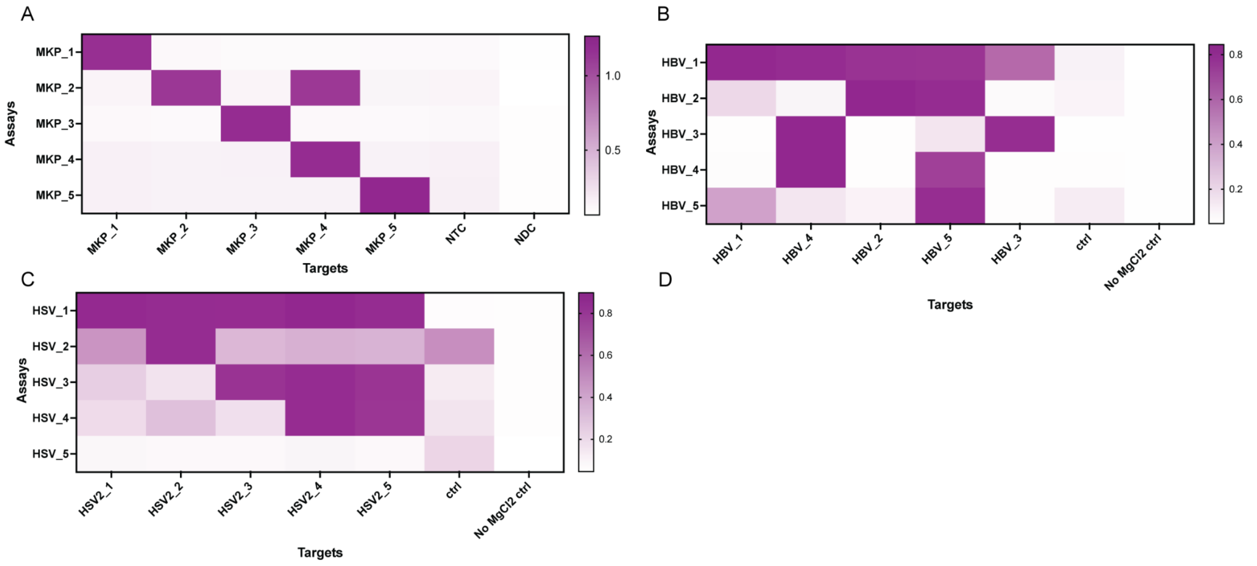

**Figure S6. Downselection of BBP3 assay design**. Heatmaps illustrating CARMEN assay performance for (**A**) *mpox virus* (MKP, formerly *monkeypox virus*), (**B**) *Hepatitis B virus* (HBV), and (**C**) *Herpes Simplex Virus Type 2* (HSV2), showing fluorescence intensity of synthetic DNA fragments, at $10^8$ copies per μL, and corresponding viral Cas13 crRNAs at 1 hour post-reaction initiation.

A

| | CARMEN LoD | qPCR LoD |
|---|---|---|
| **CCHFV** | 1.00E+01 | 1.00E+04 |
| **EBOV** | 1.00E+03 | 1.00E+04 |
| **HIV1** | 1.00E+01 | 1.00E+03 |
| **HIV2** | 1.00E+01 | 1.00E+04 |
| **LASV** | 1.00E+01 | 1.00E+03 |
| **MBV** | 1.00E+01 | 1.00E+04 |
| **WNV** | 1.00E+02 | 1.00E+05 |
| **YFV** | 1.00E+02 | 1.00E+04 |

B

| | CARMEN LoD | qPCR LoD |
|---|---|---|
| **CHI** | 1.00E+02 | 1.00E+05 |
| **DENV** | 1.00E+04 | 1.00E+04 |
| **HTV** | 1.00E+01 | 1.00E+05 |
| **MMV** | 1.00E+02 | 1.00E+02 |
| **NPV** | 1.00E+03 | 1.00E+02 |
| **RBV** | 1.00E+02 | 1.00E+08 |
| **RVFV** | 1.00E+02 | 1.00E+05 |
| **Zika** | 1.00E+01 | 1.00E+05 |
| **HCV** | 1.00E+04 | 1.00E+05 |

C

| | CARMEN LoD | qPCR LoD |
|---|---|---|
| **MKP** | 1.00E+01 | 1.00E+01 |
| **HBV** | 1.00E+02 | 1.00E+02 |
| **HSV-2** | 1.00E+02 | 1.00E+03 |
| **Chla** | 1.00E+01 | 1.00E+04 |
| **Syph** | 1.00E+02 | 1.00E+02 |
| **Trich** | 1.00E+02 | 1.00E+04 |

D

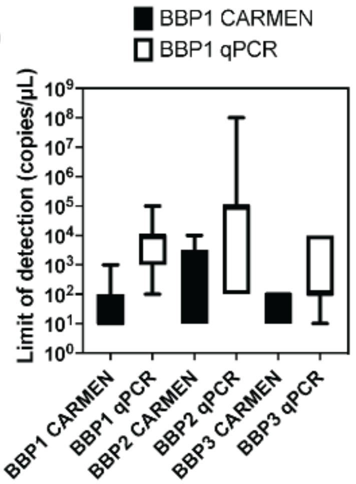

**Figure S7. Benchmarking CARMEN and RT-qPCR limits of detection**. Comparison of the limits of detections for (**A**) BBP1, (**B**) BBP2, and (**C**) BBP3 targets using CARMEN or RT-qPCR analysis. (D) Comparison plot between CARMEN and qPCR LODs. Data represent average LODs and standard deviations across all targets for each panel.

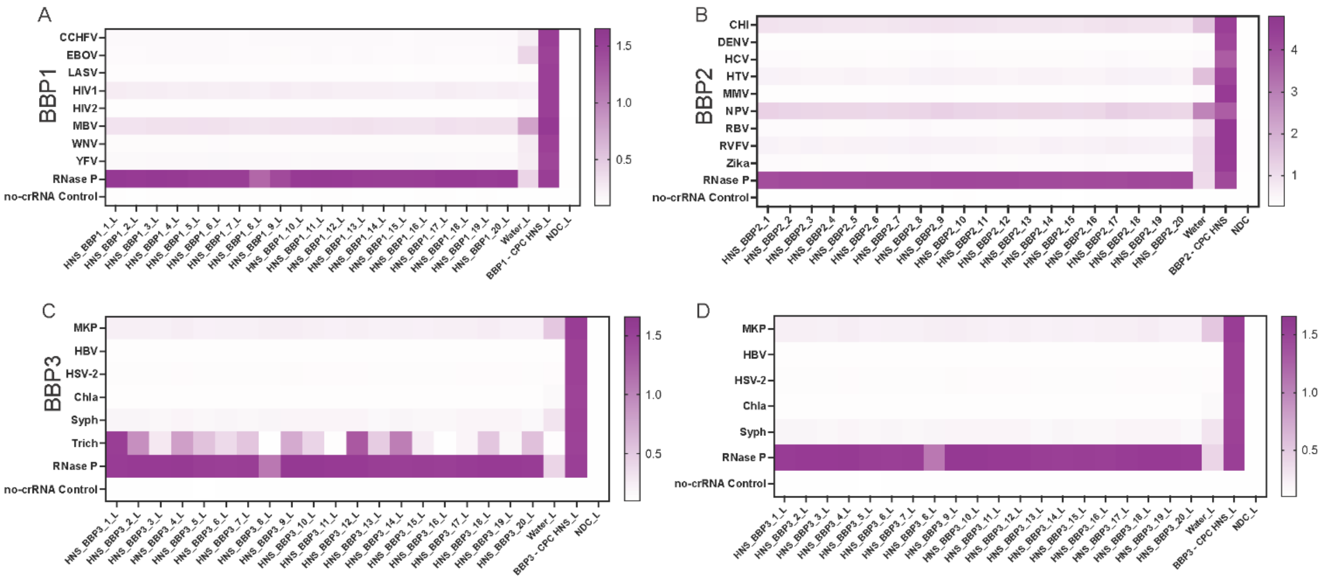

**Figure S8. Sensitivity and specificity analysis with contrived human normal samples**. Heatmaps illustrating CARMEN background signal in healthy normal serum (HNS) for (**A**) BBP1, (**B**) BBP2, and (**C**) BBP3 panels, showing fluorescence intensity and corresponding viral Cas13 crRNAs at 1 hour post-reaction initiation. (**D**) Heatmap showing fluorescence values of BPP3 after removal of Trich.

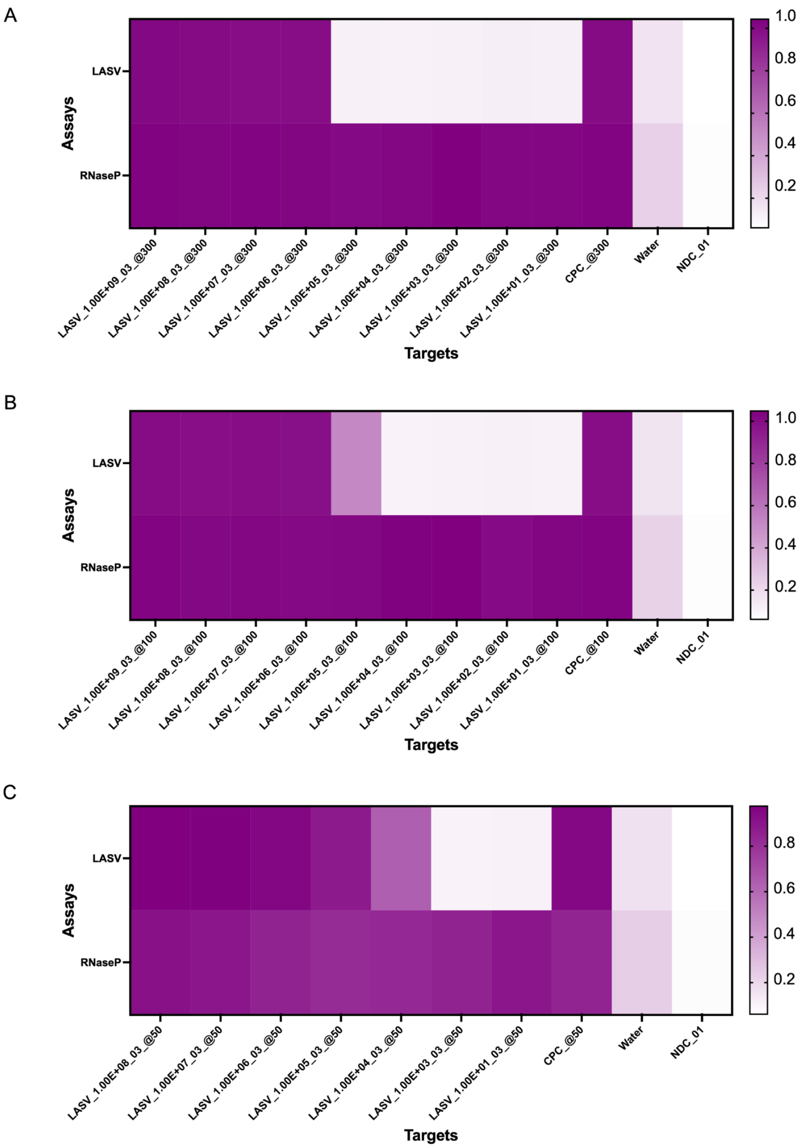

**Figure S9. Rnase P primer concentration analysis with contrived human normal samples spiked with synthetic LASV**. Heatmap illustrating CARMEN background signal in healthy normal serum (HNS) spiked-in with LASV, at the indicated concentrations (ranging from $10^9$ to $10^1$ copies per µL), followed by CARMEN detection. The RNAse P control sample was amplified using (**A**) 300 nM, (**B**) 100 nM, or (**C**) 50 nM primer concentration. Normalized fluorescence signal at 1 h post-reaction initiation of corresponding viral Cas13 crRNAs is shown.

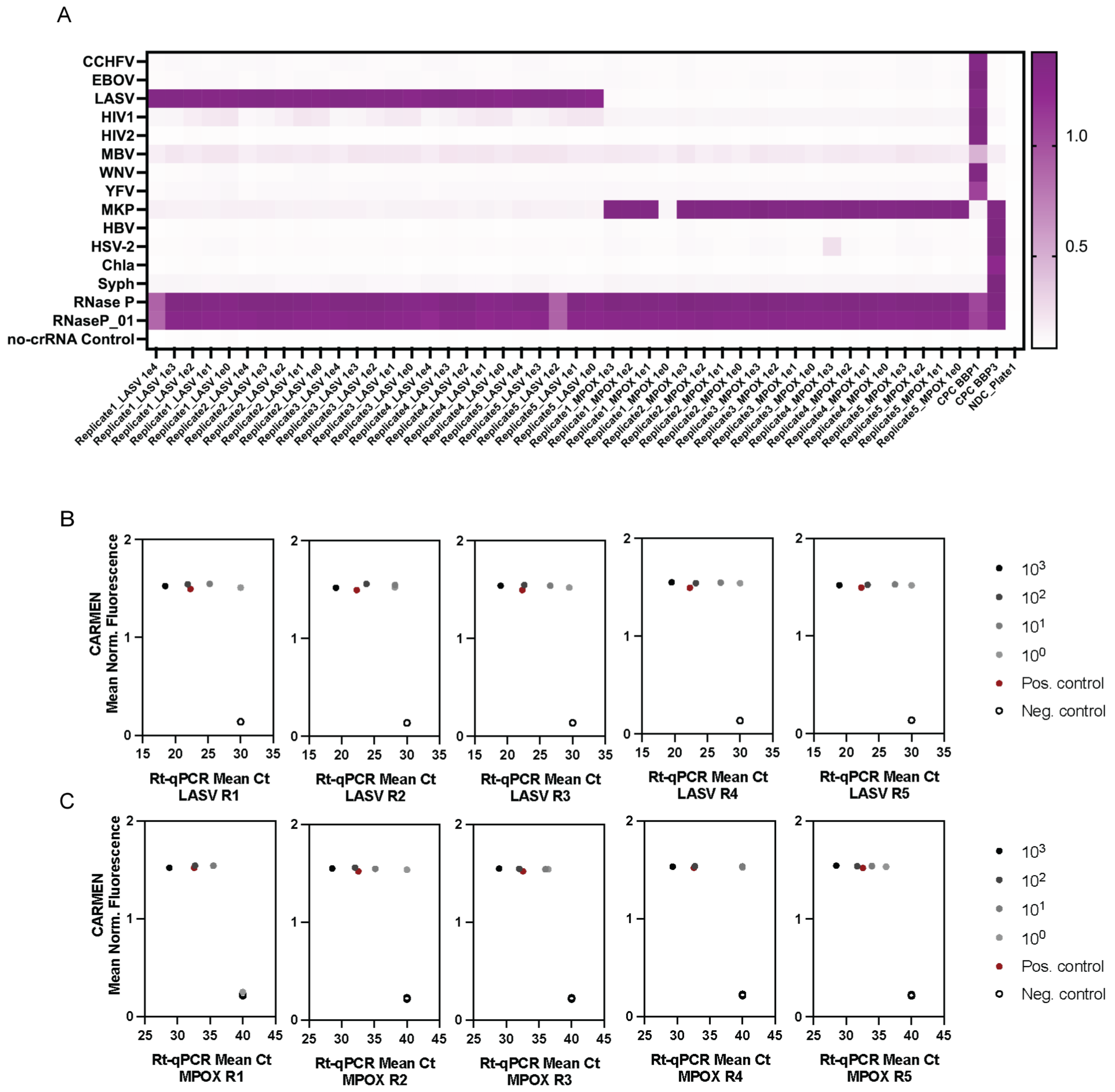

**Figure S9. Sensitivity and specificity analysis with contrived human normal samples spiked with LASV RNA and MPOX DNA extracts**. (**A**) Heatmap illustrating CARMEN background signal in healthy normal serum (HNS) spiked-in with LASV or MPOX RNA extracts, at the indicated concentrations (ranging from $10^3$ to $10^0$ copies per µL), followed by CARMEN detection with BBP1 and BBP3 respectively. Normalized fluorescence signal at 1 h post-reaction initiation of corresponding viral Cas13 crRNAs is shown. (**B-C**) Comparison analysis of CARMEN fluorescence to RT-qPCR Ct values for (**B**) LASV and (**C**) MPOX HNS samples, at the indicated concentrations (ranging from $10^3$ to $10^0$ copies per µL). Experiments were run in replicates of five.

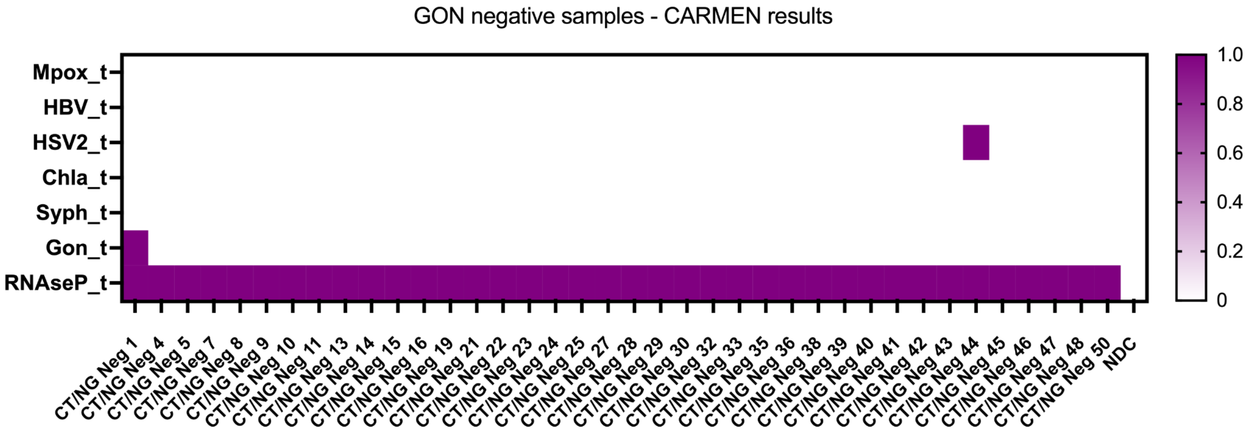

**Figure S10. Sensitive, accurate, detection of Gonorrhea in patient samples**. Heatmap illustrating CARMEN detection of *Neisseria gonorrhoeae* clinically negative patient samples. For this heatmap, normalized fluorescence signal at 1 h post-reaction initiation of corresponding viral Cas13 crRNAs is shown.

### CARMEN Blood Borne Pathogen Panel (CARMEN-BBP) Standard Operating Protocol

*CARMEN-BBP Standard Operating Protocol was constructed and modified from the mCARMEN-RVP Standard Operating Protocol.*

#### Table of Contents

I. Intended Use
II. Summary and Explanation
III. Principles of the Procedure
   - Materials and Reagents Required
   - Instrumentation, Equipment and Consumables Required
   - Warnings and Precautions
   - Reagent Storage, Handling, and Stability
   - Specimen Collection, Handling, and Storage
   - Reagent, Controls, and Equipment Preparation
IV. Detailed Assay Protocol
   - Nucleic Acid Extraction
   - RT-PCR master mix preparation and RT-PCR amplification
   - Preparation of Cas13 Detection reactions and Sample Master Mix plate Priming and loading the Fluidigm IFC
   - Running the IFC on the IFC Controller
   - Running and imaging on the Fluidigm Biomark
   - Data Analysis Using CARMEN-RVP Software
   - Interpretation of Results and Reporting
V. Appendix 1: CARMEN-BBP Primers, crRNA, and Reporter Sequences
VI. Appendix 2: Detailed interpretation of control results
VII. Appendix 3: In vitro transcription of CARMEN-BBP gene fragments for LoD testing, contrived sample evaluation, and CPC

### I. Intended Use

The CARMEN-BBP platform is a multiplexed CRISPR-Cas13-based test which is based on and modified from the original mCARMEN-RVP platform. CARMEN-BBP is intended for qualitative detection of nucleic acid from 23 distinct blood borne pathogens in blood serum from individuals suspected of febrile infection of unknown origin, including Lassa Virus, Ebola Virus, and Dengue Virus, or sexually transmitted diseases, such as Hepatitis B, Chlamydia trachomatis, and Trichomonas vaginalis, by their healthcare provider. Additionally, an internal human control, Ribonuclease P is tested alongside pathogen detection. To date, testing has been done by synthetic target detection, contrived samples composed of known concentrations of genomic RNA spiked into healthy human serum samples, and clinical samples tested at The African Centre of Excellence for Genomics of Infectious Diseases (ACEGID).

Results are available for all pathogens listed in Table 1. Pathogen DNA/RNA is generally detectable in blood serum specimens during the acute phase of infection. Positive results are indicative of the presence of pathogen DNA or RNA; clinical correlation with patient history and other diagnostic information is necessary to determine patient infection status. Positive results do not rule out bacterial infection or infection with other pathogens not included in the CARMEN-BBP1, 2, 3 assays. The results of this test should not be used as the sole basis for diagnosis, treatment, or other patient management decisions. The agent detected may not be the definite or only cause of symptoms or disease.

Negative results should be considered presumptive and do not preclude current or future infection with pathogens targeted by the assay, obtained through community transmission or other exposures. Negative results should not be used as the sole basis for diagnosis, treatment or other patient management decisions. Negative results obtained from individuals who are exhibiting symptoms associated with any pathogen target tested at the time of specimen collection should be interpreted with particular caution. Negative results must be considered in the context of an individual's recent exposures, history, presence of clinical signs and symptoms consistent with pathogens tested for by this platform.

#### II. Summary and Explanation

CARMEN is a CRISPR/Cas13-based assay for detection of nucleic acid from up to 24 separate targets in up to 184 patient samples. CARMEN has been adapted for multiple use cases including a Respiratory Viral Panel (RVP, Table 1), Blood-borne pathogens panel (BBP, Table 2), and an STI panel (Table 3). CARMEN has been validated for use on source samples such as nasal swabs and serum but could further be adapted to other source samples such as skin swabs, urine, feces, wastewater or more.

**SOP Table 1:** Pathogens targeted by CARMEN-BBP

| Pathogen Name | Panel | Acronym | Gene Targeted |
|---|---|---|---|
| Crimean Congo Hemorrhagic Fever Virus | CARMEN-BBP-1 | CCHFV | L segment |
| Zaire ebolavirus | CARMEN-BBP-1 | EBOV | Matrix protein |
| Human Immunodeficiency Virus 1 | CARMEN-BBP-1 | HIV1 | Pol protein |
| Human Immunodeficiency Virus 2 | CARMEN-BBP-1 | HIV2 | Gag polyprotein |
| Lassa Virus | CARMEN-BBP-1 | LASV | L segment |
| Marburg Virus | CARMEN-BBP-1 | MBV | Membrane associated protein |
| West-Nile Virus | CARMEN-BBP-1 | WNV | Nonstructural protein NS2A |
| Yellow Fever Virus | CARMEN-BBP-1 | YFV | RNA-dependent RNA polymerase |
| Chikungunya Virus | CARMEN-BBP-2 | CHI | Structural polyprotein |
| Dengue Virus | CARMEN-BBP-2 | DENV | Polyprotein/Type 2 vector |
| Rift Valley Fever Virus | CARMEN-BBP-2 | RVFV | L segment |
| Zika Virus | CARMEN-BBP-2 | ZIKV | Nonstructural protein NS3 |
| Measles morbillivirus | CARMEN-BBP-2 | MMV | IC323 gene |
| Rabies lyssavirus | CARMEN-BBP-2 | RBV | Nucleoprotein N |
| Hepatitis C Virus | CARMEN-BBP-2 | HCV | Polyprotein |
| Hantaan orthohantavirus | CARMEN-BBP-2 | HTV | Segment M |
| Nipah henipavirus | CARMEN-BBP-2 | NPV | Polymerase L gene |
| Monkeypox Virus | CARMEN-BBP-3 | MKP | A26L |

| | | | |
|---|---|---|---|
| Hepatitis B Virus | CARMEN-BBP-3 | HBV | Large S envelope protein |
| Herpes Simplex Virus Type 2 | CARMEN-BBP-3 | HSV-2 | Glycoprotein B (UL27) |
| Chlamydia trachomatis | CARMEN-BBP-3 | Chla | 16S |
| Treponema pallidum subsp. pallidum | CARMEN-BBP-3 | Syph | PolA |
| Trichomonas vaginalis | CARMEN-BBP-3 | Trich | 18S |
| Ribonuclease P | CARMEN-BBP-1/2/3 | RNase P | Ribonuclease P/MRP subunit (POP7) |

#### III. Principles of the Procedure

The CARMEN platform is a molecular *in vitro* diagnostic test that aids in the detection and differentiation of nucleic acid from up to 23 different pathogen targets and a human internal control. CARMEN is based on widely used nucleic acid extraction and amplification (RT-PCR) technology, a novel CRISPR-Cas13 detection system, and imaging of fluorescence on the Standard BioTools (formerly known as Fluidigm) BiomarkX system.

The workflow, in brief, is as follows:

1. Nucleic acid is extracted from source samples with the ThermoFisher KingFisher Flex MagMAX Viral/Pathogen Nucleic Acid Isolation kit or similar automated extracted method.
2. Patient sample nucleic acid is combined with custom oligonucleotide primer pools and amplified using the QiagenOneStep RT-PCR kit.
3. Amplified product is combined with a Sample Master Mix that contains a quenched fluorescent reporter (FAM-7-PolyU Reporter) in a 96-well PCR plate format.
4. Separately, Cas13, T7-polymerase, and individual crRNAs (23 pathogen crRNAs and 1 human control crRNA) are combined to produce 24 Assay Master Mixes in a 3 column format in a 96-well PCR plate.
5. The Assay Master Mixes are pipetted into the assay inlets on a Standard BioTools 192.24 Integrated Fluidics Circuit (IFC).
6. The amplified products plus Sample Master Mix are pipetted into the Sample inlets on a Standard BioTools 192.24 IFC.
7. The Standard BioTools 192.24 IFC is placed on a Standard BioTools BioMarkX instrument where the IFC is loaded and mixed and then is imaged.
8. The output data is analyzed with a specially designed CARMEN Detection Software. The CARMEN Detection Software automatically validates the various controls, and then makes one of two calls, “positive” or “negative”, for every combination of sample and pathogen target based off of a unique threshold calculated from the performance of the negative template control (NTC).

##### Materials and Reagents

Materials and reagents are required at each of the 5 physical areas of the CARMEN workflow:

1. Pre-amplification / isolation of DNA/RNA from sample
2. Pre-amplification / Master Mix manufacturing clean area
3. Pre-amplification / RT-PCR spike-in target area
4. Post-amplification / Post-RT-PCR spike-in target area
5. Post-amplification / Standard BioTools hardware

**Note**: Step 1 occurs in a BSL2+ lab environment within a biosafety cabinet. Steps 2, 3, 4 occur in designated dead air boxes.

Materials and reagents are also broken down into 3 broad categories related to their use in the workflow: (1) Extraction, (2) Amplification, and (3) Detection,

Materials & reagents are listed below in *SOP Table 2: Materials and Reagents*

**SOP Table 2:** Materials and Reagents

| Reagent Category | Component/Kit | Manufacturer | Catalog # | Storage Conditions | Areas Needed |
|---|---|---|---|---|---|
| Extraction | Quick-DNA/RNA™ MagBead | Zymo Research | R2131 | RT | 1 |

| | | | | | |
|---|---|---|---|---|---|
| Amplification | QIAGEN OneStep RT-PCR Kit | QIAGEN | 210215 | -20C | 2 |
| Amplification | Primers (forward and reverse, per target, per *Appendix 1: mCARMEN RVP Primers, crRNA, and Reporter sequences*) | IDT | Custom Order | −20℃ | 2 |
| Detection | CRISPR RNA (crRNA, per target, per *Appendix 1: mCARMEN-RVP Primers, crRNA, and Reporter sequences*) | IDT | Custom Order | −20℃ | 4 |
| Detection | Cas13 | Genscript | Custom Order/Z 03486 | −80℃ | 4 |
| Detection | RNase Inhibitor (Murine) | NEB | M0314L | −20℃ | 4 |
| Detection | HiScribe® T7 Quick High Yield RNA Synthesis Kit | NEB | E2050S | −20℃ | 4 |
| Detection | rNTP mix @ 25 mM each (mixed at 100 mM total) | NEB | N0450L | −20℃ | 4 |
| Detection | 16uM FAM-7-PolyU Reporter | IDT | Custom | −20℃ | 4 |
| Detection | ROX Reference Dye | Invitrogen | 1222301 2 | −20℃ | 4 |
| Detection | UltraPure™ 1 M Tris-HCI Buffer, pH 7.5 | Invitrogen | 1556702 7 | RT | 4 |
| Detection | USB Dithiothreitol (DTT), 0.1M Solution | Thermo Scientific | 707265 ML | −20℃ | 4 |

| | | | | | |
|---|---|---|---|---|---|
| Detection | $MgCl_2$ (1 M) | Invitrogen | AM9530 G | RT | 4 |
| Detection | Standard BioTools 20x Sample Loading Reagent | Standard BioTools | 100-6265 | −20℃ | 4 |
| Detection | Standard BioTools 2x Assay Loading Reagent | Standard BioTools | 100-6265 | −20℃ | 4 |
| Detection | Standard BioTools Actuation Fluid | Standard BioTools | 100-6265 | −20℃ | 4 |
| Detection | Standard BioTools Pressure Fluid | Standard BioTools | 100-6265 | −20℃ | 4 |
| Detection | Control Line Fluid Kit—192.24 | Standard BioTools | 100-6265 | RT | 4 |
| Detection | 192.24 GE Dynamic Array IFCs | Standard BioTools | 100-6265 | RT | 4 |
| All | Nuclease-Free Water (not DEPC-treated) | Invitrogen | AM9937 | RT | All |
| Extraction (Cleaner) | CITRUS II Deodorizing Cleaner | CITRUS II | CGDC04 6754 | RT | 1 |
| All (Cleaner) | 70% Ethanol | Any | Any | RT | All |
| All (Cleaner) | RNase AWAY® Spray | Thermo Scientific | 21-402-178 | RT | All |

#### Instrumentation, Equipment and Consumables

Instrumentation, Equipment and Consumable are required at each of the 5 physical areas of the CARMEN workflow:

1. Pre-amplification / isolation of DNA/RNA from sample
2. Pre-amplification / Master Mix manufacturing clean area
3. Pre-amplification / RT-PCR spike-in target area
4. Post-amplification / Post-RT-PCR spike-in target area
5. Post-amplification / Standard BioTools hardware

Note: Step 1 occurs in a BSL2+ lab environment within a biosafety cabinet. Steps 2, 3, 4 occur in designated dead air boxes.

Instrumentation, Equipment and Consumables are listed below in *SOP Table 3: Instrumentation required for CARMEN workflow, SOP Table 4: Equipment required for CARMEN workflow*, and *SOP Table 5: Consumables required for CARMEN workflow* .

**SOP Table 3:** Instrumentation required for CARMEN workflow

| Instrument | Manufacturer | Catalog # | Area Needed |
|---|---|---|---|
| KingFisher Flex Purification system | ThermoFisher | 5400630 | 2 |
| Mastercycler Pro Thermal Cycler | Eppendorf | EP6331000 025- 1EA | 3 |
| Standard BioTools BiomarkX | Standard BioTools | | 5 |
| GENIE® SI-0236 Vortex-Genie 2 Mixer | Cole-Parmer | EW-04724-00 | All |
| Microplate centrifuge, PCR Plate Spinner | ThermoFisher | 14-100-143 | All |
| Small refrigerator (4℃, temporary storage) or ice buckets | Any | Any | All |

**SOP Table 4:** Equipment required for CARMEN workflow

| Equipment | Manufacturer | Catalog # | Area Needed |
|---|---|---|---|
| Pipet-X Pipet Controller PX-100 | Rainin | 17011733 | 1 |
| Pipet-Lite Multi Pipette L8-200XLS+ | Rainin | 17013805 | 1, 2 (Should have dedicated pipet for both locations for CARMEN) |
| Pipet-Lite Multi Pipette L8- | Rainin | 17013803 | 1, 2, 3, 4 (Should have |

| | | | |
|---|---|---|---|
| 20XLS+ | | | dedicated pipet for all four locations for CARMEN) |
| Mylar™ Plate Sealer for Microtiter™ Plates, Mylar | ThermoFisher | 5701TS1 | All |
| Start Kit PL-LTS 2, 20, 200, 1000µL | Rainin | 30386597 | All (Should have dedicated set for all five locations for CARMEN) |

**SOP Table 5:** Consumables required for CARMEN workflow

| Consumable | Manufacturer | Catalog # | Area Needed |
|---|---|---|---|
| KingFisher Deepwell plates | ThermoFisher | 95040450 | 1 |
| Elution plates | ThermoFisher | 97002540 | 1 |
| Tip combs | ThermoFisher | A43074 | 1 |
| 50 mL serological pipettes | Any | Any | 1 |
| 96-well PCR plates | Any | Any | 2, 3, 4 |
| PCR strip tubes | Any | Any | 2, 3, 4 |
| Tubes LoBind (1.5 mL) | Any | Any | 2, 3, 4 |
| Tubes LoBind (5 mL) | Any | Any | 1, 2, 4 |
| Reagent reservoirs (50 mL) | Any | Any | All |
| Plate seals (Foil) | Any | Any | All |
| Kim wipes (large) | Any | Any | All |
| Pipette tips (P10) | Any (Compatible with Ranin pipettes) | Any (Compatible with Ranin pipettes) | All |
| Pipette tips (P20) | Any (Compatible with Ranin | Any (Compatible | All |

| | | | |
|---|---|---|---|
| | pipettes) | with Ranin pipettes) | |
| Pipette tips (P200) | Any (Compatible with Ranin pipettes) | Any (Compatible with Ranin pipettes) | All |
| Pipette tips (P1000) | Any (Compatible with Ranin pipettes) | Any (Compatible with Ranin pipettes) | All |

**Additional Instrumentation, Equipment and Consumables Not Explicitly Listed:**

- **Freezer/Refrigerated Storage:** −80℃, −20℃, 4℃
- **Appropriate PPE:** Gloves, safety glasses, face shields, lab coats/gowns, etc.
- **Various Laboratory Storage:** Tube racks, ice trays, freezer storage containers, etc.

#### Warnings and Precautions

1. Follow standard precautions. All patient specimens and positive controls should be considered potentially infectious and handled accordingly.
2. Do not eat, drink, smoke, apply cosmetics or handle contact lenses in areas where reagents and human specimens are handled.
3. Refer to Biosafety in Microbiological and Biomedical Laboratories (BMBL) 6th Edition BMBL (https://www.cdc.gov/labs/pdf/CDC-BiosafetyMicrobiologicalBiomedicalLaboratories-2020-P.pdf) for standard biological safety guidelines for all procedures.
4. Specimen processing should be performed in accordance with national biological safety regulations.
5. All patient samples, both suspected and not suspected cases of blood borne pathogen infection, based on clinical and epidemiological screening criteria recommended by public health authorities, should be collected using appropriate infectious control precautions.
6. Perform all manipulations of potentially infectious virus specimens within a Class II (or higher) biological safety cabinet.
7. Personal protective equipment such as, but not limited to, gloves, sleeves, and lab coats should be worn when handling kit reagents, while conducting laboratory work, and while handling materials, including samples, reagents, pipettes, and other laboratory equipment and chemicals.
8. Amplification technologies, such as RT-PCR and CRISPR-Cas13 detection assays, are sensitive to accidental introduction of products from previous amplification reactions and introduction of Ribonuclease P from the operator or from the external environment. Incorrect results could occur if either the clinical specimen or the reagents used in the amplification or detection steps become contaminated by accidental introduction of amplification product (amplicon) and/or Ribonuclease P.
9. Workflow in the laboratory should proceed in a unidirectional manner from clean environments to target based environments.
10. Maintain separate and dedicated areas for assay setup and handling of nucleic acids.
11. Maintain separate, dedicated equipment (e.g., pipettes, microcentrifuges) and supplies (e.g., microcentrifuge tubes, pipette tips) for assay setup and handling of extracted nucleic acids.
12. Always check the expiration date of reagents prior to use. Do not use expired reagent(s). Do not substitute or mix reagents from different kit lots or from other manufacturers without first testing compatibility.
13. Change pipette tips between all manual liquid transfers.
14. During preparation of samples, compliance with good laboratory techniques is essential to minimize the risk of cross-contamination between samples, and the inadvertent introduction of nucleases into samples during and after the extraction procedure. Proper aseptic technique should always be used when working with nucleic acids.
15. Wear a clean lab coat and powder-free disposable gloves (not previously worn) when setting up assays.

16. Change gloves between sample handling and whenever contamination is suspected.
17. Keep reagent and reaction tubes capped or covered as much as possible.
18. Primers, crRNA (including aliquots), enzyme Master Mix, reagents, and patient sample RNA must be thawed and maintained on cold block or ice at all times during preparation and use to ensure stability.
19. Work surfaces, pipettes, and centrifuges should be cleaned and decontaminated with cleaning products before and after usage. It is suggested to use 10% bleach or 5% lysol and "RNase AWAY® " to minimize risk of nucleic acid or nuclease contamination. Residual bleach should be removed using 70% ethanol. Workspaces that occur in hoods or biosafety cabinets should be treated with an internal UV light for 15 minutes prior to usage. In the event that bleach can not be utilized, such as in environments where guanidinium thiocyanate is present, an appropriate bleach alternative should be used, such as "CITRISII®". It should be noted that extraction lysis buffers containing guanidinium thiocyanate, or guanidine-containing materials, can create highly reactive and/or toxic compounds if combined with sodium hypochlorite (bleach).
20. Dispose of unused kit reagents and human specimens according to local, state, and federal regulations.

#### Reagent Storage, Handling, and Stability

- Store all primers and probes at the storage temperatures listed in the *Materials and Reagents Required* section (*SOP Table 2: Materials and Reagents*).
- Always check the expiration date prior to use. Do not use expired reagents.
- Many of the reagents, including Kingfisher reagents, FAM-7-PolyU Reporter, and ROX dye, are sensitive to light exposure. Please be aware and follow manufacturer's instructions for handling and storage.
- Primers, probes (including aliquots), and enzyme Master Mix must be thawed and kept on a cold block at all times during preparation and use.
- To ensure reagent quality, do not refreeze primers, probes, and aliquoted master mixes.
- Controls and aliquots of controls must be thawed and kept on ice at all times during preparation and use.

**Specimen Collection, Handling, and Storage**

*A. Specimen collection***.** The assay will be performed on samples from a matrix type that is consistent with pathogen presentation in symptomatic individuals. Samples should be collected following appropriate procedures.

*B. Specimen transport.* Samples should be transported at 2-8°C. If a delay in testing or shipping is expected store specimens at –80°C.

*C. Specimen storage.* Upon processing the specimen, make an aliquot of the specimen to be saved in the –80 °C freezer for long term storage:

- Label one 2 mL microcentrifuge tube with the appropriate specimen labels.
- Pipette 1.5 mL of the specimen from the original vessel after vortexing into the labeled microcentrifuge tube.
- Aliquots are saved in a –80 °C freezer.

*D. Storing Purified Specimen Nucleic Acid.* Store purified nucleic acids in a –80 °C freezer.

#### Reagent, Controls, and Equipment Preparation

Preparation of the CARMEN reagents and equipment is broken down into 8 steps:

1. Preparation of primer pools
2. Preparation of crRNA and Cas13 working stocks
3. Preparation of RT-PCR Master Mix
4. Preparation of Sample Master Mix

5. Preparation of Assay Master Mix
6. Preparation of controls
7. Preparation of equipment
8. Preparation of Sample Assignment Sheet

**Note**: In many cases, various combined reagents are listed below in excess of their exact needs, to account for pipetting and aliquoting errors. This includes the Sample Master Mix, the Assay Master Mix predicate, the Binding Bead Mix and RT-PCR Master Mix, and the individual aliquots of each primer pool**.**

**1. Preparation of equipment**

Clean and decontaminate all work surfaces, pipettes, centrifuges, and other equipment prior to and after use to minimize the risk of nucleic acid contamination. Decontamination agents include: 10% bleach, RNase AWAY®, and 70% ethanol. Workspaces that occur in hoods or biosafety cabinets should be treated with an internal UV light for 15 minutes prior to usage. In the event that bleach can not be utilized, such as in environments where guanidinium thiocyanate is present, an appropriate bleach alternative should be used, such as "CITRISII®". It should be noted that extraction lysis buffers containing guanidinium thiocyanate, or guanidine-containing materials, can create highly reactive and/or toxic compounds if combined with sodium hypochlorite (bleach).

**2. Preparation of primer pools (Target-free, pre-amplification clean-room environment):** Upon receipt, either (1) resuspend lyophilized primers to 100 µM with nuclease-free water (not DEPC-treated) (nmol concentration provided from manufacturer multiplied by 10) and pool the primers into two separate primer pools (one forward, one reverse). Follow instructions detailed in *SOP Table 6: Primer pools preparation* below. OR (2) Leave lyophilized primers at room temperature until needed.

After pooling primers to a working solution of 5 µM of each pathogen-specific primer and 0.83 µM RNase P primer, split the pools into four 700 µL aliquots into pre-labeled tubes.Store the primer pools and individual 100 µM primer stocks at -20°C.

**Note:** For Dengue Virus, and Hepatitis C Virus in the CARMEN-BBP 2 primer pool there are multiple reverse primers; this is to account for the sequence diversity of these pathogens.

**SOP Table 6:** Primer pools preparation

**BBP1:**

| **Forward Primer Pool** | | | **Reverse Primer Pool** | | |
|---|---|---|---|---|---|
| **Primer** | **Amount (µL)** | **Desired Concentration** | **Primer** | **Amount (µL)** | **Desired Concentration** |
| CCHFV_F | 140 | 5 µM | CCHFV_R | 140 | 5 µM |
| EBOV_F | 140 | 5 µM | EBOV_R | 140 | 5 µM |
| HIV1_F | 140 | 5 µM | HIV1_R | 140 | 5 µM |
| HIV2_F | 140 | 5 µM | HIV2_R | 140 | 5 µM |
| LASV_F | 140 | 5 µM | LASV_R | 140 | 5 µM |
| MBV_F | 140 | 5 µM | MBV_R | 140 | 5 µM |
| WNV_F | 140 | 5 µM | WNV_R | 140 | 5 µM |
| YFV_F | 140 | 5 µM | YFV_R | 140 | 5 µM |

| | | | | | |
|---|---|---|---|---|---|
| RNaseP_F | 23.24 | 0.83 µM | RNaseP_R | 23.24 | 0.83 µM |
| **Nuclease-free water (not DEPC-treated)** | **1656.76** | | **Nuclease-free water (not DEPC-treated)** | **1656.76** | |
| **TOTAL:** | **2800** | | **TOTAL:** | **2800** | |

**BBP2:**

| **Forward Primer Pool** | | | **Reverse Primer Pool** | | |
|---|---|---|---|---|---|
| **Primer** | **Amount (µL)** | **Desired Concentration** | **Primer** | **Amount (µL)** | **Desired Concentration** |
| CHI_F | 140 | 5 µM | CHI_R | 140 | 5 µM |
| DENV_F | 140 | 5 µM | DENV_R_1 | 140 | 5 µM |
| - | - | - | DENV_R_2 | 140 | 5 µM |
| - | - | - | DENV_R_3 | 140 | 5 µM |
| - | - | - | DENV_R_4 | 140 | 5 µM |
| RVFV_F | 140 | 5 µM | RVFV_R | 140 | 5 µM |
| ZIKV_F | 140 | 5 µM | ZIKV_R | 140 | 5 µM |
| MMV_F | 140 | 5 µM | MMV_R | 140 | 5 µM |
| RBV_F | 140 | 5 µM | RBV_R | 140 | 5 µM |
| HCV_F | 140 | 5 µM | HCV_R_1 | 140 | 5 µM |
| - | - | - | HCV_R_2 | 140 | 5 µM |
| HTV_F | 140 | 5 µM | HTV_R | 140 | 5 µM |
| NPV_F | 140 | 5 µM | NPV_R | 140 | 5 µM |
| RNaseP_F | 23.24 | 0.83 µM | RNaseP_R | 23.24 | 0.83 µM |
| **Nuclease-free water (not DEPC-treated)** | **1516.76** | | **Nuclease-free water (not DEPC-treated)** | **956.76** | |
| **TOTAL:** | **2800** | | **TOTAL:** | **2800** | |

**BBP3:**

| **Forward Primer Pool** | | | **Reverse Primer Pool** | | |
|---|---|---|---|---|---|
| **Primer** | **Amount** | **Desired** | **Primer** | **Amount** | **Desired** |

| | (µL) | Concentration | | (µL) | Concentration |
|---|---|---|---|---|---|
| MKP_F | 140 | 5 µM | MKP_R | 140 | 5 µM |
| HBV_F | 140 | 5 µM | HBV_R | 140 | 5 µM |
| HSV-2_F | 140 | 5 µM | HSV-2_R | 140 | 5 µM |
| Chla_F | 140 | 5 µM | Chla_R | 140 | 5 µM |
| Syph_F | 140 | 5 µM | Syph_R | 140 | 5 µM |
| Trich_F | 140 | 5 µM | Trich_R | 140 | 5 µM |
| RNase P_F | 23.24 | 0.83 µM | RNase P_R | 23.24 | 0.83 µM |
| **Nuclease-free water (not DEPC-treated)** | **1936.76** | | **Nuclease-free water (not DEPC-treated)** | **1936.76** | |
| **TOTAL:** | **2800** | | **TOTAL:** | **2800** | |

**3. Preparation of crRNA and Cas13 working stocks (Target-free, pre-amplification clean-room environment):**

**crRNA.** Upon receipt, either (1) Resuspend lyophilized crRNAs to 100 µM with nuclease-free water (not DEPC-treated) (nmol concentration provided from manufacturer multiplied by 10). Post-resuspension, make a 10 µM stock solution of each crRNA by combining 90 µL of nuclease-free water (not DEPC-treated) and 10 µL of 100 µM crRNA either in a 96-well PCR plate, a strip tube, or a 1.5 mL Lobind tube. Next, make a 1 µM working stock of each crRNA by combining 90 µL of nuclease-free water (not DEPC-treated) and 10 µL of 10 µM crRNA either in a 96-well PCR plate, a strip tube, or a 1.5 mL Lobind tube. This final 1 µM crRNA solution is what is used in the BBP-CARMEN protocol. OR (2) Leave lyophilized crRNAs at room temperature until needed.

**Cas13.** Upon receipt of Cas13, adjust working concentration with provided diluent from GenScript to be 0.55 mg/mL. If diluent is not provided, use an appropriate dilution buffer for Cas proteins. Store all Cas13 aliquots at 80°C.

**4. Preparation of RT-PCR Master Mix (Target-free, pre-amplification clean-room environment)**

- Ensure that primer pools are prepared ahead of making RT-PCR master mix.
- Make RT-PCR master mix per *SOP Table 7: RT-PCR master mix for 96 reactions* below.
- Aliquot 40 µL of RT-PCR master mix into a 96-well PCR plate.

**SOP Table 7:** RT-PCR master mix for 96 reactions

| **Reagent** | **Amount Per SIngle 50 µL Reaction (µL)** | **Amount Per 96 50 µL reactions + 10% Overage (µL)** |
|---|---|---|
| Qiagen 5x OneStep RT-PCR Buffer | 12.5 | 1320 |

| | | |
|---|---|---|
| Target Specific Primer Pool Forward | 3 | 316.8 |
| Target Specific Primer Pool Reverse | 3 | 316.8 |
| Qiagen dNTP mix | 2 | 211.2 |
| Qiagen Enzyme Mix | 2 | 211.2 |
| Target* | 10 | 0 |
| Nuclease-free water (not DEPC-treated) | 17.5 | 1848 |
| TOTAL: | 50 | 4224 |

**Target* –** To be added outside of the target-free, pre-amplification clean-room environment.

**5. Preparation of Sample Master Mix (Target-free, pre-amplification clean-room environment)**

**Note:** Mix all reagents listed in *SOP Table 7: 10x Cleavage Buffer,* in a 15 mL tube. Mix all reagents listed in *SOP Table 8: Sample Mastermix for 96 reactions,* in a 1.5 mL tube (keep all reagents on ice or on a cold block).

- Make 10x Cleavage Buffer per *SOP Table 8: 10x cleavage buffer* below Resuspend lyophilized FAM-7-PolyU Reporter to 100 uM with nuclease-free water (not DEPC-treated) (nmol concentration provided from manufacturer multiplied by 10). Post-resuspension, aliquot 16 uM working stocks of FAM-7-PolyU Reporter by with nuclease-free water (not DEPC-treated).
- Aliquot 1 mL of 10x Cleavage Buffer into 10 1.5 mL pre-labeled nuclease-free, sterile tubes (additional aliquots of 10x Cleavage Buffer can be stored between 20°C - 80°C)
- Make Sample Master Mix per *SOP Table 9: Sample master mix for 96 reactions* below.
- Aliquot 12.6 µL of sample master mix into a 96-well PCR plate. Protect reagents from light and keep on ice or on a cold block.

**SOP Table 8:** 10x cleavage buffer

| Reagent | Amount (mL) |
|---|---|
| 1M Tris-HCl (pH 7.5) | 4 |
| 0.1M DTT | 1 |
| Nuclease-free water (not DEPC-treated) | 5 |

| | |
|---|---|
| **TOTAL:** | **10** |

**SOP Table 9:** Sample master mix for 96 reactions

| Reagent | Amount Per SIngle 14 µL Reaction (µL) | Amount Per 96 14 µL reactions + 10% Overage (µL) |
|---|---|---|
| 10x Cleavage Buffer | 1.43 | 151 |
| NEB rNTPs | 0.57 | 60.2 |
| RNase Inhibitor (Murine) | 0.72 | 76 |
| 20x Sample Loading Reagent | 0.72 | 76 |
| FAM-7-PolyU Reporter (16 uM) | 0.45 | 47.5 |
| ROX dye (50x) | 0.03 | 3.2 |
| Nuclease-free water (not DEPC-treated) | 8.52 | 899.7 |
| $MgCl_2$ (1 M)* | 0.13 | 13.7 |
| Target* | 1.4 | 0 |
| **TOTAL:** | **14** | **1327.4** |

**MgCl2 (1 M)* –** Before adding in MgCl2 (1 M), take 12.6 uL of prepared solution and add to designated no detection control (NDC) position on a 96-well PCR plate.
**Target* –** To be added outside of the target-free, pre-amplification clean-room environment.

**6. Preparation of Assay Master Mix (Target-free, pre-amplification clean-room environment)**

- Mix assay master mix reagents listed in *SOP Table 10: Assay master mix for 24 reactions* below in a 1.5 mL nuclease-free tube on a cold block.
- Aliquot 12.6 µL of assay master mix to either a strip tube or 96-well PCR plate.
- Using aliquots of specific crRNAs prepared in *Step 2: Preparation of crRNA and Cas13 working stocks (Target-free, pre-amplification clean-room environment)* of *Reagent, Controls, and Equipment Preparation*, spike-in 3.4 µL of 1 µM of specific crRNAs to each well of 12.6 µL assay master mix.
- Each 16 µL well position is now a pathogen-specific crRNA assay mix for the CARMEN process.
- Follow *Figure 1: crRNA assay master mix layouts* for crRNA assay master mix formatting for the specific CARMEN panel that is being run.

- Be sure to include at least one “No-crRNA Control”. Instead of inputting 3.4 µL of a pathogen-specific crRNA, input 3.4 µL of nuclease-free water (not DEPC-treated).

**Note:** Some assays have multiple crRNAs, this is to account for the sequence diversity of these pathogens.

**SOP Table 10:** Assay master mix for 24 reactions

| Reagent | Amount Per SIngle 16 µL Reaction (µL) | Amount Per 96 14 µL reactions + 10% Overage (µL) |
|---|---|---|
| Cas13a Working Stock (0.55 mg/mL) | 0.9 | 23.8 |
| NEB T7 Polymerase | 2.5 | 66 |
| 2x Assay Loading Reagent | 8.2 | 216.5 |
| Nuclease-free water (not DEPC-treated) | 1.4 | 37 |
| 1 uM Pathogen-Specific crRNA | 3.4 | 0 |
| **TOTAL:** | **16** | **343.3** |

**Figure 1:** crRNA assay master mix layouts
**BBP1 crRNA assay master mix (strip tube or 96-well PCR plate):**

| 1 | 2 | 3 | 4 | 5 | 6 | 7 | 8 |
|---|---|---|---|---|---|---|---|
| CCHFV | EBOV | HIV1 | HIV2 | LASV | MBV | WNV | YFV |

| 9 | 10 | 11 | 12 | 13 | 14 | 15 | 16 |
|---|---|---|---|---|---|---|---|
| RNase P | No-crRNA Control | | | | | | |

| 17 | 18 | 19 | 20 | 21 | 22 | 23 | 24 |
|---|---|---|---|---|---|---|---|

**Note:** Well positions 11-24 do not have a pathogen-specific crRNA. Operators may opt to duplicate crRNA assays for technical replicates of each assay. To do so, make well positions 11-15 “No-crRNA Controls”, well position 16 a duplicate of RNase P, and well positions 17-24 duplicates of well positions 1-8.

**BBP2 crRNA assay master mix (strip tube or 96-well PCR plate):**

| 1 | 2 | 3 | 4 | 5 | 6 | 7 | 8 |
|---|---|---|---|---|---|---|---|
| CHI | DENV | RVFV | ZIKV | MMV | RBV | HCV | HTV |

| 9 | 10 | 11 | 12 | 13 | 14 | 15 | 16 |
|---|---|---|---|---|---|---|---|
| NPV | RNaseP | No-crRNA Control | | | | | |

| 17 | 18 | 19 | 20 | 21 | 22 | 23 | 24 |
|---|---|---|---|---|---|---|---|

**Note:** Well positions 12-24 do not have a pathogen-specific crRNA. Operators may opt to duplicate crRNA assays for technical replicates of each assay. To do so, make well positions 12-14 “No-crRNA Controls”, well positions 15 and 16 a duplicate of well positions 9 and 10, and well positions 17-24 duplicates of well positions 1-8.

**BBP3 crRNA assay master mix (strip tube or 96-well PCR plate):**

| 1 | 2 | 3 | 4 | 5 | 6 | 7 | 8 |
|---|---|---|---|---|---|---|---|
| MKP | HBV | HSV-2 | Chla | Syph | Trich | RNase P | No-crRNA Control |

| 9 | 10 | 11 | 12 | 13 | 14 | 15 | 16 |
|---|---|---|---|---|---|---|---|

| 17 | 18 | 19 | 20 | 21 | 22 | 23 | 24 |
|---|---|---|---|---|---|---|---|

**Note:** Well positions 9-24 do not have a pathogen-specific crRNA. Operators may opt to do triplicates of crRNA assays for technical replicates of each assay. To do so, make well positions 9-16 and 17-24 duplicates of well positions 1-8.

##### 7. Preparation of controls

The CARMEN protocol needs the following controls: extraction negative control (EC), no template control (NTC), combined positive controls (CPC), negative detection control (NDC), and no-crRNA control (no-crRNA). Prepare each as follows below (a $6^{th}$ control, RNase P, is included with the pathogen target primer and crRNA sets).

##### Extraction Control - Extraction negative Control (EC)

For EC, input 200 µL of nuclease-free water (not DEPC-treated) into well positions A1, A12, H1, and H12 of a 96-deep well extraction plate alongside clinical samples and extract as outlined in *Nucleic Acid Extraction*. The EC is used to check for contamination during the extraction protocol.

**RT-PCR Controls**

**i. No Template Control (NTC):**

For NTC, input 10 µL of nuclease-free water (not DEPC-treated) into at least one designated NTC well position on the RT-PCR plate alongside clinical samples and amplify as outlined in *RT-PCR master mix preparation and RT-PCR amplification*. The NTC is used to check for contamination during RT-PCR and detection plate set-up.

**ii. Combined Positive Control (CPC):**

Pool equal quantities of $10^5$ copies/µL of *in vitro* transcribed synthetic RNA or DNA of all pathogen targets on each specific panel (see *Appendix 3: In vitro transcription of CARMEN gene fragments for LoD testing, contrived sample evaluation, and CPC)* to create a $10^5$ CPC. Aliquot 12 µL of CPC in strip tubes and store at -80°C. To make the CPC, input 10 µL of $10^5$ CPC into at least one designated CPC well positioned on the RT-PCR plate alongside clinical samples and amplify as outlined in *RT-PCR master mix preparation and RT-PCR amplification*.

**Detection Controls**

**i. Negative Detection Control (NDC)**

The NDC is composed of sample master mix prior to the addition of $MgCl_2$ (see *Step 4: Preparation of Sample Master Mix (Target-free, pre-amplification clean-room environment)* of *Reagent, Controls, and Equipment Preparation*). Add 12.6 µL of sample master mix without $MgCl_2$ to at least one designated NDC well of a 96-well PCR plate while plating sample master mix. Be sure to add 1.4 µL of nuclease-free water (not DEPC-treated) to this well position to ensure correct concentration and volume of reagents. This is a negative control for the Standard BioTools detection step and should yield a negative result for all pathogen-specific targets on the panel.

**ii. No crRNA control (no-crRNA):**

The no-crRNA control is composed of 12.6 µL assay master mix and 3.4 µL of nuclease-free water (not DEPC-treated) to ensure correct concentration and volume of reagents. This is a negative control for the Standard BioTools detection step and should yield a negative result for all pathogen-specific targets on the panel.

**8. Preparation of Sample Assignment Sheet**

Prior to running a CARMEN experiment, determine the sample set you will be working from, and fill in this template with the sample IDs.

**Note**: no cells should be empty. If there are not samples in given wells, they should be given the label “empty” or “null”

#### IV. Detailed Assay Protocol

With the previous preparations on hand, the protocol itself can be broken down into five distinct steps:

1. Nucleic Acid Extraction
2. RT-PCR master mix preparation and RT-PCR amplification
3. Preparation of Cas13 Detection reactions and Sample Master Mix plate

4. Priming and loading the Standard BioTools Integrated Flow Circuit (IFC)
5. Running and imaging the IFC on the Standard BioTools Biomark

#### 1. Nucleic Acid Extraction

In this step both DNA and RNA will be extracted from patient samples using the Zymo Research Quick-DNA/RNA™ MagBead Kit on the KingFisher Flex Magnetic Particle Processor with 96 Deep-Well Head. All reagents (except for ethanol and isopropanol) are found in the Zymo Research Quick-DNA/RNA™ MagBead kit. Follow protocol steps starting at the "*Sample Preparation*" in the Quick-DNA/RNA™ MagBead protocol provided by Zymo Research (pages 6 – 9, catalog nos. R2130/R2131).

Pre-treat patient samples following recommendations from manufacturer for "Whole Blood (mammalian)" as outlined below:

1. Add 200µL of 2X Concentrate DNA/RNA Shield™ to 200µL of patient sample, mix well.
2. Add 8µL of Proteinase K to each sample, mix well, incubate at room temperature for 30 minutes.
3. Vortex samples and centridge samples at maximum speed for 2 minutes.
4. Transfer supernatant into a new nuclease-free tube and discard any precipitate.
5. Add 408µL of 100% isopropanol, mix well.
6. Store product on ice until use and continue to the "Total Nucleic Acid Purification" section of Zymo Research Quick-DNA/RNA™ MagBead protocol.
7. Prepare Sample Plate, Wash 1 Plate, Wash 2 Plate, Ethanol Plate 1, Ethanol Plate 2, and Elution Plate according to *SOP Table 10: Isolation plate set up*.
8. Add 200µL of sample from Step A, Section f into Sample Plate
9. Add KingFisher Flex 96-well tip comb to Sample Plate and run KingFisher Flex instrument with protocol file obtained from Zymo Research (R2130_Quick DNA RNA Magbead_KingFisherFlex_Copurification_v2.bdz).
10. Follow instructions detailed by manufacturer's KingFisher protocol for loading plates into instrument.
11. Post extraction, properly dispose of all plates except the Elution Plate. Store Elution Plate on ice or at -80°C until proceeding to Step 2 RT-PCR amplification.

**SOP Table 10:** Isolation plate set up

| Plate ID | Plate type | Reagent | Volume per well (µL) |
|---|---|---|---|
| Wash 1 | KingFisher Deepwell 96 Plate | Wash Buffer 1 | 500 |
| Wash 2 | KingFisher Deepwell 96 Plate | Wash Buffer 2 | 500 |
| Ethanol 1 | KingFisher Deepwell 96 Plate | 95-100% Ethanol | 500 |
| Ethanol 2 | KingFisher Deepwell 96 Plate | 95-100% Ethanol | 500 |

| | | | |
|---|---|---|---|
| Sample | KingFisher Deepwell 96 Plate | DNA/RNA Lysis Buffer | 200 |
| | | 95-100% Ethanol | 400 |
| | | **MagBiding Beads*** | 30 |
| Elution | KingFisher Deepwell 96 Plate | Nuclease-free water | 50 |

**MagBinding Beads*** – Ensure to mix well before adding to Sample Plate

#### 2. RT-PCR master mix preparation and RT-PCR amplification

In this step the preparation of the RT-PCR master mix and the amplification of synthetic target/clinical sample DNA or RNA with the Qiagen OneStep RT-PCR kit will be detailed.

Before starting:

1. If using a hood or a biosafety cabinet, treat the workspace with an internal UV light for 15 minutes prior to usage. If not, proceed to Next Step.
2. Clean bench using 10% bleach, RNase AWAY® spray, and 70% ethanol
3. Clean an ice bucket or cold block with bleach, RNase AWAY® and 70% ethanol.
4. If using an ice bucket, fill with ice.

Then, thaw the following reagents in ice bucket or on cold block:

- Qiagen 5x OneStep RT-PCR Buffer
- Qiagen dNTP mix
- CARMEN panel specific forward primer pool (see *Step 1: Preparation of primer pools (Target-free, pre-amplification clean-room environment) in Reagent, Controls, and Equipment Preparation*)
- CARMEN panel specific reverse primer pool (see *Step 1: Preparation of primer pools (Target-free, pre-amplification clean-room environment) in Reagent, Controls, and Equipment Preparation*)

Keep the following reagent in -20°C storage until needed:

- Qiagen enzyme mix

Prepare one 96-well PCR plate and label the plate with the following information:

- Unique experiment number/name
- Date of experiment
- Operator identifier

**Procedure for RT-PCR master mix preparations and amplification:**

This step requires the following reagents/kit components:

- Qiagen 5x OneStep RT-PCR Buffer
- Qiagen dNTP mix
- CARMEN panel specific forward primer pool (see *Step 1: Preparation of primer pools (Target-free, pre-amplification clean-room environment) in Reagent, Controls, and Equipment Preparation*)
- CARMEN panel specific reverse primer pool (see *Step 1: Preparation of primer pools (Target-free, pre-amplification clean-room environment) in Reagent, Controls, and Equipment Preparation*)
- Qiagen enzyme mix

**Location**: Target-free, pre-amplification clean-room environment

1. Ensure cleaning procedures have been followed before starting.
2. Mix the RT-PCR master mix according to *SOP Table 12: RT-PCR master mix preparation* in a sterile nuclease-free 5 mL tube and mix by inverting several times.
3. Aliquot 40 µL of the RT-PCR master mix into the pre-labeled 96-well PCR plate. Add 10 µL of water to at least one designated amplification negative control well (NTC).
4. Seal the plate with a foil seal before transferring to a designated area for working with synthetic target/patient sample DNA or RNA.
5. Keep the RT-PCR master mix plate on ice, on a cold block, or in a 4°C storage refrigerator until needed.

**SOP Table 12:** RT-PCR master mix preparation

| Reagent | Amount Per SIngle 50 µL Reaction (µL) | Amount Per 96 50 µL reactions + 10% Overage (µL) |
|---|---|---|
| Qiagen 5x OneStep RT-PCR Buffer | 12.5 | 1320 |
| Target Specific Primer Pool Forward | 3 | 316.8 |
| Target Specific Primer Pool Reverse | 3 | 316.8 |
| Qiagen dNTP mix | 2 | 211.2 |
| Qiagen Enzyme Mix | 2 | 211.2 |
| Target* | 10 | 0 |
| Nuclease-free water (not DEPC-treated) | 17.5 | 1848 |

| TOTAL: | 50 | 4224 |
|---|---|---|

**Target* –** To be added outside of the target-free, pre-amplification clean-room environment.
**Note –** Double master mix calculations above if two unique and separate plates of samples are to be run through workflow.

This step requires the following reagents/kit components:

- Prepared RT-PCR master mix plate
- Synthetic target/patient sample DNA or RNA

**Location**: Pre-amplification, target addition in BSL2+ environment

1. Ensure cleaning procedures have been followed before starting. Using a microplate centrifuge PCR plate spinner, centrifuge the prepared RT-PCR master mix plate to ensure that all liquid is collected at the bottom of the well positions. Add 10 µL of synthetic target or patient sample DNA or RNA from the clinical sample elution prepared on the KingFisher to the 96-well PCR plate containing the RT-PCR Master Mix.
2. Add 10 µL of 1E5 combined positive control (CPC) to at least one designated positive control well.
3. Seal the plate with a foil seal and ensure that all edges are fully sealed.
4. Lightly vortex the plate to adequately mix RT-PCR master mix reagent and synthetic target/patient sample DNA or RNA.
5. Using a microplate centrifuge PCR plate spinner, centrifuge the prepared plate.
6. Load the plate onto the thermocycler and run the cycling conditions listed in *SOP Table 13: Amplification conditions* below.
7. After the protocol has finished, proceed to Step 3 or store RT-PCR product at -20°C until needed.

**SOP Table 13:** Amplification conditions

| Step | Temperature | Time | Number of Cycles |
|---|---|---|---|
| Reverse Transcription | 50°C | 30 min | 1 |
| RT inactivation/ Initial denaturation | 95°C | 15 min | 1 |
| Amplification | 94°C | 30 s | 40 |
| | 58°C | 30 s | |
| | 72°C | 30 s | |

| | | | |
|---|---|---|---|
| Final Extension | 72°C | 5 min | 1 |
| End | 4°C | Hold | Indefinite |

**3. Preparation of sample and assay master mixes and amplified RT-PCR product spike-in.**

In this step the preparation of the sample and assay master mix plates will be detailed.

**Before starting:**

1. If using a hood or a biosafety cabinet, treat the workspace with an internal UV light for 15 minutes prior to usage. If not, proceed to Step 2.
2. Clean bench using 10% bleach, RNase AWAY® spray, and 70% ethanol
3. Clean an ice bucket or cold block with bleach, RNase AWAY® and 70% ethanol. If using an ice bucket, fill with ice.

Thaw the following reagents in ice bucket or on cold block:

- Sample master mix:
  - 10x Cleavage Buffer
  - NEB rNTPs
  - 20x Sample Loading Reagent
  - FAM-7-PolyU Reporter (16 uM) (Keep shielded from light)
  - ROX dye (50x) (Keep shielded from light)
  - Nuclease-free water (not DEPC-treated)
  - $MgCl_2$ (1 M)
- Assay master mix:
  - 2x Assay Loading Reagent
  - Nuclease-free water (not DEPC-treated)
  - Pathogen-Specific crRNA

Keep the following reagent in -20°C storage or on ice until needed:

- Sample master mix:
  - RNase Inhibitor (Murine)
- Assay master mix:
  - Cas13a Working Stock (0.55 mg/mL)
  - NEB T7 Polymerase

Prepare two 96-well PCR plate and label the plate with the following information (if operator would rather use strip tube for assay mix label only one 96-well PCR plate):

- Unique experiment number/name
- Date of experiment
- Operator identifier

**Sample master mix preparations:**
This step requires the following reagents/kit components:

- 10x Cleavage Buffer
- NEB rNTPs
- RNase Inhibitor (Murine)
- 20x Sample Loading Reagent
- FAM-7-PolyU Reporter (16 uM)
- ROX dye (50x)
- Nuclease-free water (not DEPC-treated)
- MgCl2 (1 M)

**Location**: Target-free, pre-amplification clean-room environment

1. Ensure cleaning procedures have been followed before starting. Mix the sample master mix according to *SOP Table 14: Sample master mix preparation* in a sterile nuclease-free 1.5 mL tube and mix by inverting several times.
2. Be sure to aliquot 12.6 µL os sample master mix without $MgCl_2$ to serve as a no detection control (NDC). Aliquot 12.6 µL of the sample master mix into the pre-labeled 96-well PCR plate.
3. Seal the plate with a foil seal before transferring to a designated area for working with synthetic target/patient sample DNA or RNA.
4. Keep the sample master mix plate covered on ice, on a cold block, or in a 4°C storage refrigerator until needed.
5. Sample and assay mixes can be stored at 4°C and used for up to 24 hours post-target addition in the event that experiment needs to be rerun.

**SOP Table 14:** Sample master mix preparation

| Reagent | Amount Per SIngle 14 µL Reaction (µL) | Amount Per 96 14 µL reactions + 10% Overage (µL) |
|---|---|---|
| 10x Cleavage Buffer | 1.43 | 151.008 |
| NEB rNTPs | 0.57 | 60.192 |
| RNase Inhibitor (Murine) | 0.72 | 76.032 |
| 20x Sample Loading Reagent | 0.72 | 76.032 |
| FAM-7-PolyU Reporter (16 uM) | 0.45 | 47.52 |
| ROX dye (50x) | 0.03 | 3.168 |

| | | |
|---|---|---|
| Nuclease-free water (not DEPC-treated) | 8.52 | 899.712 |
| $MgCl_2$ (1 M)* | 0.13 | 13.728 |
| Target* | 1.4 | 0 |
| **TOTAL:** | **14** | **1327.392** |

**MgCl2 (1 M)* –** Before adding in MgCl2 (1 M), take 12.6 uL of prepared solution and add to designated no detection control (NDC) position on a 96-well PCR plate.
**Target* –** To be added outside of the target-free, pre-amplification clean-room environment.
**Note –** Double master mix calculations above if two unique and separate plates of samples are to be run through workflow.

**Assay master mix preparations:**
This step requires the following reagents/kit components:

- Cas13a Working Stock
- (0.55 mg/mL)
- NEB T7 Polymerase
- 2x Assay Loading Reagent
- Nuclease-free water (not DEPC-treated)
- 1 uM Pathogen-Specific crRNA

**Location**: Target-free, pre-amplification clean-room environment

1. Ensure cleaning procedures have been followed before starting.
2. Mix the assay master mix according to *SOP Table 15: Assay master mix preparation* in a sterile nuclease-free 1.5 mL tube and mix by inverting several times.
3. Aliquot 12.6 µL of the assay master mix into a pre-labeled 96-well PCR plate or strip tube.
4. Spike-in 3.4 µL of 1 uM pathogen specific crRNAs into desired configuration.Ensure that there is at least one designated no-crRNA control composed of 12.6 µL of the assay master mix and 3.4 µL of nuclease-free water (not DEPC-treated).
5. If using a 96-well PCR plate, seal the plate with a foil seal before transferring to a designated area for working with synthetic target/patient sample DNA or RNA.
6. Keep the crRNA assay plate or strip tube on ice, on a cold block, or in a 4°C storage refrigerator until needed. This plate will now be referred to as “crRNA assay plate”.

**SOP Table 15:** Assay master mix preparation

| Reagent | Amount Per SIngle 16 µL Reaction (µL) | Amount Per 96 14 µL reactions + 10% Overage (µL) |
|---|---|---|

| | | |
|---|---|---|
| Cas13a Working Stock (0.55 mg/mL) | 0.9 | 23.76 |
| NEB T7 Polymerase | 2.5 | 66 |
| 2x Assay Loading Reagent | 8.2 | 216.48 |
| Nuclease-free water (not DEPC-treated) | 1.4 | 36.96 |
| 1 uM Pathogen-Specific crRNA | 3.4 | 0 |
| **TOTAL:** | **16** | **343.2** |

This step requires the following reagents/kit components:

- Amplified RT-PCR plate (see above, Detailed Assay Protocol Step 2: RT-PCR master mix preparations and amplification)
- Sample master mix plate

**Location**: Post-amplification, target addition BSL2+ environment

1. Ensure cleaning procedures have been followed before starting.
2. Using a microplate centrifuge PCR plate spinner, centrifuge the prepared sample master mix plate to ensure that all liquid is collected at the bottom of the well positions.
3. Add 1.4 µL of amplified RT-PCR product from the RT-PCR plate to the 96-well PCR plate containing the sample master mix. Ensure accuracy when pipetting amplified products.
4. Add 1.4 µL of nuclease-free water (not DEPC-treated) to designated NDC well(s) and to any "null" or "empty" sample wells.
5. Seal the plate with a foil seal and ensure that all edges are fully sealed.
6. Lightly vortex the plate to adequately mix sample master mix reagent and RT-PCR product.
7. Using a microplate centrifuge PCR plate spinner, centrifuge the prepared plate.
8. Keep the sample master mix plate with added RT-PCR product covered on ice, on a cold block, or in a 4°C storage refrigerator until needed. This plate will now be referred to as "sample plate with RT-PCR product".

#### 4. Priming and Loading the Fluidigm IFC

In this step the preparation and loading of the Standard BioTools Integrated Fluidic Circuit (IFC) with the necessary accessory reagents, the crRNA assay mastermix, and the sample mastermix with RT-PCR product will be detailed.

**Before starting:**

1. If using a hood or a biosafety cabinet, treat the workspace with an internal UV light for 15 minutes prior to usage. If not, proceed to Step 2.
2. Clean bench using 10% bleach, RNase AWAY® spray, and 70% ethanol
3. Clean an ice bucket or cold block with bleach, RNase AWAY® and 70% ethanol. If using an ice bucket, fill with ice.

Thaw the following reagents in ice bucket or on cold block:

- Sample plate(s) with RT-PCR product (Light sensitive)
- crRNA assay plate
- Standard BioTools Actuation Fluid
- Standard BioTools Pressure Fluid

Keep the following reagents at room temperature:

- Standard BioTools Integrated Fluidic Circuit
- Standard BioTools Control Line Fluid Syringe

**Standard BioTools IFC priming and loading**
**Location**: Post-amplification, target addition BSL2+ environment

Refer to *Figure 2: Standard BioTools Integrated Fluidic Circuit (IFC)* below:

- Ensure correct configuration of plate by looking for the **A1** marking in the top left corner of the IFC.
- Barcode is on the right side of the plate.
- There will be a black seal on the bottom of the IFC. Do not remove this until loading the IFC onto the BioMarkX.
- crRNA assay plate is to be loaded into the leftmost and rightmost columns which are denoted as **Assay inlets**.
- If running one 96-well plate through workflow, load sample plate with RT-PCR product in both left and right columns denoted as “Sample inlets”.
- If running two separate and unique 96-well plates through workflow, load sample plate with RT-PCR product 1 in the left columns denoted as “Sample inlets” and sample plate with RT-PCR product 2 in the right columns denoted as “Sample inlets”

**Figure 2 - Fluidigm Integrated Fluidics Circuit (IFC)**

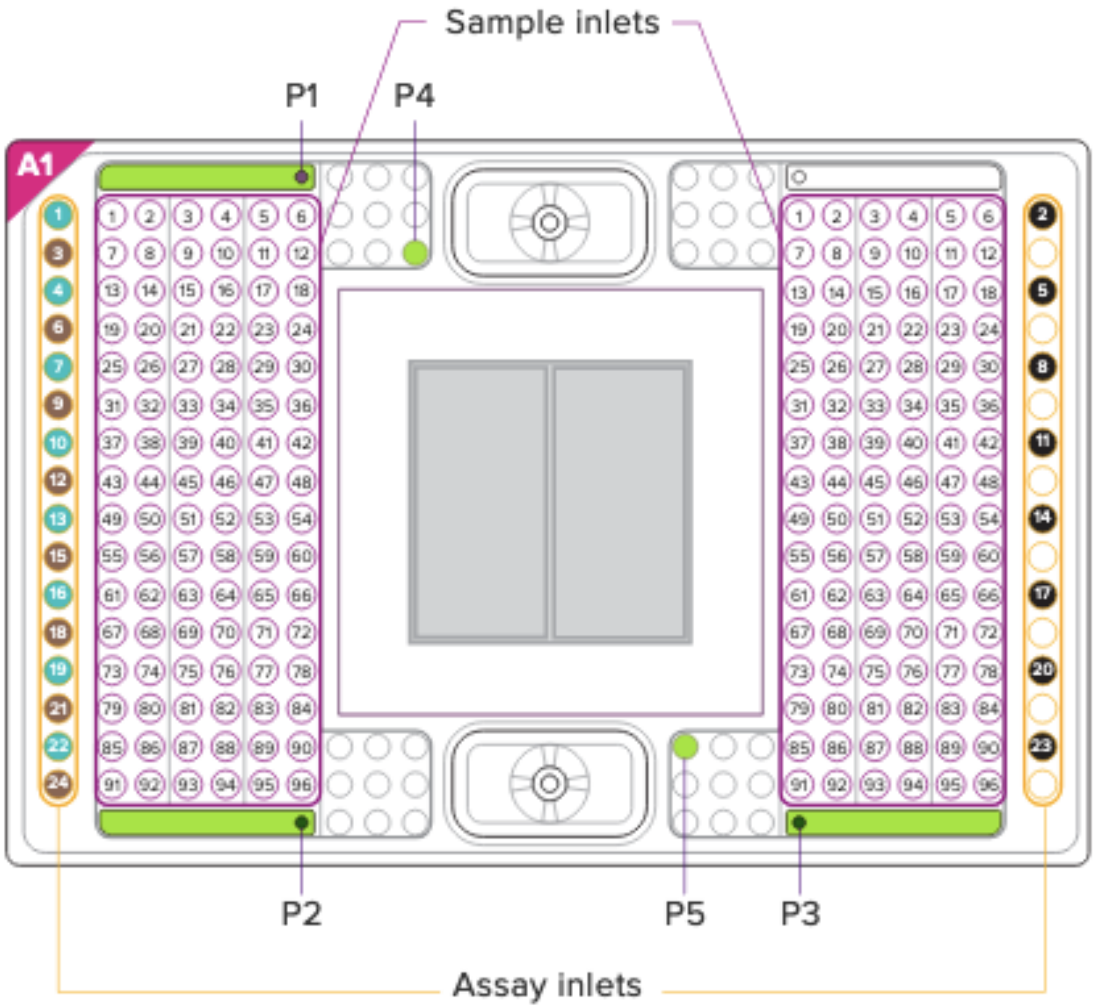

**Follow the below steps to properly load the Standard BioTools IFC:**

1. Remove the 192.24 IFC from the package. Be sure to carefully remove IFC from packaging and place it on the bench top.
2. Load the IFC control line fluid:
    a. Remove IFC control line fluid from the packaging.
    b. Repeat following steps until no bubbles remain in control line fluid syringe:
        i. Flick/tap syringe to remove bubbles.
        ii. Pull back on the plunger of the syringe
    c. Inject control line fluid syringe into the top chamber of the IFC:
        i. Insert syringe tip into the center of the top chamber of the IFC. There should be some resistance, however, the gasket with black rubber should depress allowing the tip of the syringe to enter the chamber.
        ii. Depress the plunger slowly until the syringe is empty and carefully pull the syringe out of the chamber. The gasket with black rubber should spring back into place and seal the chamber.

3. Load actuation fluid into the IFC:
    a. Pipette 150 µL of actuation fluid into the P1 well position on the IFC; see *Figure 2: Standard BioTools Integrated Fluidic Circuit (IFC)*.
    b. Ensure that no bubbles are in the P1 well position.
4. Load pressure fluid into the IFC:
    a. Pipette 150 µL of pressure fluid into the P2 and P3 well positions on the IFC; see *Figure 2: Standard BioTools Integrated Fluidic Circuit (IFC)*.
    b. Pipette 20 µL of pressure fluid into the P4 and P5 well positions on the IFC; see *Figure 2: Standard BioTools Integrated Fluidic Circuit (IFC)*.
    c. Ensure that no bubbles are in the P2, P3, P4, and P5 well positions.
5. Load crRNA assay plate:
    a. Ensure that the crRNA assay plate has been spun down before removing the foil seal.
    b. Carefully remove and discard the foil seal on the crRNA assay plate and change gloves.
    c. Using a multichannel pipette, pipette 4 µL of each column of the crRNA assay plate into the IFC assay inlet positions:
        i. Load 4 µL from well positions in column 1 of the crRNA assay plate into the left assay inlet positions. Multichannel pipette tips should be placed into the green assay inlet positions 1, 4, 7, 10, 13, 16, 19, 22; see *Figure 2: Standard BioTools Integrated Fluidic Circuit (IFC)*.
        ii. Load 4 µL from well positions in column 2 of the crRNA assay plate into the right assay inlet positions. Multichannel pipette tips should be placed into the black assay inlet positions 2, 5, 8, 11, 14, 17, 20, 23; see *Figure 2: Standard BioTools Integrated Fluidic Circuit (IFC)*.
        iii. Load 4 µL from well positions in column 3 of the crRNA assay plate into the right assay inlet positions. Make sure to not pipette into the already filled positions done in Step 5.c.ii above. Multichannel pipette tips should be placed into the brown assay inlet positions 3, 6, 9, 12, 15, 18, 21, 24; see *Figure 2: Standard BioTools Integrated Fluidic Circuit (IFC)*.
        iv. Important crRNA assay loading notes:
            1. Ensure that pipette tips are straight up and down and that pipette tips go to the bottom of the IFC well positions. It is important that the crRNA assays dispense evenly into the IFC well positions and that no bubbles are present.
            2. Do not pipette to the second stop. Only pipette to the first stop of the multichannel pipette. This is to reduce the introduction of bubbles.
            3. Ensure that all 24 assay inlet well positions are filled. Failure to do so may jeopardize the ability for the IFC data to be correctly analyzed.
            4. Note that there are 8 well positions located in the right assay inlet positions that are empty.
    d. Seal the crRNA assay plate with a foil seal and store in a 4°C storage refrigerator. This plate can be used again within 24 hours in the event that the BioMarkX run fails or needs to be re-run.
    e. Clean foil sealer and multichannel pipette following appropriate cleaning procedures.
6. Load sample plate with RT-PCR product:
    a. Ensure that the sample plate(s) with RT-PCR product have been spun down before removing the foil seal(s).
    b. Carefully remove and discard the foil seal(s) on the sample plate(s) with RT-PCR product and change gloves.
    c. Using a multichannel pipette, pipette 4 µL of each column of the sample plate(s) with RT-PCR product into the IFC sample inlet positions:
        i. Going in order from column 1-6 on the sample plate with RT-PCR product, load 4

µL into the left sample inlet positions. Note that the first pipette tip in the multichannel pipette should be placed into the inlet positions that matches the same number of the column being loaded (i.e.: column 1 samples on the sample plate with RT-PCR product should be placed in the column on the IFC starting with 1); see *Figure 2: Standard BioTools Integrated Fluidic Circuit (IFC)*.

ii. Going in order from column 7-12 on the sample plate with RT-PCR product, load 4 µL into the left sample inlet positions. Note that the first pipette tip in the multichannel pipette should be placed into the inlet positions that matches the same number of the column being loaded (i.e.: column 7 samples on the sample plate with RT-PCR product should be placed in the column on the IFC starting with 7). Note that these positions will be below columns 1-6 on the IFC; see *Figure 2: Standard BioTools Integrated Fluidic Circuit (IFC)*.

iii. Following the above two steps load the right side of the IFC.

iv. Important sample plate with RT-PCR product loading notes:
   1. If two unique and separate plates of samples are to be run through workflow, load the first plate into the left sample inlet and load the second plate into the right sample inlet.
   2. If a singular plate of samples are to be run through the workflow, load the same plate into the left and right sample inlets.
   3. Ensure that pipette tips are straight up and down and that pipette tips go to the bottom of the IFC well positions. It is important that the crRNA assays dispense evenly into the IFC well positions and that no bubbles are present.
   4. Do not pipette to the second stop. Only pipette to the first stop of the multichannel pipette. This is to reduce the introduction of bubbles.
   5. Ensure that all 24 assay inlet well positions are filled. Failure to do so may jeopardize the ability for the IFC data to be correctly analyzed.

#### 5. Running the Standard BioTools IFC on the Standard BioTools BioMarkX

In this step the Standard BioTools IFC will be placed and run on the Standard BioMarkX for both the mixing and priming of the IFC and imaging of the IFC and data will be exported to the operator. The following steps require the appropriate one hour CARMEN-BBP BioMarkX protocol.

**Before starting:**

1. Ensure that the Standard BioTools BioMarkX loading tray and Standard BioTools XA Interface Plate are appropriately cleaned following manufacturers instructions.
2. Gather the necessary equipment:
   - Standard BioTools XA Interface Plate

**Standard BioTools BioMarkX Protocol Run**
**Location**: Standard BioTools BioMarkX Instrument

1. Power on BioMarkX by pressing anywhere on the touch screen.
2. Open the door of BioMarkX by pressing the arrow on the touch screen. The IFC tray should extend out of the instrument.
3. Carefully remove the black seal on the bottom of the IFC. It is now important to not place the IFC down on any surface unless it is on the IFC tray on the BioMarkX.

4. Place IFC into the BioMarkX via the IFC tray. Ensure that the barcode is facing towards the operator.
5. Carefully place the XA Interface Plate on top of the IFC. Ensure that the XA Interface Plate barcode is facing towards the operator.
6. Close the IFC tray by pressing on the arrow on the touch screen. The BioMarkX will scan both the IFC and the XA Interface Plate.
7. Operator should be presented with a set of preset protocols that are compatible with the 192.24 IFC and the XA Interface Plate. Select protocol for CARMEN-BBP.
8. Wait until protocol is complete (~1 hour and 45 minutes) to collect data.

**Standard BioTools BioMarkX Data Exporting**
**Location**: Standard BioTools BioMarkX Instrument

1. Post-completion of BioMarkX CARMEN-BBP run, the operator should be presented with an option to export the run data. To do so, the operator should input a USB drive into the USB port at the front of the BioMarkX instrument and select the export button.
2. Once the export is complete, the operator will be prompted with an arrow button on the touch screen of the BioMarkX. Operator should select this.
3. The IFC tray on the BioMarkX should open and the IFC should be placed into an appropriate biohazardous waste disposal stream. The XA Interface Plate should be cleaned and stored following manufacturer's instructions.
4. Users will then take exported material, in the form of a .bml file, and upload and analyze data via the Standard BioTools Real-time PCR Analysis Software. Once the .bml file has been analyzed, the user can download the raw data file needed in the form of a .csv. This can be done by:
    a. Inserting the USB drive containing the exported BioMarkX .bml file into the computer with the Standard BioTools Real-time PCR Analysis Software.
    b. Open Standard BioTools Real-time PCR Analysis Software and select “Open a Chip Run” under “Quick Tasks”.
    c. Navigate to and select the exported BioMarkX .bml file.
    d. The Standard BioTools Real-time PCR Analysis Software should present back to the user a “Chip Run Summary”. If so, select “File” in the top left corner of the program and select “export” to the USB drive. The USB drive should now have the necessary .csv file for the CARMEN-BBP Software Pipeline.

#### 6. Data Analysis Using CARMEN-BBP Software Pipeline

Data that is retrieved from the Standard BioTools BioMarkX in the .csv format will be analyzed via a locally hosted software run in the command line on the operator’s computer. Please follow installation and user instructions found here.

To run the above software, operators will need to provide two data files.

1. Standard BioTools BioMarkX raw data file (.csv format) - Retrieved from BioMarkX instrument as described in *Step 5 - Standard BioTools BioMarkX Data Exporting*. This data file will be called “Results_all.csv” in the exported folder.
2. Sample/Assay assignment sheet (.xlsx format) - Created by operator following instructions found in software instructions.

#### 7. Interpretation of Results and Reporting

Details regarding interpretation of results and important control information is provided.

**Interpretation of Control Results**

CARMEN utilizes four process controls and two internal controls to monitor several different aspects of the assay; including extraction, RT-PCR, and detection. The CARMEN software uses these controls to test for proper performance of each assay and to calculate a threshold for positivity/negativity of each assay, which is then applied to each sample.

See *SOP Table 16: Expected performance of controls* below for a summary of the controls used with CARMEN and their expected performance. See *Appendix 2: Detailed interpretation of control results* for a more detailed discussion of the controls used with CARMEN.

**SOP Table 16:** Expected performance of controls

| Control Type | Use-Case | Input into Pipeline | Expected RNase P Result | Expected no-crRNA Result | Expected Pathogen Target Result |
|---|---|---|---|---|---|
| **Extraction negative Control (EC)** | Failure in extraction due to contamination | Nuclease-Free Water (not DEPC-treated) into four dedicated extraction well positions (A1, A12, H1, and H12) | Negative | Negative | All negative* |
| **No Template negative Control (NTC)** | RT-PCR reagent and/or environmental contamination | Nuclease-Free Water (not DEPC-treated) into dedicated RT-PCR well position | Negative | Negative | All negative* |
| **Combined Positive Control (CPC)** | Detection/RT-PCR reagent failure including primer, crRNA, and CRISPR-Cas13 integrity | Combined positive control input into dedicated RT-PCR well position | Positive | Negative | All positive* |
| **Negative Detection Control (NDC)** | Sample master mix and/or environmen tal | Nuclease-Free Water (not DEPC-treated) into dedicated well position | Negative | Negative | All negative* |

| | | | | | |
|---|---|---|---|---|---|
| | contamination | without Magnesium Chloride in the sample master mix | | | |
| **No-crRNA Control** | Assay master mix and/or environmental contamination | Nuclease-Free Water (not DEPC-treated) into dedicated assay master mix well position | Negative | Negative | Negative |
| **Internal Sample Controls** | RNase P | Patient samples | Positive** **†** | Negative** | N/A |

(*) If results for any of the four process controls do not exhibit expected performance for any of the pathogen targets, *results are invalidated only for that pathogen target assay*. (**) denotes internal controls. If these do not exhibit expected performance, *only the individual sample results are invalidated*.

(**†**) If RNase P is negative for a given sample, but another pathogen target for the same sample produces a signal, the result is valid. This is due to competitive amplification, a high pathogen titer may cause a low or absent RNase P signal.

If results for the four process controls do not exhibit expected performance for either RNase P or no crRNA controls, the *entire **CARMEN run is invalidated*** and must be redone from the biosample/extraction step.

**Interpretation of Patient Sample Results**

Detection of a pathogen target in a sample depends on the pathogen target specific sample signal compared to the NTC. The determination of results from the CARMEN is conducted automatically by the CARMEN software.

A pathogen target is **detected** in a sample if the sample signal is greater than the average of the NTC plus three times the standard deviation of the NTC per the respective assay. Therefore, a unique threshold is calculated per each target assay and is applied to all tested samples. Otherwise, it is **not detected**. Multiple viruses may be detected in a single sample.

**Pathogen Gene Targets**

If all process controls exhibit the expected results as defined above, the results for the samples can be interpreted as follows (see *SOP Table 17: Sample results summary*):

1. If one or more of the pathogen targets for a given sample generates a signal above the calculated threshold, the sample is considered detected for those viruses.
    a. Note: RNase P reaction may or may not be positive as described above, but the pathogen target results are still valid if at least one of them is above the threshold.
2. If all pathogen targets for a given sample *do not* generate a signal above the calculated threshold

but the RNase P for that sample *does* exceed the calculated threshold, the sample is considered not detected for all pathogen targets.

3. If the signals of the pathogen targets for a given sample *do not* meet or exceed the calculated threshold and the RNase P for the same sample *does not* meet or exceed the calculated threshold, the results *for that sample* are invalid.
    a. Note: In the case of invalid results, repeat CARMEN runs for the invalid sample(s) from the biosample/extraction step. If the sample(s) remains invalid upon retest, collection of a new specimen and subsequent testing should be considered

**SOP Table 17:** Sample results summary

| Sample Result | Definition of Result |
|---|---|
| Positive | Positive result. The pathogen DNA/RNA is present in the sample, above the calculated threshold for positivity. |
| Negative | Negative result. The signal falls below the calculated threshold for positivity. |
| Invalid | Invalid assay result. This is usually due to an issue with one or more control (see above). |

#### V. Appendix 1: CARMEN-BBP Primers, crRNA, and Reporter Sequences

All materials sourced from IDT.

**Table A1: CARMEN-BBP1 primers and crRNA sequences**

| Number | Pathogen Name | Primer/crRNA | Oligonucleotide Sequence (5'-3') | Gene Targeted |
|---|---|---|---|---|
| 1 | Crimean Congo Hemorrhagic Fever Virus | Forward Primer | gaaatTAATACGACTCACTATAgggCAATGAGTGCAGGCAAGATTG | L segment |
| | | Reverse Primer | GCTTTCTATGCCATCACAAATTATG | |
| | | crRNA | GAUUUAGACUACCCCAAAAACGAAGGGGACUAAAACCGGCUCUACUUCAGAAUCUUCUUGAAGA | |
| 2 | Zaire ebolavirus | Forward Primer | gaaatTAATACGACTCACTATAgggTTGCCTACTGCTCCTCCTG | Matrix protein |
| | | Reverse Primer | GCTGTTGCCACCCCTAG | |
| | | crRNA | gauUUAGACUACCCCAAAAACGAAGGGGACUaaaacUUUGACCUGGCAGGGUAUAUGGCCUCCA | |
| 3 | Human Immunodeficiency Virus 1 | Forward Primer | gaaatTAATACGACTCACTATAgggCTTTGGCAACGACCCCTC | Gag polyprotein |
| | | Reverse Primer | CCCCTATCATTTTTGGTTTCCATT | |
| | | crRNA | GAUUUAGACUACCCCAAAAACGAA | |

| | | | | |
|---|---|---|---|---|
| | | | GGGGACUAAAA CAUACUGUAUCA UCUGCUCCUGU AUCUAA | |
| 4 | Human Immunodeficiency Virus 2 | Forward Primer | gaaatTAATACGA CTCACTATAgggT GCACGCAGAAG AGAAAGTG | Gag polyprotein |
| | | Reverse Primer | GCCCCGAACTTC TTTTCCTC | |
| | | crRNA | gauUUAGACUAC CCCAAAAACGAA GGGGACUaaaac UUUAGGGUUCG GGGACUCAGCG GCACAU | |
| 5 | Lassa Virus | Forward Primer | gaaatTAATACGA CTCACTATAgggC TCAACCATTTTG TGCATATGCC | L segment |
| | | Reverse Primer | GATCACAGTAAG TGGGGCC | |
| | | crRNA-1 | gauUUAGACUAC CCCAAAAACGAA GGGGACUaaaac AAGGACUUGCAA GCUGACAUAAAA GGGA | |
| | | crRNA-2 | gauUUAGACUAC CCCAAAAACGAA GGGGACUaaaac GUAGAUUUACAA GCUGAUUUAAAA GGCA | |
| 6 | Marburg Virus | Forward Primer | gaaatTAATACGA CTCACTATAgggC TGAGGATCTTGC TTGGAGTTG | Membrane associated protein |
| | | Reverse Primer | GCATATGAACAA TGGATCTAAATC CAG | |

| | | | | |
|---|---|---|---|---|
| | | crRNA | gauUUAGACUACCCCAAAAACGAAGGGGACUaaaacUCAAUGAUUGCUGUAAUUCUUGAUCCUU | |
| 7 | West-Nile Virus | Forward Primer | gaaatTAATACGACTCACTATAgggCTTCGCAAGAGGTGGACAG | Nonstructural protein NS2A |
| | | Reverse Primer | CAGTGTAAGTAATGCCCCCAAAC | |
| | | crRNA | gauUUAGACUACCCCAAAAACGAAGGGGACUaaaacCUAGCAGAGCAAUCAGUAUAGCUGGCAU | |
| 8 | Yellow Fever Virus | Forward Primer | gaaatTAATACGACTCACTATAgggCCGTGCCATCTGGTACATG | RNA-dependent RNA polymerase |
| | | Reverse Primer | GTGTCCCATCCAGCTGTG | |
| | | crRNA | gauUUAGACUACCCCAAAAACGAAGGGGACUaaaacUGCAGCCAGGUCCCUGAUCACGUAGCCU | |
| 9 | Ribonuclease P | Forward Primer | GAAATTAATACGACTCACTATAGGGTTGATGAGCTGGAGCCA | |
| | | Reverse Primer | ATGTGGATGGCTGAGTTGTT | |
| | | crRNA | GAUUUAGACUACCCCAAAAACGAAGGGGACUAAAACUCCGAGUCAGUGGCUCCCGUGUGUCGGU | |

**Table A2: CARMEN-BBP2 primers and crRNA sequences**

| Number | Pathogen Name | Primer/crRNA | Oligonucleotide Sequence (5'-3') | Gene Targeted |
|---|---|---|---|---|
| 1 | Chikungunya Virus | Forward Primer | gaaatTAATACGACTCACTATAgggGAAGGGCTCGAGGTCACG | Structural polyprotein |
| 1 | Chikungunya Virus | Reverse Primer | GTGGCCATGGGCTGTAC | Structural polyprotein |
| 1 | Chikungunya Virus | crRNA | GAUUUAGACUACCCCAAAAACGAAGGGGACUAAAACGGAUAACUGCGGCCAAUACUUGUACGGC | Structural polyprotein |
| 2 | Dengue Virus | Forward Primer | gaaatTAATACGACTCACTATAgggCTGAAACGCGCGAGAAAC | |
| 2 | Dengue Virus | Reverse Primer-1 | CACCATCCGTAAGGGTCCTTT | |
| 2 | Dengue Virus | Reverse Primer-2 | AACCAATTTCATTGGTCCCTG | |
| 2 | Dengue Virus | Reverse Primer-3 | CACCAATTTCATGGGTCCTTG | |
| 2 | Dengue Virus | Reverse Primer-4 | GAACAGTTTTAATGGTCCTCG | |
| 2 | Dengue Virus | crRNA-1 | GAUUUAGACUACCCCAAAAACGAAGGGGACUAAAACUUUGAGAAUCUCUUCGCCAACUGUGAAA | |
| 2 | Dengue Virus | crRNA-2 | GAUUUAGACUACCCCAAAAACGAAGGGGACUAAAACAGUGAGAAUCUCUUUGUCAACUGUUGUA | |

| | | | | |
|---|---|---|---|---|
| 3 | Rift Valley Fever Virus | Forward Primer | gaaatTAATACGACTCACTATAgggGCCGGAAGATATATTATTCCTATTTCC | L segment |
| | | Reverse Primer | GATGATGTCTATTTCTCCCTTGTTG | |
| | | crRNA | gauUUAGACUACCCCAAAAACGAAGGGGACUaaaacCUAUAGAAUCCAGUGAUUCUAAUUCCUC | |
| 4 | Zika Virus | Forward Primer | gaaatTAATACGACTCACTATAgggTGGAACTCATGAAAAGAGGAGATC | Nonstructural protein NS3 |
| | | Reverse Primer | CGTGCCATCAAAGCACCATC | |
| | | crRNA | gauUUAGACUACCCCAAAAACGAAGGGGACUaaaacGUAGGUUAUUCCGGCAGAUGCAACCUGA | |
| 5 | Measles morbillivirus | Forward Primer | gaaatTAATACGACTCACTATAgggGAAAAACGTAGGGTCCAAGTG | |
| | | Reverse Primer | CTAGGTGAACTTCAGGGTATAAGATC | |
| | | crRNA | GAUUUAGACUACCCCAAAAACGAAGGGGACUAAAACGUUGACAGAUAGCGAGUCCAUAACGGGA | |
| 6 | Rabies lyssavirus | Forward Primer | gaaatTAATACGACTCACTATAgggCACAAAATGTGTG | Nucleoprotein N |

| | | | | |
|---|---|---|---|---|
| | | | CTAACTGGAG | |
| | | Reverse Primer | CTTCCTCAAAGT TCTTGTGGAAG | |
| | | crRNA | GAUUUAGACUAC CCCAAAAACGAA GGGGACUAAAA CUGCUGAAUAG AGAUGCUCAAUC CGGGAG | |
| 7 | Hepatitis C Virus | Forward Primer | gaaatTAATACGA CTCACTATAgggG CAATTTGGGTAA GGTCATCG | polyprotein (POLY) |
| | | Reverse Primer-1 | GGGCAGATTCCC TGTTGC | |
| | | Reverse Primer-2 | CGGGCAGATTCC CTGTTG | |
| | | crRNA | GAUUUAGACUAC CCCAAAAACGAA GGGGACUAAAA CGUACCCCAUG AGGUCGGCGAA GCCGCAC | |
| 8 | Hantaan orthohantavirus | Forward Primer | gaaatTAATACGA CTCACTATAgggC TGAGTTCCCAGG TAGTTTCAG | Segment M |
| | | Reverse Primer | GTTGCCATTACT TTCTTGTAACCT G | |
| | | crRNA | GAUUUAGACUAC CCCAAAAACGAA GGGGACUAAAA CAAUAGGAGUA GUAGCAAAGUU GCAGUGC | |
| 9 | Nipah henipavirus | Forward Primer | gaaatTAATACGA CTCACTATAgggG CCAAAAAACATG GACTATAGCAAC | |

| | | | |
|---|---|---|---|
| | | Reverse Primer | GACAATTGCAGC AATCCTCG |
| | | crRNA | GAUUUAGACUAC CCCAAAAACGAA GGGGACUAAAA CUUUUAUUCUU GAGUGCCUAUG AGACAAA |
| 10 | Ribonuclease P | Forward Primer | GAAATTAATACG ACTCACTATAGG GTTGATGAGCTG GAGCCA |
| | | Reverse Primer | ATGTGGATGGCT GAGTTGTT |
| | | crRNA | GAUUUAGACUAC CCCAAAAACGAA GGGGACUAAAA CUCCGAGUCAG UGGCUCCCGUG UGUCGGU |

**Table A3: CARMEN-STI primers and crRNA sequences**

| Number | Pathogen Name | Primer/crRNA | Oligonucleotide Sequence (5'-3') | Gene Targeted |
|---|---|---|---|---|
| 1 | Monkeypox Virus | Forward Primer | gaaatTAATACGA CTCACTATAgggC TCCTTTTCGACC TCAATAACTTTA G | A26L |
| | | Reverse Primer | GCCAGGGAATCA GATAAAAATGAT AG | |
| | | crRNA | GAUUUAGACUAC CCCAAAAACGAA GGGGACUAAAA CCUACAAGAGAG AGCUUGAUGAG ACAACG | |
| 2 | Hepatitis B Virus | Forward Primer | gaaatTAATACGA CTCACTATAgggG GACGAATCTTTC | Large S envelope protein |

| | | | | |
|---|---|---|---|---|
| | | | TGTTCCCA | |
| | | Reverse Primer | GAGTTGGCTCCG AATGCAG | |
| | | crRNA | GAUUUAGACUAC CCCAAAAACGAA GGGGACUAAAA CCCAACUGAUGA UCGGGAAAGAA UCCCAG | |
| 3 | Herpes Simplex Virus Type 2 | Forward Primer | gaaatTAATACGA CTCACTATAgggG TCATGTGTATTTT TGAACATCGG | Glycoprotein B (UL27) |
| | | Reverse Primer | GACGAAGACGA GCTCTAAGG | |
| | | crRNA | GAUUUAGACUAC CCCAAAAACGAA GGGGACUAAAA CGAGGGGAGCU GGGCUUGUGUA UAAAUAA | |
| 4 | Chlamydia trachomatis | Forward Primer | gaaatTAATACGA CTCACTATAgggG TTCAGATTGAAC GCTGGC | 16S |
| | | Reverse Primer | CGCCACTAAACA ATTGCCG | |
| | | crRNA | GAUUUAGACUAC CCCAAAAACGAA GGGGACUAAAA CACAAUUGCUCC GUUCGACUUGC AUGCCU | |
| 5 | Treponema pallidum subsp. pallidum | Forward Primer | gaaatTAATACGACT CACTATAgggCCTA TAAGGCAAAAAGG GATAAGAC | PolA |
| | | Reverse Primer | CCAGTGCACACAG GATTTCT | |

| | | | | |
|---|---|---|---|---|
| | | crRNA | GAUUUAGACUACCCCAAAAACGAAGGGGACUAAAACAAGGGGAAUUUGCGCAUAAAGCUCUGCA | |
| 6 | Trichomonas vaginalis | Forward Primer | gaaatTAATACGACTCACTATAgggCGTTGAATACGTCCCTGCC | 18S |
| | | Reverse Primer | CGGTGATTTCACCGAGTCA | |
| | | crRNA | GAUUUAGACUACCCCAAAAACGAAGGGGACUAAAACGGAGCGACGGGCGGUGUGUACAAAGGGC | |
| 7 | Ribonuclease P | Forward Primer | GAAATTAATACGACTCACTATAGGGTTGATGAGCTGGAGCCA | |
| | | Reverse Primer | ATGTGGATGGCTGAGTTGTT | |
| | | crRNA | GAUUUAGACUACCCCAAAAACGAAGGGGACUAAAACUCCGAGUCAGUGGCUCCCGUGUGUCGGU | |

#### VI. Appendix 2: Detailed interpretation of control results

Observed unexpected behavior of process controls can invalidate either an entire run, or results from specific pathogen target(s):

**1. Extraction negative control (EC)**- *Extraction process control*

The EC is used as an non-template extraction control to serve as an indicator for possible cross-contamination between extraction wells during nucleic acid isolation. The EC consists of nuclease-free (not DEPC-treated) water in addition to the necessary reagents needed for proper extraction. The EC should be carried through the entire process, from extraction to detection.

- If the EC is non-detectable for all targets this confirms the success of the extraction step and the run is valid with respect to the extraction step.
- If the EC is detected in any of the pathogen targets in a given run, this may be an indication of possible cross-contamination during extraction, RT-PCR or detection reaction set-up.
    - In this case, *only the results from samples for the EC detected viral target are invalid*, and need to be repeated.

**2. No Template Control (NTC)** - *RT-PCR master mix control*

The NTC is used as a control for the integrity of RT-PCR reagents and cross-contamination during the amplification process. To observe a detected result in the NTC reaction, a signal from a viral target must be above three standard deviations from the mean of all the NTC signals from all targets and RNase P (with the exception of the NTC signal from the no-crRNA control (see below.)) The NTC consists of nuclease-free (not DEPC-treated) water in RT-PCR reaction well(s) instead of sample input.

- If the NTC is not detected in the RNase P target, not detected in the no-crRNA control, and not detected in any pathogen targets, this indicates that the RT-PCR amplification and detection reagents are not contaminated, and the run is valid with respect to the RT-PCR.
- If the NTC is detected in:
    - either the RNase P target or the no-crRNA target, in a given run, *this will invalidate the entire run*.
    - any of the pathogen targets, *only the results from samples for the NTC detected* pathogen *target are invalid*. This might suggest possible cross-contamination during the RT-PCR or detection reaction set-up steps. Extraction, RT-PCR, and detection need to be repeated for these pathogen targets.

**3. Combined Positive Control (CPC)** - *Positive detection control*

The CPC consists of all targets on each CARMEN-BBP panel (CARMEN-BBP1, 2, and 3). The CPC consists of *in vitro* transcribed gene fragments of all targeted pathogens and RNase P and is combined as directed in *Appendix 3: In vitro transcription of CARMEN-BBP gene fragments for LoD testing, contrived sample evaluation, and CPC*. The CPC serves as a control for the integrity of the RT-PCR reagents and primer sets, and to ensure amplification and detection were properly performed.

The CPC is run alongside each batch of clinical samples after extraction and is to be added into the RT-PCR master mix. The CPC should produce a detected result for all pathogen and RNase P targets in a given run. To observe a detected result in the CPC reaction, a signal from a viral target must be above three standard deviations from the mean of all the NTC signals from all viral targets and RNase P (with the exception of the NTC signal from the no-crRNA control (see below.))

- If the CPC is detected for each of the pathogen and RNase P targets, this indicates that the RT-PCR and detection steps were properly performed and the run is valid with respect to the RT-PCR and detection steps.
- If the CPC is not detected for any pathogen or RNaseP target, this indicates that there is a problem with the primer mixes or reagents used in the RT-PCR amplification, or that the detection step failed.
    - In this case, only the results from samples for the not detected CPC pathogen target are invalid and need to be repeated.
- The no-crRNA detection control (see below) should always result in a no detection outcome for the CPC.

**4. Negative Detection Control (NDC)** - *Sample master mix control*

The NDC is used to test the validity and integrity of the Sample Master Mix reagents in the absence of $MgCl_2$. The NDC consists of using nuclease-free water (not DEPC-treated) into a dedicated sample master mix well position instead of an amplified product. Additionally, this dedicated sample master mix is created without the addition of Magnesium Chloride ($MgCl_2$), which is needed to facilitate CRISPR-Cas13 enzymatic activation.

- If the NDC produces no detection for all pathogen and RNase P targets, this suggests that the sample master mix was properly set-up, and therefore the run is valid with respect to the sample master mix.
- If the NDC produces a detected result for any target according to the CARMEN-BBP software, that indicates cross-contamination may have occurred during the detection set-up.
    - In this case, only the results from samples for the NDC detected pathogen target are invalid, and need to be repeated.

**6. Internal Controls** - evaluated only after all process controls have exhibited expected behavior

**a. No crRNA Control (no-crRNA)** - *Assay master mix control*

The no-crRNA control is used to test the validity of the assay master mix predicates, and as a control for crRNA cross-contamination during the detection set-up. The no-crRNA control consists of the assay master mix predicate with nuclease-free water (not DEPC-treated) instead of crRNA and should produce no detection for all samples and controls.

- If the no-crRNA control is detected for:
    - any sample: results from that sample alone are invalid and that sample should be rerun from the biosample/extraction step.
    - any control: cross-contamination during the detection set-up likely occurred and the run is invalid.
- If the no-crRNA control produces a not detected result for all controls and samples, then the run is valid.

**b. RNase P control (RNase P)**

The RNase P control is used as an internal amplification control and a verification of adequate sample collection. The RNase P control consists of a primer pair and crRNA set that targets the Human RNase P gene (see *Appendix 1: CARMEN-BBP Primers, crRNA, and Reporter Sequences).* The RNase P control is run alongside all the other pathogen targets and the no-crRNA control.

- If the RNase P control is detected for a given sample, even if no other pathogen targets are detected, then results from that sample are valid.

- <u>If the RNase P control is not detected</u> for a given sample:
  - and no other pathogen targets are detected, then the results from that sample are invalid.
  - but at least one other pathogen target is detected, these sample results are not invalidated. This is due to competitive amplification where a high pathogen titer may cause a low or absent RNase P signal.

#### VII. Appendix 3: *In vitro* transcription of CARMEN-BBP gene fragments for LoD testing, contrived sample evaluation, and CPC

DNA targets were ordered as dsDNA fragments, gBlocks, from Integrated DNA Technologies (IDT) *in vitro* transcribed using the HiScribe T7 High Yield RNA Synthesis Kit (New England Biolabs). Transcriptions were performed according to the manufacturer's recommendations with a reaction volume of 20 µL (>300 base pairs) or 30 µL (<300 base pairs) that was incubated overnight at 37 °C. The transcribed RNA products were purified using RNAClean XP beads (Beckman Coulter) and quantified using NanoDrop One (Thermo Scientific). For experimental validation, the RNA was serially diluted from $10^8$ down to $10^1$ cp/µL and used as input into CARMEN-BBP. CPC was made by pooling equal quantities of all *in vitro* transcribed RNA targets per CARMEN-BBP1, 2, 3 to 1,000,000 copies/µL (see *Appendix Table A3: Sequences of synthetic targets for CARMEN-BBP1, 2, 3*). In the case that the BBP-CARMEN assay was targeting DNA pathogens, like in CARMEN-BBP3, DNA gBlocks of these pathogen targets were diluted to 1,000,000 copies/µL without processing through *in vitro* transcription and were pooled together to create a CARMEN-BBP3 CPC.

**Appendix Table A4:** Sequences of synthetic targets for CARMEN-BBP1

| Number | Pathogen | Oligonucleotide Sequence (5'-3') |
|---|---|---|
| 1 | Crimean Congo Hemorrhagic fever Virus | gaaatTAATACGACTCACTATAgggTCAGGCAGAGCAATGAGTGCAGGCAAGATTGGTAATGTCCTTTCCTATGCACATAACCTTTATTTGAGTAAGTCTAGCCTAAATATGACCTCTGAGGACATCTCACAGCTTCTGATCGAGATCAAGAGATTGTATGCTCTACAAGAAGATTCTGAAGTAGAGCCGATAGCCATAATTTGTGATGGCATAGAAAGCAATATGA |
| 2 | Zaire ebolavirus | gaaatTAATACGACTCACTATAgggCGGGTTATATTGCCTACTGCTCCTCCTGAATATATGGAGGCCATATACCCTGCCAGGTCAAATTCAACAATTGCTAGGGGTGGCAACAGCAATACAGGGG |
| 3 | Human Immunodeficiency Virus 1 | gaaatTAATACGACTCACTATAgggCTTCCCTCAAATCACTCTTTGGCAACGACCCCTCGTCACAATAAAGATAGGGGGACAGCTAAAGGAAGCTCTATTAGATACAGGAGCAGATGATACAGTATTAGAAGAAATAAATTTGCCAGGAAAATGGAAACCAAAAATGATAGGGGGAATTGGAGGTTT |

| | | |
|---|---|---|
| 4 | Human Immunodeficiency Virus 2 | gaaatTAATACGACTCACTATAgggGTGCTTGCACGCAGAAGAGAAAGTG<br>AAAGATACTGAAGAAGCAAAAAAGATAGTACAGAGACATCTAGCGGC<br>AGAAACAGGAACTGCAGAGAAAATGCCAAATACAAGTAGACCAACAG<br>CACCACCTAGCGGGAAAGGAGGAAACTACCCCGTGCAACAAGTAGC<br>CGGCAACTATACCCATGTGCCGCTGAGTCCCCGAACCCTAAATGCTT<br>GGGTAAAATTAGTAGAGGAAAAGAAGTTCGGGGCAGAAG |
| 5 | Lassa Virus | gaaatTAATACGACTCACTATAgggTGCAGGATACCCTGTCCCATATCAA<br>GGATTGAACTAATGTGTGAGGGGACTGTGCCAATTTGAAAGTTGGAA<br>TTAAAAAAGTCTTCGGTCATTGATTGAGTTGTCTTCTTCTTCAATCCAA<br>GTTGGGATTTGATGTATGATTTCATCATTGCTGATACAACATTAAATG<br>GGATCTCAACCATTTTGTGCATATGCCATGTTAATAATGTTGATAGGT<br>AATCTTTACCTTTTAAATCAGCTTGTAGGTCATCAGAAAGTAAGATTAG<br>GTTTTGAATTATTGTTAAAAACAAGAAAGGACACATCATAGGGCCCCA<br>CTTACTGTGATCCATACTATATGATACATGTGCCGATGAGACATTCAG<br>CTTCATTGACAAAATGGCATTGTCAAACTCCTTCTCATTGTTTAGACA<br>GCTGCCTGCTAATTGTGATGTGAGCGCTTCAAAGTAATCCTCAATGA<br>GCCTTGTGAACATCTTTGTCCTTAAATCTCCGATGTACAACTC |
| 6 | Marburg Virus | gaaatTAATACGACTCACTATAgggCGTTCTTGGCTCTGAGGATCTTGCT<br>TGGAGTTGCTTTGAAGGATCAAGAATTACAGCAATCATTGATTCCTGG<br>ATTTAGATCCATTGTTCATATGCTTTCAGAAT |
| 7 | West-Nile Virus | gaaatTAATACGACTCACTATAgggGTCCTTCGCAAGAGGTGGACAGCC<br>AAGATCAGCATGCCAGCTATACTGATTGCTCTGCTAGTCCTGGTGTTT<br>GGGGGCATTACTTACACTGATGGGGGGG |

| | | |
|---|---|---|
| 8 | Yellow Fever Virus | gaaatTAATACGACTCACTATAgggGAAGCCGTGCCATCTGGTACATGTG<br>GCTGGGAGCCCGATATCTTGAGTTTGAAGCCCTGGGATTCCTGAATG<br>AGGACCACTGGGCATCCAGGGAGAACTCAGGGGGAGGGGTTGAAGG<br>TATCGGTCTACAATATCTAGGCTACGTGATCAGGGACCTGGCTGCAC<br>TAGAGGGTGGTGGATTCTATGCGGATGACACAGCTGGATGGGACACA<br>CGCATAAC |
| 9 | RNaseP | GAAATTAATACGACTCACTATAGGGTCCTTGCAGGTGGCTGCCAATAC<br>CTCCACCGTGGAGCTTGTTGATGAGCTGGAGCCAGAGACCGACACAC<br>GGGAGCCACTGACTCGGATCCGCAACAACTCAGCCATCCACATCCGA<br>GTCTTCAGGGTCACACCCAAGTAATTGAAAAGACACTCCTCCACTTAT<br>CCCCTCCGTGATATGGCTCTTCGCATGCTGAGTACTGGACCTCGGAC<br>CAGAGCCATGTAAGAAAAGGCCTGTTCCCTGGAAGCCCAAAGGACTC<br>TGCATTGAGGGTGGGGGTAATTGTCTCTTGGTGGCCCAGTTAGTGGG<br>CCTTCCTGA |

**Appendix Table A5:** Sequences of synthetic targets for CARMEN-BBP2

| Number | Pathogen | Oligonucleotide Sequence (5'-3') |
|---|---|---|
| 1 | Chikungunya | gaaatTAATACGACTCACTATAgggCCGTGCCGACTGAAGGGCTCGA<br>GGTCACGTGGGGTAACAACGAGCCGTATAAGTATTGGCCGCAGTT<br>ATCTACAAACGGTACAGCCCATGGCCACCCGCATGAGATAATTC |
| 2 | Dengue Virus | gaaatTAATACGACTCACTATAgggCTTTCAATATGCTGAAACGCGCG<br>AGAAACCGCGTGTCAACTGTTTCACAGTTGGCGAAGAGATTCTCAA<br>AAGGATTGCTCTCAGGCCAAGGACCCATGAAATTGGTGATGGCTT<br>TCATAGCGTTCC |
| | | gaaatTAATACGACTCACTATAgggCTTTCAATATGCTGAAACGCGAGA<br>GAAACCGCGTGTCAACTGTGCAACAGCTGACAAAGAGATTCTCACTT<br>GGAATGCTGCAAGGACGCGGACCATTAAAACTGTTCATGGCCCTTG<br>TGGCGTTCC |
| | | gaaatTAATACGACTCACTATAgggCTATCAATATGCTGAAACGCGTGA |

| | | |
|---|---|---|
| | | GAAACCGTGTGTCAACTGGATCACAGTTGGCGAAGAGATTCTCAAA<br>AGGACTGCTGAACGGCCAGGGACCAATGAAATTGGTTATGGCGTT<br>CATAGCTTTCCT |
| | | gaaatTAATACGACTCACTATAgggCTTTCAATATGCTGAAACGCGAGA<br>GAAACCGCGTATCAACCCCTCAAGGGTTGGTGAAGAGATTCTCAAC<br>CGGACTTTTTTCTGGGAAAGGACCCTTACGGATGGTGCTAGCATTC<br>ATCACGTTTTTGCG |
| 3 | Rift Valley Fever Virus | gaaatTAATACGACTCACTATAgggTATGGAGCCGGAAGATATATTATT<br>CCTATTTCCGAACATTGAGGAATTAGAATCACTGGATTCTATAGTTT<br>ACAACAAGGGAGAAATAGACATCATCCCAAGAG |
| 4 | Zika Virus | gaaatTAATACGACTCACTATAgggACCTTTGTGGAACTCATGAAAAG<br>AGGAGATCTTCCTGTTTGGCTGGCCTATCAGGTTGCATCTGCCGG<br>AATAACCTACACAGATAGAAGATGGTGCTTTGATGGCACGACCAAC |
| 5 | Measles morbillivirus | gaaatTAATACGACTCACTATAgggGACATCAGAATTAAGAAAAACGT<br>AGGGTCCAAGTGGTTCCTCGTTATGGACTCGCTATCTGTCAACCA<br>GATCTTATACCCTGAAGTTCACCTAGATAGCCCGATAG |
| 6 | Rabies lyssavirus | gaaatTAATACGACTCACTATAgggTCTGATGACAACTCACAAAATGT<br>GTGCTAACTGGAGTACTATACCGAACTTCAGATTCCTGGCCGGAA<br>CCTACGACATGTTTTTCTCCCGGATTGAGCATCTATATTCAGCAAT<br>CAGAGTGGGTACAGTTGTCACTGCTTATGAAGACTGCTCAGGGCT<br>GGTATCATTTACTGGGTTCATAAAGCAGATAAATCTCACCGCAAGG<br>GAAGCAATACTATATTTCTTCCACAAGAACTTTGAGGAAGAGATAA |

| | | |
|---|---|---|
| | | GAA |
| 7 | Nipah henipavirus | gaaatTAATACGACTCACTATAgggGAGATACGAATCAATGGCTATATTTGCTGAACGTCTGGATGAGATATACGGTTTACCTGGATTTTTTAATTGGATGCACAAACGACTAGAAAGATCTGTTATCTATGTTGCAGACCCTAATTGCCCCCCTAATATTGACAAACATATGGAACTAGAAAAAACTCCTGAAGATGATATATTCATTCATTATCCTAAAGGCGGTATTGAAGGATATAGCCAAAAAACATGGACTATAGCAACTATCCCCTTTTTATTCTTGAGTGCCTATGAGACAAACACGAGGATTGCTGCAATTGTCCAAGGAGACAATGAATCAATTGCTATCACTCAAAAAGTTCATCCTAATCTTCCCTACAAGGTAAAGAAAGAGATCTGTGCAAAGCAAGCTCAGCTTTATTTTGAAAGGTTAAGGATGAACTTAAGAGCCCTCGGCCACAATCTTAAAGCTACAGAAACTATCATCAGTACACATCTTTTTATTTATTCGAAGAAAATTCATT |
| 8 | Hantaan orthohantavirus | gaaatTAATACGACTCACTATAgggCAAAGGTTTTTTATGCCCTGAGTTCCCAGGTAGTTTCAGGAAGAAATGCAACTTTGCTACTACTCCTATTTGTGAGTATGACGGAAATATGGTCTCAGGTTACAAGAAAGTAATGGCAACAATTGATTCCTTTCA |
| 9 | Hepatitis C Virus | gaaatTAATACGACTCACTATAgggGAGATACGAATCAATGGCTATATTTGCTGAACGTCTGGATGAGATATACGGTTTACCTGGATTTTTTAATTGGATGCACAAACGACTAGAAAGATCTGTTATCTATGTTGCAGACCCTAATTGCCCCCCTAATATTGACAAACATATGGAACTAGAAAAAACTCCTGAAGATGATATATTCATTCATTATCCTAAAGGCGGTATTGAAGGATATAGCCAAAAAACATGGACTATAGCAACTATCCCCTTTTTATTCTTGAGTGCCTATGAGACAAACACGAGGATTGCTGCAATTGTCCAAGGAGACAATGAATCAATTGCTATCACTCAAAAAGTTCATCCTAATCTTCCCTACAAGGTAAAGAAAGAGATCTGTGCAAAGCAAGCTCAGCTTTATTTTGAAAGGTTAAGGATGAACTTAAGAGCCCTCGGCCACAATCTTAAAGCTACAGAAACTATCATCAGTACACATCTTTTTATTTATTCGAAGAAAATTCATT |
| 10 | RNaseP | GAAATTAATACGACTCACTATAGGGTCCTTGCAGGTGGCTGCCAATACCTCCACCGTGGAGCTTGTTGATGAGCTGGAGCCAGAGACCGACACACGGGAGCCACTGACTCGGATCCGCAACAACTCAGCCATCCACATCCGAGTCTTCAGGGTCACACCCAAGTAATTGAAAAGACACTCCTCCACTTATCCCCTCCGTGATATGGCTCTTCGCATGCTGAGTACTGGACCTCGGACCAGAGCCATGTAAGAAAAGGCCTGTTCCCTGGAAGCCCAAAGGACTCTGCATTGAGGGTGGGGGTAATTGTCTCTTGGTGGCCCAGTTAGTGGGCCTTCCTGA |

**Appendix Table A6:** Sequences of synthetic targets for CARMEN-BBP3

| Number | Pathogen | Oligonucleotide Sequence (5'-3') |
|---|---|---|

| | | |
|---|---|---|
| 1 | Monkeypox Virus | gaaatTAATACGACTCACTATAgggCGTTCCAGCTCCTTTTCGACCTC<br>AATAACTTTAGCACGTTGTCTCATCAAGCTCTCTCTTGTAGTACTAT<br>CATTTTTATCTGATTCCCTGGCACGTTTAA |
| 2 | Hepatitis B Virus | gaaatTAATACGACTCACTATAgggCGCAAAGGCATGGGGACGAATCT<br>TTCTGTTCCCAACCCTCTGGGATTCTTTCCCGATCATCAGTTGGAC<br>CCTGCATTCGGAGCCAACTCAAACAATCCAGATTGGGA |
| 3 | Herpes Simplex Virus Type 2 | gaaatTAATACGACTCACTATAgggATACCAGAAGTCATGTGTATTTTT<br>GAACATCGGTGTCTTTTTATTTATACACAAGCCCAGCTCCCCTCCC<br>CTCCCTTAGAGCTCGTCTTCGTCTCCGGCCTCGTCCTC |
| 4 | Chlamydia trachomatis | gaaatTAATACGACTCACTATAgggTTTGATCTTGGTTCAGATTGAAC<br>GCTGGCGGCGTGGATGAGGCATGCAAGTCGAACGGAGCAATTGT<br>TTCGGCAATTGTTTAGTGGCGGAAGGGTTAGTAATGCATA |
| 5 | Treponema pallidum subsp. pallidum | gaaatTAATACGACTCACTATAgggTACAGTACCCAGCCTATAAGGCA<br>AAAAGGGATAAGACTTCTGCAGAGCTTTATGCGCAAATTCCCCTTA<br>TCGAAGAAATCCTGTGTGCACTGGGCATTACAGTTTTGC |

| | | |
|---|---|---|
| 6 | Trichomonas vaginalis | gaaatTAATACGACTCACTATAgggTGTAAATGTGTGTCAACAACGCACGTTGAATACGTCCCTGCCCTTTGTACACACCGCCCGTCGCTCCTACCGATTGGATGACTCGGTGAAATCACCGGATGCTTACG |
| 7 | RNaseP | GAAATTAATACGACTCACTATAGGGTCCTTGCAGGTGGCTGCCAATACCTCCACCGTGGAGCTTGTTGATGAGCTGGAGCCAGAGACCGACACACGGGAGCCACTGACTCGGATCCGCAACAACTCAGCCATCCACATCCGAGTCTTCAGGGTCACACCCAAGTAATTGAAAAGACACTCCTCCACTTATCCCCTCCGTGATATGGCTCTTCGCATGCTGAGTACTGGACCTCGGACCAGAGCCATGTAAGAAAAGGCCTGTTCCCTGGAAGCCCAAAGGACTCTGCATTGAGGGTGGGGGTAATTGTCTCTTGGTGGCCCAGTTAGTGGGCCTTCCTGA |
